# Machine Learning–based Prediction of LASIK Console Inputs for Aspheric Planning (Q-factor, Defocus, Astigmatism): A Translational Methods Study

**DOI:** 10.64898/2025.12.09.25341758

**Authors:** Steven Garnier

**Affiliations:** Laser Vision Bourgogne, Chalon sur Saone, France

**Keywords:** laser refractive surgery, LASIK, Q-factor, spherical aberration, machine learning, artificial intelligence

## Abstract

**Purpose:** Aspheric planning in laser refractive surgery remains difficult: surgeons often rely on empirical nomograms or simple linear regression for defocus and astigmatism, while console Q-factor modulation yields a variably predictable effect on asphericity and an inconsistent cross-effect on defocus. This translational methods proof-of-concept frames planning as supervised prediction of console-programmable inputs (defocus, astigmatism, Q-factor) and evaluates competing models; it is not a clinical effectiveness study.

**Methods:** We analyzed an anonymized, retrospective, single-platform dataset of 2,448 complete-case treatments. Multi-output regressors (linear and nonlinear) were trained and compared using prespecified metrics (R^2^, MAE/MSE) and residual-distribution visualization/calibration. Actuator–response checks related programmed inputs to changes in defocus (Z_2_^0^) and primary spherical aberration (Z_4_^0^). External validation used a temporally later, device-shift cohort (n=147).

**Results:** Linear regression predicted defocus and astigmatism well (e.g., defocus R^2^=0.98) but degraded for Q-factor (R^2^=0.47), whereas nonlinear models improved Q-factor error and calibration. Actuator–response analyses showed strong coupling for defocus input (R^2^=0.97), moderate coupling of Q-factor to ΔZ_4_^0^ (R^2^=0.51), and a weak Q→defocus cross-effect (R^2^=0.12). On external validation, the best model generalized: Defocus MAE 0.22 D (R^2^=0.98) and Q-factor MAE 0.21 (R^2^=0.81).

**Conclusions:** Supervised nonlinear multi-output models achieve lower error and better calibration for Q-factor than linear baselines, supporting a metric-driven pathway toward more reliable control of low-order refractive targets and primary asphericity. Potential clinical implications include tissue sparing, improved contrast, and near-vision gains. Prospective, human-in-the-loop evaluation with safety and patient-reported endpoints is warranted.

## Introduction

Artificial intelligence (AI) denotes computational systems performing tasks requiring human intelligence; machine learning (ML) is the subfield that learns patterns from data. ML shows promise in medicine,^1–3^ including ophthalmic applications for diagnostic classification,^4^ and regression tasks as intraocular lens power estimation for cataract surgery.^5^ Regression estimates a mapping from predictors to continuous outcomes by minimizing a loss function.

In laser refractive surgery, treatment parameters such as spherical equivalent (defocus) and cylinder (astigmatism) can be estimated using empirical nomograms or linear-regression formulas. Several excimer platforms implement aspheric ablation options—either via an explicit conic-constant programmable aspheric shape “Q-factor” entry or via fixed aspheric compensation—to steer primary spherical aberration (Z_4_^0^) and corneal asphericity (dimensionless; more negative Q indicates a more prolate cornea), with potential benefits including tissue preservation, halo reduction, and modulation of depth of focus^6–8^. A clinical gap persists: the coupling between the programmed Q and the delivered asphericity is weak and context-dependent. Moreover, Q-factor modulation has a weakly predictable cross-coupling on defocus, despite plausible theoretical models.^9^

Against this background, we formulate treatment planning as a multi-output regression problem that directly predicts console-programmable inputs (Q-factor, defocus, and the two orthogonal astigmatism components), and we benchmark higher-complexity ML regressors against traditional linear models. Here, “complexity” denotes model specifications that may trade interpretability for accuracy (e.g., nonlinear, high-parameter, or latent-structure models).

This translational, methods-oriented proof-of-concept bridges algorithmic development and clinical decision-making by recommending console-ready parameters. We focus on out-of-sample accuracy and calibration; to mitigate black-box concerns^10^ we visualize residuals to expose bias, dispersion, and tail behavior. Establishing analytic validity remains the prerequisite to any prospective clinical evaluation.

## Patients and Methods

### Study design

This retrospective translational methods study used anonymized data independent of medical records: no enrollment or follow-up; histories, complications, and outcomes were unavailable and out of scope. All patients provided consent for the use of their data in research.

### Setting and devices

We analyzed a dataset of LASIK treatments performed by a single surgeon (the author) between July 2017 and September 2023 at Laser Vision Bourgogne (Chalon-sur-Saône, France) using an excimer laser (TENEO 317, Model 1, Technolas Perfect Vision GmbH [Bausch + Lomb], Munich, Germany) and a femtosecond laser (FEMTO LDV Z2, Ziemer Ophthalmic Systems AG, Port, Switzerland).^11^ We also used a later dataset (Sep 2023–Aug 2024) from the same center and surgeon with a second excimer laser (TENEO 317, Model 2, Technolas Perfect Vision GmbH [Bausch + Lomb], Munich, Germany). This validation set reflects a temporal and device shift under matched protocols. Only spherical and aspheric (Q-factor) ablation profiles were used; neither presbyopia-mode nor topography-guided nor aberrometry-guided (wavefront-guided) treatments were performed.

### Derivation and Validation

Data were split chronologically (derivation n=2448; external validation n=147).The derivation dataset underwent k-fold nested cross-validation with all preprocessing and selection refit inside folds to prevent leakage; the external validation dataset was held out and evaluated once.

### Features

For each eye-level instance, we collected 254 objective features, grouped by timepoint and modality: 107 preoperative, 107 postoperative and 40 intraoperative variables (optical zone, maximum/ central ablation, treatment area, total pulse, duration, etc.). Pre-/postoperative measures included 67 biometric/keratometric parameters (Kmax/Kmin, axes, 3- and 5-mm zones keratometric data, geometric descriptors from anterior/posterior corneal maps, pachymetry, thinnest point, white-to-white, anterior chamber depth) acquired on scanning-slit elevation topographer (ORBSCAN 3, Technolas Perfect Vision GmbH [Bausch + Lomb], Munich, Germany) and 40 aberrometric variables (Zernike polynomial coefficients; total and high-order RMS, PPR 3.5mm [predicted phoropter refraction], etc.) obtained under cycloplegia (tropicamide 1% ophthalmic solution) on a Shack-Hartmann wavefront-sensor aberrometer (ZY-WAVE 3, Technolas Perfect Vision GmbH [Bausch + Lomb], Munich, Germany). Full definitions are provided in Supplementary Tables S1-S3.

### Feature selection and model size

After transforming and standardizing the 254 features, substantial collinearity indicated that unconstrained models would overfit, destabilize coefficients, and degrade out-of-sample calibration; dimensionality reduction was therefore required. Starting from the prespecified 10-variable core, we targeted ≈10 total inputs under L1 (lasso) regularization and ≈10–15 under L2 (ridge), consistent with effective degrees of freedom at our sample size. Candidates were screened by MANOVA (Pillai’s trace) with Bonferroni multiplicity correction. Final inclusion required incremental out-of-fold improvement under forward addition within nested cross-validation, stopping when Root Mean Squared Error (RMSE) and Mean Absolute Error (MAE) change is below 0.01 D. Only a small, domain-guided subset was inspected manually; just two additional variables met criteria beyond the core set. Retained variables are listed in Table 1.

**Table 1.** Summary of the input “features” (X) and the predicted outputs “target labels” (y) distributions.

| X: 12 inputs/features (units) | Pre – op Asphericity (μmRMS) | Pre – op Defocus (D) | Pre – op Astigm + (D) | Pre – op Astigmatx (D) |
| --- | --- | --- | --- | --- |
| Mean | +0.17 | -0.48 | -0.12 | -0.03 |
| Standard Deviation | 0.17 | 2.94 | 0.95 | 0.56 |
| Range [min; max] | [-0.63 ; +1.12] | [-13.12 ; +8.42] | [-5.69 ; +4.08] | [-3.75 ; +2.72] |
|  | Post – op Asphericity (μmRMS) | Post – op Defocus (D) | Post – op Astigm + (D) | Post – op Astigmatx (D) |
|  | +0.01 | -0.05 | -0.02 | -0.01 |
|  | 0.26 | 0.73 | 0.42 | 0.34 |
|  | [-0.83 ; +1.14] | [-3.74 ; +2.69] | [-2.13 ; +4.75] | [-2.03 ; +1.32] |
|  | Optical Zone (Q – factor) (mm) | Optical Zone (Defocus) (mm) | Pre – op Km (D) | Pre – op Q (no unit) |
|  | 6.93 | 6.97 | 43.82 | -0.21 |
|  | 0.65 | 0.59 | 1.64 | 0.24 |
|  | [5.70 ; 8.50] | [5.70 ; 8.50] | [34.20 ; 52.90] | [-1.62 ; 3.15] |
| Y: 4 outputs/labels (units) | Q – factor $yZ_4^0$ (no unit) | Defocus $yZ_2^0$ (D) | Astigm + $yZ_2^{-2}$ (D) | Astigmatx $yZ_2^{+2}$ (D) |
| Mean | +0.42 | -0.36 | -0.25 | -0.00 |
| Standard Deviation | 0.81 | 2.54 | 0.89 | 0.47 |
| Range [min; max] | [-0.50 ; +3.00] | [-8.18 ; +6.65] | [-5.01 ; +3.94] | [-3.22 ; +3.04] |

### Quantization

Because regression requires sufficiently resolved target, we did not use manifest refraction, which is constrained to coarse console increments (0.25 D; 5°). Such discretization renders the regression problem statistically underidentified and imposes a quantization-induced irreducible error floor (for step Δ, MAE ≥ Δ/4, RMSE ≥ Δ/√12; e.g., MAE ≥ 0.0625 D, RMSE ≥ 0.072 D for Δ = 0.25 D), yielding staircase fits and degraded calibration. Accordingly, we trained and evaluated on the high-resolution measurements available in our dataset (0.01 D; 1°), for which the floor is negligible (e.g., MAE ≥ 0.0025 D, RMSE ≥ 0.0029 D).

### Instrument repeatability

To quantify the irreducible measurement error of the ShackHartmann aberrometer^12–14^, we acquired 100 consecutive measurements (technical replicates) from one healthy emmetropic eye under three conditions: (i) natural pupil (no cycloplegia or dilation), (ii) cycloplegia (tropicamide 1%), and (iii) pharmacologic mydriasis without cycloplegia (phenylephrine 10%). All other conditions (alignment, fixation, operator, session) were held constant. For each condition, intrasession repeatability was summarized by the within-subject standard deviation S_w_. We interpret S_w_ as the instrument’s intrasession noise floor.

### Orthogonal-basis optical feature encoding

Raw spherocylindrical data were recast into three orthogonal power-vector components, according to Thibos’ method—spherical equivalent M, and the cardinal/ oblique astigmatic components J_0_ and J_45_ (diopters)^15,16^—which, in the Zernike formalism, correspond to the lower-order (2^nd^ order) terms for defocus and astigmatism (Z_2_^0^, Z_2_^−2^, Z_2_^+2^). For readability we label these “Defocus” (Z_2_^0^), “Astigm+” (Z_2_^−2^), and “Astigmx” (Z_2_^+2^). The primary asphericity measurement (Z_4_^0^) over a 6-mm pupil (µm RMS)—denoted “6mmZ400” in the aberrometer interface—is concatenated for each instance. Consequently, the aberrometric preoperative state is defined by a vector Z_preop_ = (Z_4_^0^, Z_2_^0^, Z_2_^−2^, Z_2_^+2^) and the postoperative state by a vector Z_postop_ = (Z_4_^0^, Z_2_^0^, Z_2_^−2^, Z_2_^+2^) of 4 normalized variables, as per the Zernike formalism. This orthogonal basis preserves proximity to the instrument’s internal Zernike fit and reduces collinearity, conditionally stabilizing estimation on the present dataset. We standardized predictors and applied a fixed, orthonormal 2×2 sum–difference (Hadamard) rotation to each pre–post pair to reduce collinearity and attenuate common-mode measurement noise. This transform is equivalent to principal component analysis (PCA) in the pre–post plane when variances are comparable and common-mode noise dominates.

### Clinical viewpoint

(Figure 1). The clinical reasoning is forward and causal: the preoperative state Z_preop_ is observed first; then the surgeon sets the console inputs y = (yZ_4_^0^, yZ_2_^0^, yZ_2_^−2^, yZ_2_^+2^) corresponding to (Q-factor, defocus, and the two astigmatism components Astigm+ and Astigmx), and the laser causally transforms Z_preop_ into Z_postop_.

**Figure 1.**
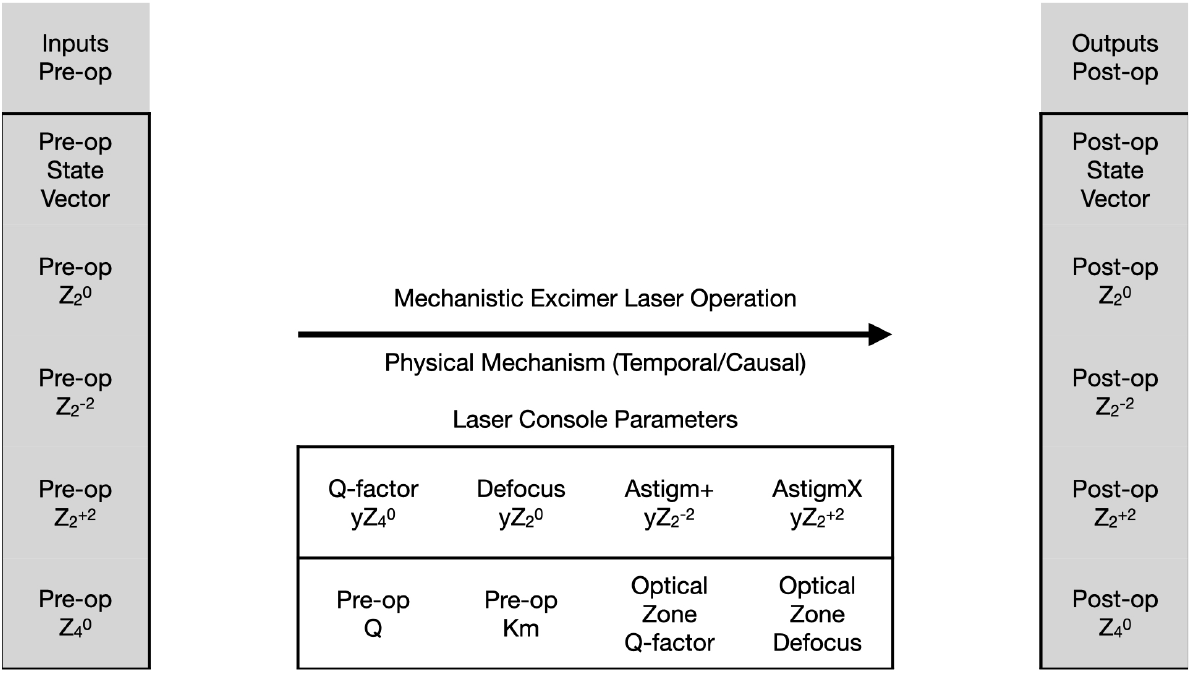
Mechanistic representation of the data: pre-operative state, intra-operative laser settings, and post-operative outcomes are shown in their temporal/causal ordering

### Data-science viewpoint

(Figure 2). We collect many triplets (Z_preop_, y, Z_postop_) from past cases. We learn a supervised mapping that, given (Z_preop_, Z_postop_), predicts the programmed console inputs (y) (inverse identification). These four variables serve as the target labels for the supervised ML model, as described in Table 1.

**Figure 2.**
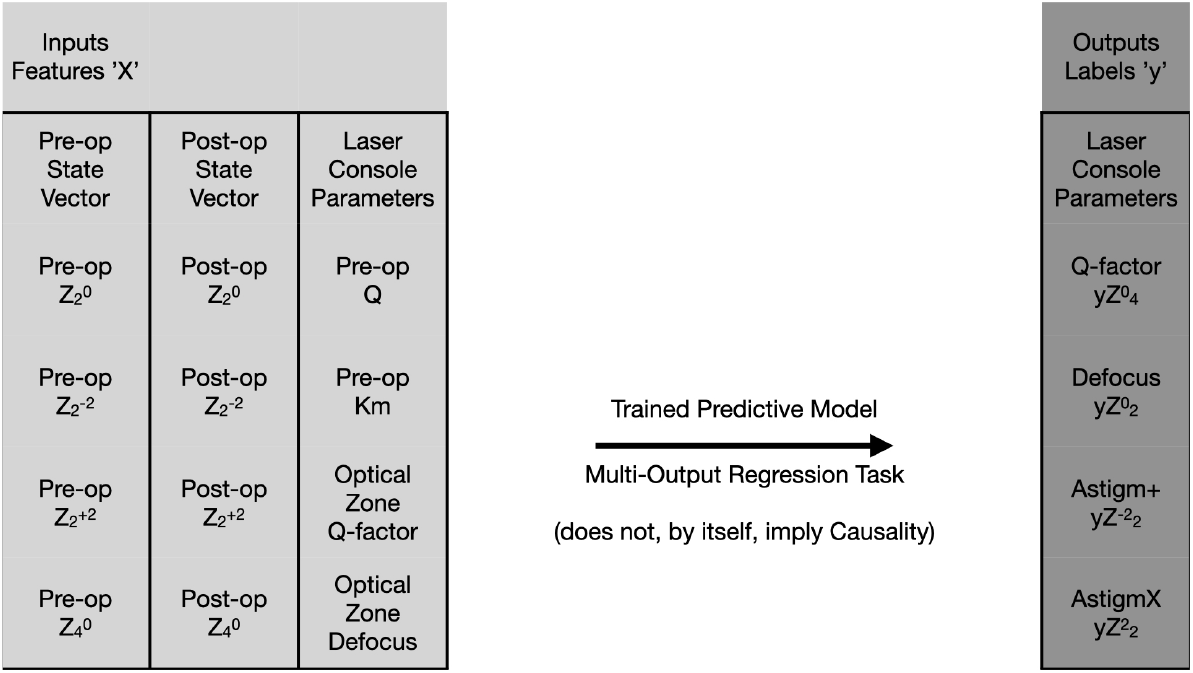
Supervised-learning representation of the data: Features and Labels formalize a Prediction Task to predict laser console parameters. (This representation does not, by itself, establish causal relations).

### Target variables

Outputs are restricted to the surgeon-programmable, orthogonal console parameters y = (yZ_4_^0^, yZ_2_^0^, yZ_2_^−2^, yZ_2_^+2^). Each output maps directly to a single console field, ensuring clinical actionability, statistical identifiability and platform portability. Higher-order controls were not available on the studied platform. We use achieved postoperative states (not intended aims) as part of the input information and assess performance by the signed prediction error (residual) defined as the difference between predicted and the programmed console inputs (ground truth). The approach is agnostic to the intended refractive aim or clinical goals and accommodates deliberate non-zero settings (e.g., monovision, intentional Q for depth-of-focus, tissue-sparing trade-offs).

### Empirical actuator–response checks

We first regressed the pre- to postoperative change in defocus (ΔZ_2_^0^) on the programmed Defocus input. Next, we related the change in primary spherical aberration (ΔZ_4_^0^) to the programmed Q-factor within a negligible defocus subset (±0.75 D). Finally, we assessed the cross-coupling by regressing (ΔZ_2_^0^) on the programmed Q-factor within a near-null defocus subset (±0.25 D), pooling all optical zone diameters in a single panel. On this console, increasing the Q-factor settings is intended to produce a more prolate aspheric profile.

### Model comparaison

Table 2 summarizes model classes: LINR, LINear Regression; BAGB^17,18^, Bagging Aggregator (Gradient Boosted); VMLP^19^, Vanilla Multilayer Perceptron; BDMH^20–24^ Bayesian Dynamic MultiHead neural network. Models were trained under different loss functions (MSE, MAE, Huber^25^). For each target label, we report the coefficient of determination (R^2^) and MSE; for clinical interpretability, we also report MAE. To characterize error distributions, we report the median absolute error (MedAE) with interquartile range [25th, 75th] and central 95% interval [2.5th, 97.5th] of absolute errors. All metrics are presented on the original (denormalized) scale (Table 3). Because summary statistics can mask heavy-tailed or heteroscedastic regimes, we complement metrics with residual-distribution and calibration diagnostics for each target and model (bias, dispersion, skewness, kurtosis) to flag tail risk and miscalibration with direct clinical implications (safety margins, avoidable retreatment).

**Table 2.** Model glossary (acronyms, full names, and descriptions)

| Acronym | Full name | Description |
| --- | --- | --- |
| LINR | Linear Regression Model | “Modeling” denotes specifying and estimating a data-driven mapping from predictors to targets. “Supervised” learning denotes learning a predictive mapping from labeled examples. “Linear” predicts outcomes with a straight-line formula learned from data; each outcome has its own weights and an intercept. |
| BAGB | Bootstrap Aggregator Gradient-Boosted | Nonlinear: input–output relation not constrained to be affine; allows curvature and interactions. “Ensemble” methods: a bagged forest of nonlinear gradient-boosted decision trees; each boosted model is trained on a bootstrap sample and predictions are averaged. |
| VMLP | Vanilla Multi-Layer Perceptron (neural network) | Canonical feed-forward neural network with fully connected layers; “deep learning” denotes stacking multiple layers; trained by gradient descent to learn nonlinear relations. |
| BDMH | Bayesian Dynamic Multi-Head neural network | Bayesian deep multi-output architecture with a shared backbone and task-specific heads, trained by variational inference with learned task-wise noise scales for calibrated predictions, augmented with structuring latent factors that enter simultaneous structural equations, and meta-learned via out-of-fold nonlinear stacking to fuse complementary base models. |

**Table 3.** Comparative unscaled metrics for each model across four predicted outputs (target labels)

| $R^2$<br>MSE (p-value)<br>MAE (p-value)<br>MedAE<br>IQR [25th-75th]<br>95%ci [2.5-97.5] | Q-Factor $yZ_4^0$ | Defocus $yZ_2^0$ | Astigm+ $yZ_2^{-2}$ | Astigmx $yZ_2^{+2}$ |
| --- | --- | --- | --- | --- |
| LINR<br>Linear Regressor | 0.47<br>0.40 (p=ref)<br>0.51 (p=ref)<br>0.44<br>[-0.46; 0.42]<br>[-1.18; 1.30] | 0.98<br>0.13 (p=ref)<br>0.28 (p=ref)<br>0.22<br>[-0.21; 0.23]<br>[-0.75; 0.72] | 0.92<br>0.06 (p=ref)<br>0.18 (p=ref)<br>0.13<br>[-0.13; 0.13]<br>[-0.45; 0.48] | 0.79<br>0.05 (p=ref)<br>0.16 (p=ref)<br>0.13<br>[-0.14; 0.13]<br>[-0.43; 0.41] |
| BAGB<br>Bootstrap Aggr.<br>Gradient Boosting | 0.66<br>0.26 (p=0.00)<br>0.37 (p=0.00)<br>0.28<br>[-0.28; 0.28]<br>[-1.02; 0.98] | 0.98<br>0.13 (p=0.24)<br>0.27 (p=0.00)<br>0.21<br>[-0.20; 0.21]<br>[-0.71; 0.71] | 0.93<br>0.06 (p=0.24)<br>0.16 (p=0.00)<br>0.12<br>[-0.12; 0.11]<br>[-0.46; 0.43] | 0.77<br>0.05 (p=0.00)<br>0.16 (p=0.42)<br>0.12<br>[-0.11; 0.12]<br>[-0.47; 0.46] |
| VMLP<br>Vanilla<br>MultiLayerPerceptron | 0.65<br>0.27 (p=0.00)<br>0.36 (p=0.00)<br>0.24<br>[-0.31; 0.18]<br>[-1.16; 0.98] | 0.98<br>0.13 (p=0.55)<br>0.27 (p=0.01)<br>0.21<br>[-0.21; 0.21]<br>[-0.75; 0.71] | 0.93<br>0.05 (p=0.01)<br>0.17 (p=0.00)<br>0.13<br>[-0.13; 0.12]<br>[-0.45; 0.45] | 0.77<br>0.05 (p=0.00)<br>0.17 (p=0.00)<br>0.13<br>[-0.12; 0.14]<br>[-0.45; 0.48] |
| BDMH<br>Bayesian Dynamic MultiHead<br>Neural Network | 0.71<br>0.22 (p=0.00)<br>0.31 (p=0.00)<br>0.22<br>[-0.22; 0.22]<br>[-0.95; 0.97] | 0.98<br>0.11 (p=0.00)<br>0.25 (p=0.00)<br>0.18<br>[-0.18; 0.19]<br>[-0.70; 0.66] | 0.94<br>0.05 (p=0.00)<br>0.16 (p=0.00)<br>0.12<br>[-0.12; 0.12]<br>[-0.44; 0.45] | 0.87<br>0.03 (p=0.00)<br>0.11 (p=0.00)<br>0.08<br>[-0.08; 0.08]<br>[-0.34; 0.35] |

The Python libraries SciPy, scikit-learn, PyTorch, Pyro and semopy were used.

Datasets and analysis scripts are available from the corresponding author upon reasonable request and subject to institutional approval.

## Results

### Datasets

The training/calibration set (complete cases with matched pre- and postoperative aberrometry, no missing values) comprised 2,448 eyes: 1728 (70.6%) aspheric and 720 (29.4%) spherical ablations. Feature and label distributions exhibited broad, continuous support. The external validation set included 147 complete-case eyes of similar quality—88(59.9%) aspheric and 59(40.1%) spherical.

### Instrument repeatability

Within-subject standard deviations S_w_ for the Shack-Hartmann aberrometer, reported for (Z_4_^0^, Z_2_^0^, Z_2_^−2^, Z_2_^+2^) were:

- undilated pupil: (not applicable, 0.26D, 0.09D, 0.18D)
- pharmacologically dilated pupil: (0.030μm RMS, 0.25D, 0.09D, 0.05D)
- cycloplegia: (0.015μm RMS, 0.15D, 0.09D, 0.09D).

These values quantify the intrasession noise floor for each condition.

### Actuator–response relationships

Defocus input strongly explained ΔZ_2_^0^ (R^2^=0.97; Figure 3), indicating that the gains and offset for laser calibration are accurately set. By contrast, Q-factor explained ~51% of

**Figure 3.**
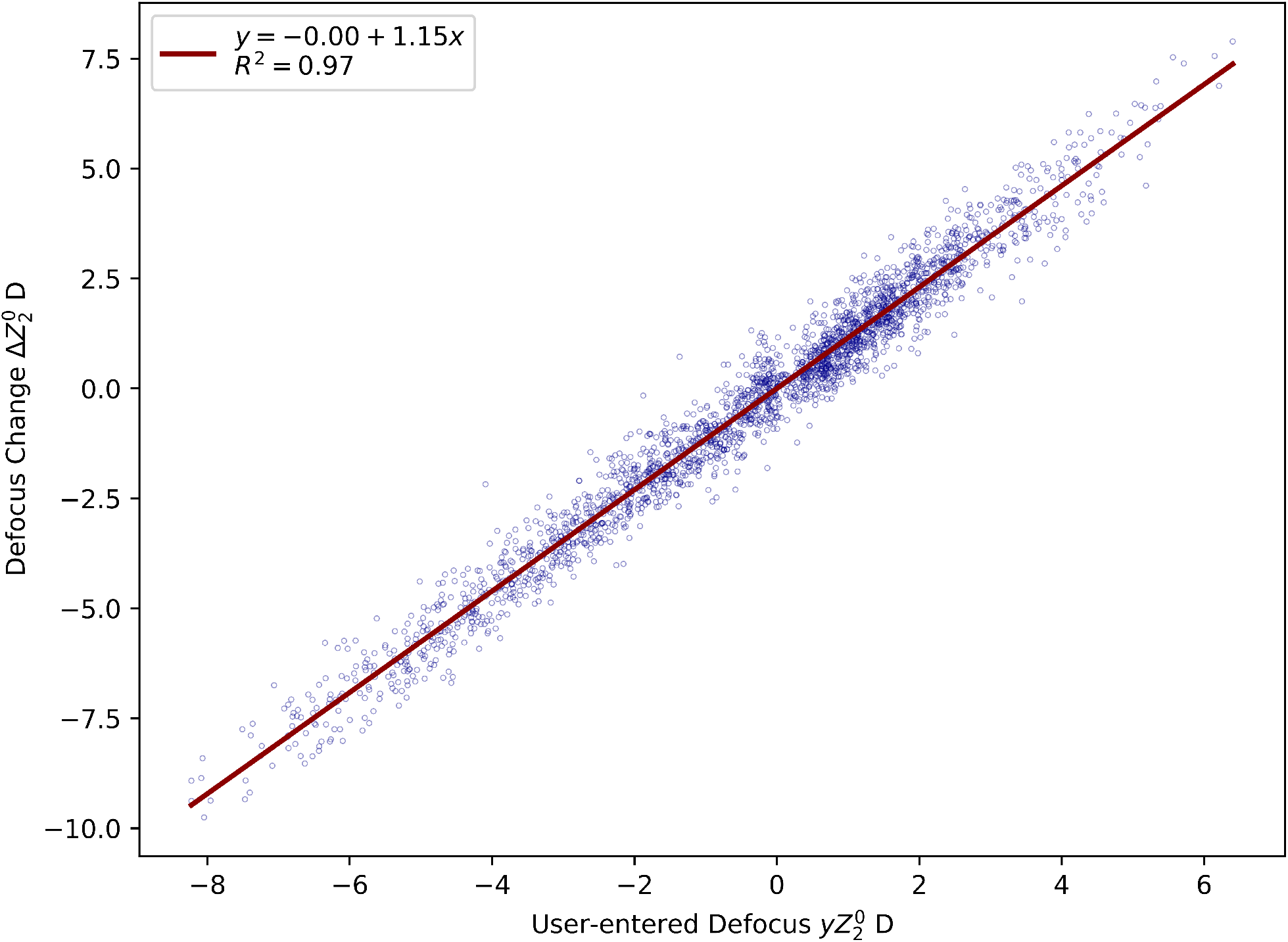
Defocus actuator–response. Programmed Defocus (D) vs postoperative change in defocus, ΔZ_2_^0^ (D); (R^2^=0.97). Each point represents one eye: (n=2448)

ΔZ_4_^0^ (Figure 4). In a near-null-defocus subset, Q-factor weakly explained ΔZ_2_^0^ (R^2^=0.12; Figure 5), implying poor identifiability without covariates (optical zone, Km, pre-op Q).

**Figure 4.**
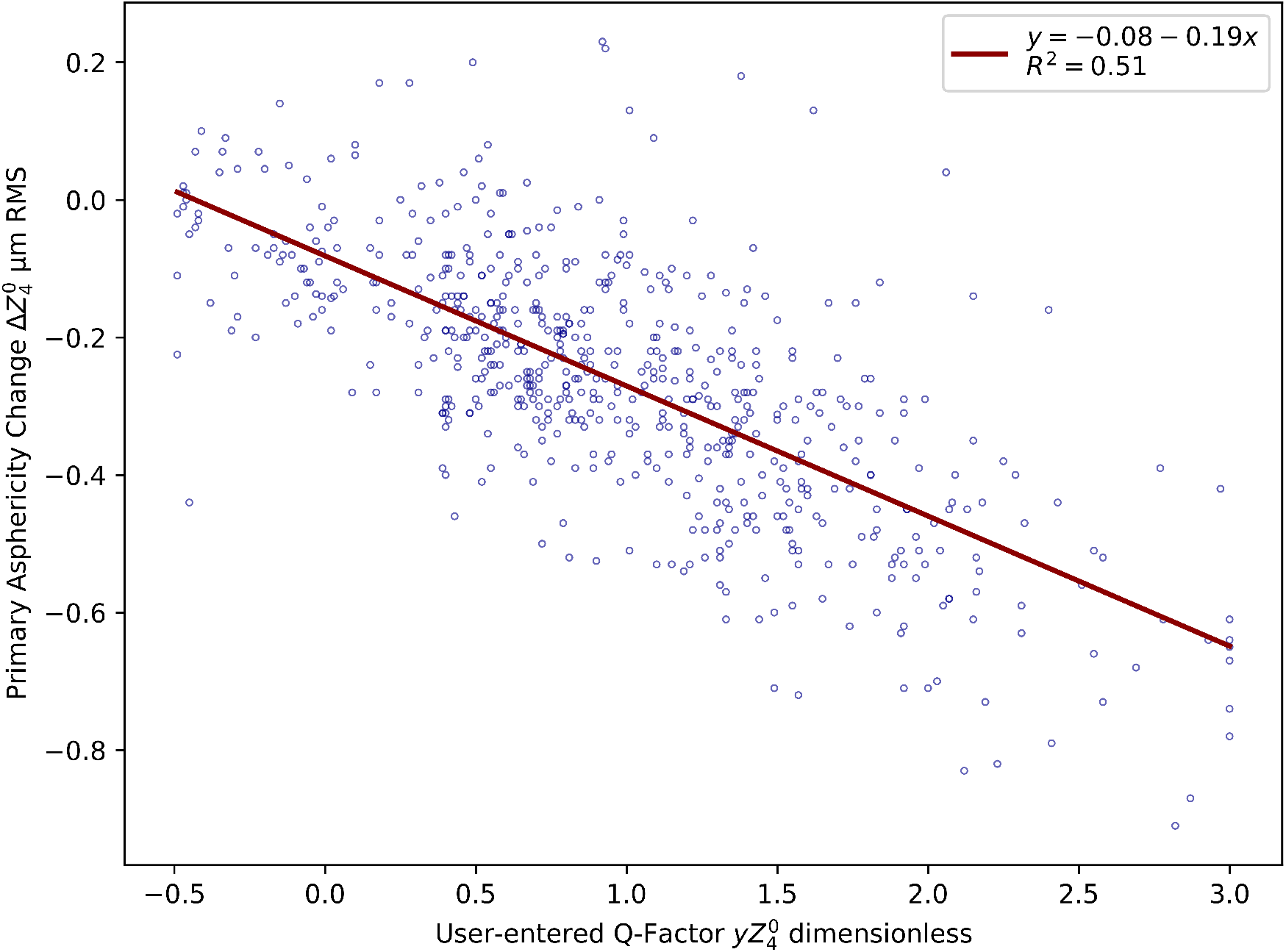
Asphericity actuator–response. Programmed Q-factor (dimensionless) vs change in primary spherical aberration, ΔZ_4_ ^0^ (µm RMS; 6-mm pupil), restricted to cases with near-null Defocus input (|Defocus|≤0.75D); (R^2^=0.51). (n=606)

**Figure 5.**
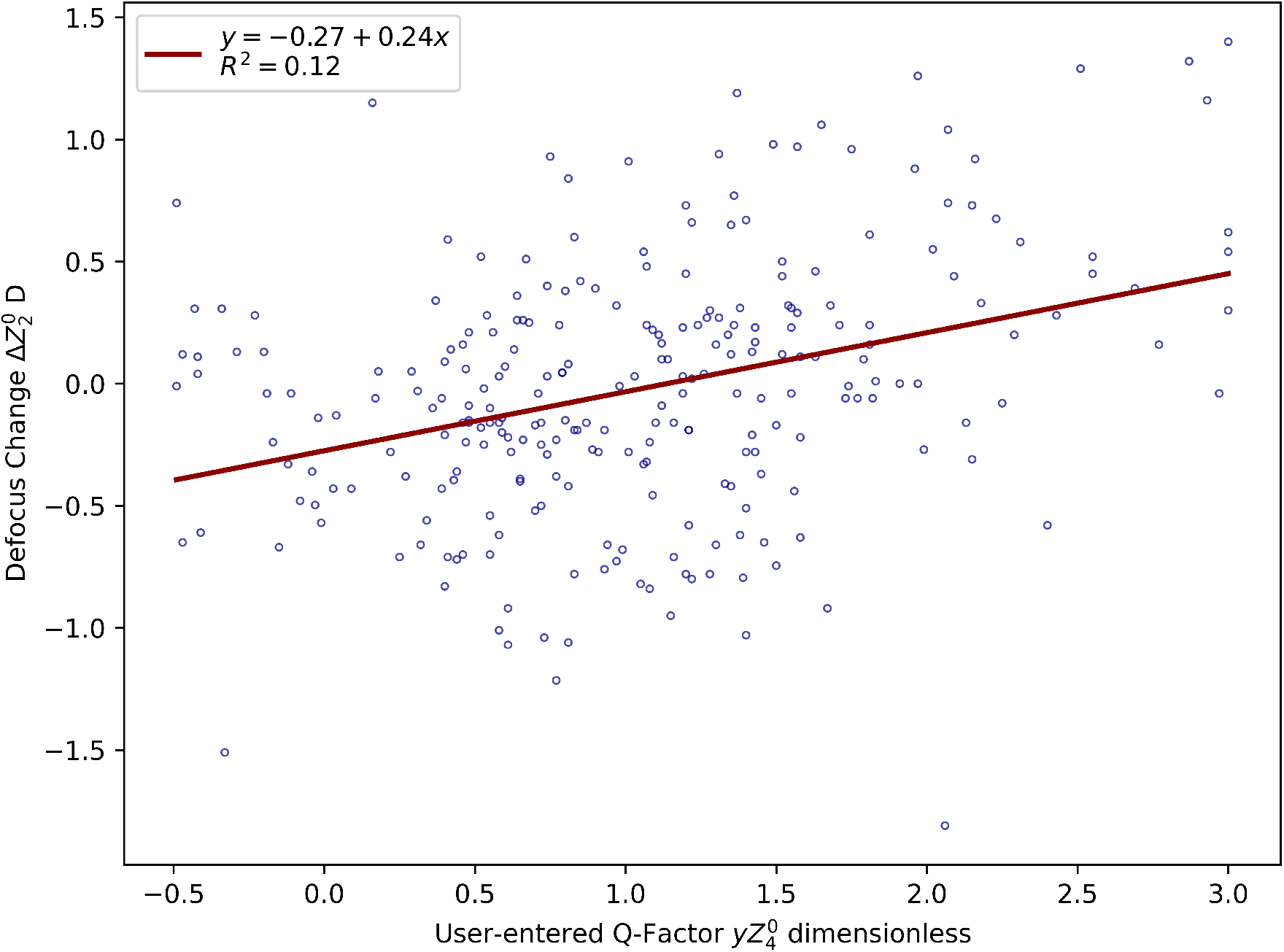
Cross-coupling of Q-factor on defocus. Programmed Q-factor vs change in defocus, ΔZ_2_^0^ (D), restricted to a near-null Defocus subset (|Defocus|≤0.25D); (R^2^=0.12); (n=259)

### Model performance

Table 3 summarizes predictive metrics. Linear regression (LINR) predicted Defocus accurately (R^2^=0.98) but underperformed for the Q-factor (R^2^=0.47). Nonlinear models (BAGB, VMLP, BDMH) achieved substantially better fits for the Q-factor target, while maintaining high accuracy for Defocus (Supplementary Notes S1–S4).

### Residual distributions

(Figure 6). For each target/model, we characterize the residuals by the first four moments:

**Figure 6.**
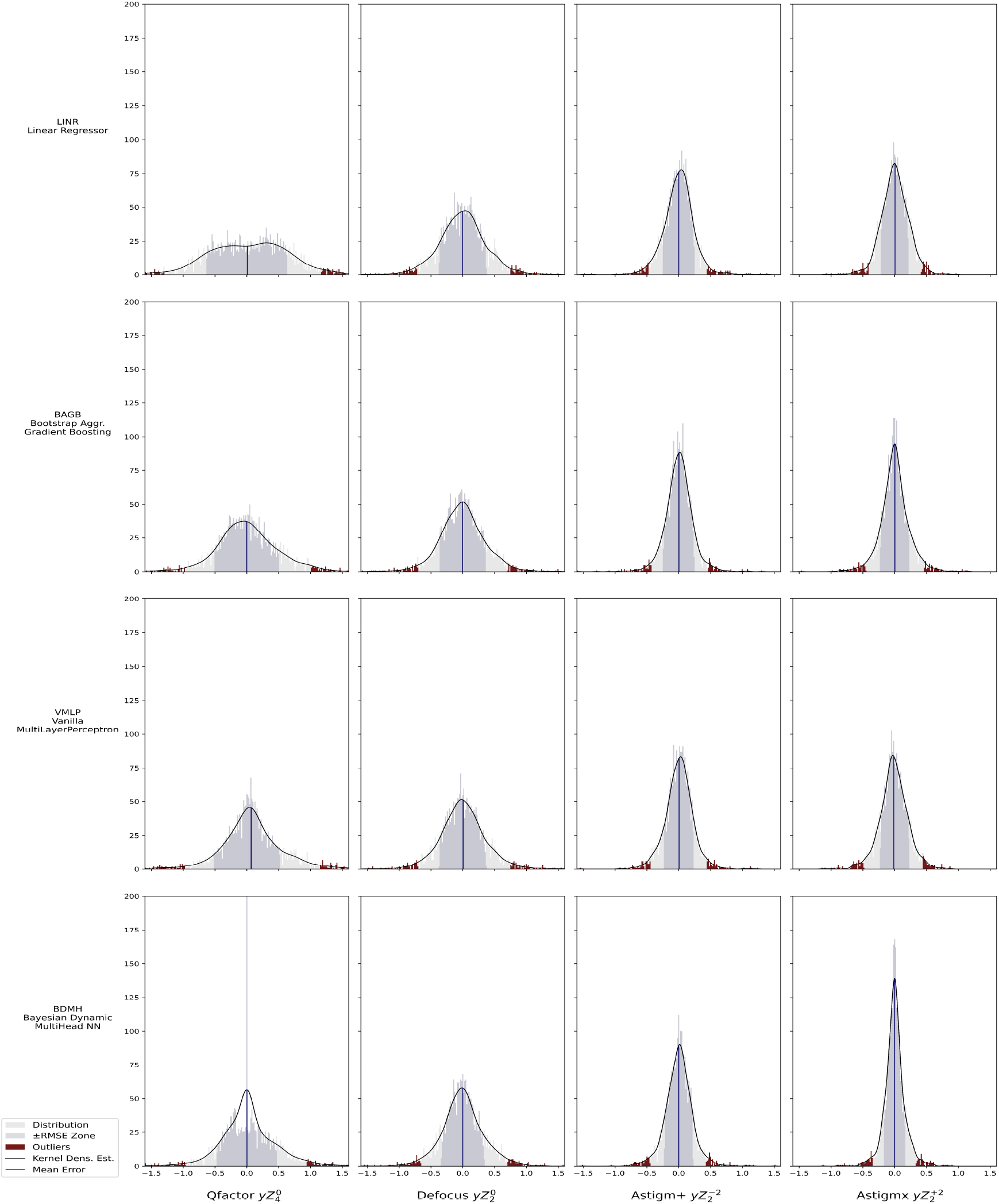
Residual distributions for each target–model pair: histograms with kernel density estimate (KDE) overlay; vertical line marks the mean error; shaded band denotes±RMSE; outliers are flagged

- Mean (bias). Vertical reference (blue line) indicates mean error: all mean errors are zero or nearly zero (no systematic bias) except VMLP on Q-factor, which shows a non-zero bias (systematic error).
- Variance (dispersion). The central gray band depicts a ±RMSE, visualizing dispersion on the original scale.
- Skewness (asymmetry). Kernel density estimates (KDE) reveal asymmetry—for example, BAGB on Q-factor shows skewed residuals suggestive of systematic under- or over-prediction in a tail.
- Kurtosis^27^ (tail weight). KDE shapes indicate tail behavior: LINR on Astigmx is more peaked with heavier tails (leptokurtic), consistent with MAE-like losses (L1 norm); BDMH on Defocus is close to Gaussian (L2 norm); Astigmx targets tend to show elevated kurtosis across models, suggesting residual underfitting in this axis; LINR on Q-factor shows an irregular density, pointing to a specification mismatch.

A residual distribution that is centered (≈0 bias), symmetric (low skew), Gaussian-like (moderate kurtosis), and narrow (low variance) signals well-behaved errors—observed most clearly for BDMH on Defocus.

### External validation

On the held-out temporal/device-shift set (n=147), BDMH generalized well: Defocus MAE = 0.22 D with R^2^=0.98; Q-factor MAE = 0.21 (dimensionless) with R^2^=0.81.

## Discussion

### Study scope and design

This preliminary, retrospective, translational methods study does not claim co-efficient-level interpretability or causal effects; such claims would require randomized interventions or a prespecified structural causal model. Prospective testing of ML-derived console recommendations is pre-mature until analytic validity—robust out-of-sample accuracy and calibration—has been demonstrated on chronologically external cohorts. Because clinical outcomes and complication annotations were unavailable, we cannot attribute tail errors to specific surgical events (e.g., eccentric flaps, biomechanical regression, complications); registry-level prospective studies with adjudicated events will be required to quantify their contribution to residual tails. The appropriate next step is a prospective, human-in-the-loop evaluation with endpoints sensitive to asphericity—mesopic contrast, halos, and depth of focus—plus patient-reported outcomes and safety metrics (e.g., retreatment), accompanied by per-case uncertainty (e.g., split conformal prediction intervals). Several prior AI studies in refractive surgery report methodological limitations, underscoring the need for stronger designs.^28–33^

### Manifest refraction as target

Forgoing manifest refraction as the direct target is a recognized limitation. Methodologically, however, its coarse discretization (≥0.25 D; 5°) creates a structural information deficit: no downstream algorithm can reconstruct sub-Δ detail that was never observed, and ordinal classification does not resolve this. A pragmatic strategy is two-layered: (i) learn on high-resolution, continuous targets when available; then (ii) append a discrete, clinically aligned decision layer calibrated to manifest refraction for deployment.Zernike orthogonality is idealized here; instrumental factors (aperture mismatch, centration/tilt relative to the instrument axis, wavefront reconstruction noise) and surgical/biomechanical effects can induce cross-coupling among coefficients. Hence, statistical independence of the estimated coefficients cannot be assumed. Our joint models—including a hierarchical Bayesian structural specification —already capture part of the reciprocal Q-factor↔defocus coupling. Future work will impose stronger cross-coupling constraints and validate these constraints across devices and populations using partial pooling (Supplementary Notes S5).

### Features and generalizability

We did not execute a fully automated, exhaustive feature search; weak higher-order synergies (e.g., pachymetry, epithelial/biomechanical metrics) may be missed. Platform dependence is therefore a central limitation. We consider several deployment axes:

- Surgeon shift, same device. Minimal covariate shift; periodic monitoring and light recalibration are likely sufficient.
- Site shift, same platform. Core structure should persist; we recommend local recalibration and device-step mapping, guided by bias–slope residuals on the target device for a simple, auditable post hoc correction. Federated learning could enable privacy-preserving model updates across institutions without exchanging raw patient data.
- Device shift (hardware/model/firmware). The framework and metrics are portable, but retraining on device-specific data is required; the feature set may expand to include device-exposed controls (e.g., higher-order terms).
- Cross-vendor deployment. Not recommended at present, even with physics-based, non-proprietary features. Multi-center pooling alone does not guarantee better generalization.

### Clinical implementation

Beyond periodic recalibration, we advocate (i) device-level guardrails (e.g., minimum residual stromal bed, optical zone constraints) that enforce safety at entry time; (ii) prespecified abstention rules backed by out-of-distribution detection; (iii) a multi-output coherence filter; and (iv) shadow-mode operation that logs safety metrics for prospective auditing and drift monitoring (all standard good practices). In deployment, per-case uncertainty can be provided via split conformal prediction, yielding distribution-free marginal 95% coverage under exchangeability; this procedure was not implemented or evaluated in the present study.

### Workflow and clinical relevance

Operationally, the system ingests six preoperative values—PPR 3.5mm refraction (sphere and astigmatism), primary spherical aberration (Z_4_^0^), mean keratometry (Km), and preoperative corneal asphericity (Q)—along with the four desired postoperative aims (target sphere, astigmatism and target Z_4_^0^) and the two predefined treatment zones (spherical, aspheric). The multi-output model then returns four console-ready inputs mapped back to clinical terms as Q-factor, sphere, cylinder, and axis (per-case prediction intervals are a deployment option; see Clinical implementation).

### Empirical starting point

Programmed Q-factor weakly controls the delivered Z_4_^0^ and its cross-coupling on Z_2_^0^ is effectively unpredictable in both magnitude and sign. If the actuator–response coupling is weak for the first HOA—then achieving tighter control over higher-order modes with fixed wavefront-guided profiles is a fortiori unlikely; coarse refractive endpoints will rarely register a benefit^34,35^ (outside pathological cases of a different order of magnitude).

### Why accurate Q-factor prediction matters

Better Q-factor prediction tightens control of delivered asphericity: in presbyopes, accurate Q-factor allows targeting the magnitude and sign of Z_4_^0^ to extend depth-of-focus. In eyes with large scotopic pupils or small optical zones, a well-calibrated Q-factor can temper the spherical aberration induced, mitigating halos without overcorrecting sphere. Under stromal-bed constraints, appropriate Q-factor adjustment may allow the same spherical aim with reduced central tissue removal, potentially improving safety margins. Because real cases often require all three levers at once, nonlinear, well-calibrated Q-factor prediction enables patient-specific trade-offs.

### Why nonlinear, multidimensional structure matters

Recommended Q-factor (and, to a lesser extent, spherocylindrical entries) varies nonlinearly and depends on interactions among optical zone diameter and preoperative optics—relationships that human heuristics and fixed, low-dimensional nomograms struggle to capture reliably. Nonlinear, multidimensionnal learning captures these couplings.

### Why multi-output prediction matters

The console entries are physically coupled: changing defocus induces or attenuates asphericity; reciprocally, tuning Q-factor shifts defocus; and these nonlinear cross-coupling depend on the optical zones. Predicting Q-factor, defocus, and the two astigmatism components jointly preserves this coupling.

Conceptually, the data-generating mechanism is better represented by a system of simultaneous structural equations with reciprocal defocus–asphericity dependencies, with observations subject to non-negligible measurement noise. These dependencies arise from physical cross-coupling, measurement-model non-orthogonality, and population-level covariance.

### Limitations and future directions

Key limitations include the retrospective single-surgeon/single-center setting, absence of clinical outcomes/complications, residual confounding, and platform specificity; thus, transportability to other sites/devices remains to be established. Future work should address: (i) prospective, human-in-the-loop trials with safety and patient-reported endpoints; (ii) validation of aberration-coefficient coupling (e.g., defocus–asphericity) and measurement repeatability across platforms; (iii) distribution-aware uncertainty quantification (e.g., heteroscedastic modeling and Mondrian conformal prediction) and optimization of feature-space denoising (generalized least squares, matrix factorization) to preserve subgroup-level coverage; (iv) explicit structural/causal modeling of the actuator–response map (simultaneous equations with reciprocal defocus↔asphericity terms), with physics-informed constraints; and (v) standardized, device-agnostic audit trails enabling traceable post hoc recalibration.

Findings support calibrated, multi-output prediction that respects optical coupling. Building a large, anonymized benchmark dataset—with continuous coverage across clinical ranges, documented provenance, and open protocols—would enable transparent, reproducible comparisons and accelerate clinically relevant advances in refractive-surgery modeling.^36^

## Data Availability

All data produced in the present study are available upon reasonable request to the authors

