## Supplemental Table 1 for "Machine Learning–based Prediction of LASIK Console Inputs for Aspheric Planning (Q-factor, Defocus, Astigmatism): A Translational Methods Study": SUP TABLE 1.pdf

| Device | Features | Category | Unit |
|---|---|---|---|
| TENEO | orbscantime | Time | Unix datetime |
| TENEO | zywavetime | Time | Unix datetime |
| TENEO | calculationtime | Time | Unix datetime |
| TENEO | surgerytime | Time | Unix datetime |
| TENEO | side | ID | OS/OD |
| TENEO | subjectiverefraction_sphere | Refraction | Diopter (D) |
| TENEO | subjectiverefraction_cylinder | Refraction | Diopter (D) |
| TENEO | subjectiverefraction_axis | Refraction | Axis (°) |
| TENEO | PPR_sphere | Refraction | Diopter (D) |
| TENEO | PPR_cylinder | Refraction | Diopter (D) |
| TENEO | PPR_axis | Refraction | Axis (°) |
| TENEO | correctionrefraction_sphere | Refraction | Diopter (D) |
| TENEO | correctionrefraction_cylinder | Refraction | Diopter (D) |
| TENEO | correctionrefraction_axis | Refraction | Axis (°) |
| TENEO | 6mmZ400 | Aberrometry | µm RMS |
| TENEO | highorderRMS | Aberrometry | µm RMS |
| TENEO | preopKm | Topography | Diopter |
| TENEO | preopQ | Topography | No unit |
| TENEO | pachymetry | Topography | µm |
| TENEO | nomogram | Treatment parameter | % |
| TENEO | opticalzone | Treatment parameter | mm |
| TENEO | Qfactor | Treatment parameter | No unit |
| TENEO | mode | Treatment parameter | Category |
| TENEO | targetsphere | Treatment parameter | Diopter |
| TENEO | flapthickness | Treatment | µm |
| TENEO | maxablation | Treatment | µm |
| TENEO | centralablation | Treatment | µm |
| TENEO | esidualstroma | Treatment | µm |
| TENEO | area_x | Treatment | mm |
| TENEO | area_y | Treatment | mm |
| TENEO | pupilshift_x | Treatment | mm |
| TENEO | pupilshift_y | Treatment | mm |

| | | | |
|---|---|---|---|
| TENEO | totalpulse | Treatment | Integer |
| TENEO | duration | Treatment | Second |
| TENEO | eyetracker | Eye-tracking | Boolean |
| TENEO | eyetrackeraxis | Eye-tracking | Degree |
| TENEO | eyetrackeraxismean | Eye-tracking | Degree |
| TENEO | eyetrackeraxisrangemin | Eye-tracking | Degree |
| TENEO | eyetrackeraxisrangemax | Eye-tracking | Degree |
| TENEO | eyetrackerreimagepulse | Eye-tracking | Integer |
