## Supplemental Table 2 for "Machine Learning–based Prediction of LASIK Console Inputs for Aspheric Planning (Q-factor, Defocus, Astigmatism): A Translational Methods Study": SUP TABLE 2.pdf

| Device | Features | Category | Unit |
|---|---|---|---|
| ZYWAVE | measurementtime | Time | Unix datetime |
| ZYWAVE | side | ID | OS/OD |
| ZYWAVE | normalband_diameter | Aberrometry Exam | mm |
| ZYWAVE | Z331 | Aberrometry | µm RMS |
| ZYWAVE | Z311 | Aberrometry | µm RMS |
| ZYWAVE | Z310 | Aberrometry | µm RMS |
| ZYWAVE | Z330 | Aberrometry | µm RMS |
| ZYWAVE | Z441 | Aberrometry | µm RMS |
| ZYWAVE | Z421 | Aberrometry | µm RMS |
| ZYWAVE | Z400 | Aberrometry | µm RMS |
| ZYWAVE | Z420 | Aberrometry | µm RMS |
| ZYWAVE | Z440 | Aberrometry | µm RMS |
| ZYWAVE | Z551 | Aberrometry | µm RMS |
| ZYWAVE | Z531 | Aberrometry | µm RMS |
| ZYWAVE | Z511 | Aberrometry | µm RMS |
| ZYWAVE | Z510 | Aberrometry | µm RMS |
| ZYWAVE | Z530 | Aberrometry | µm RMS |
| ZYWAVE | Z550 | Aberrometry | µm RMS |
| ZYWAVE | wavefront_diameter | Aberrometry | mm |
| ZYWAVE | examination_diameter | Aberrometry | mm |
| ZYWAVE | pupil_dimension | Aberrometry | mm |
| ZYWAVE | pupil_dark | Aberrometry | mm |
| ZYWAVE | subjectiverefraction_sphere | Refraction | Diopter (D) |
| ZYWAVE | subjectiverefraction_cylinder | Refraction | Diopter (D) |
| ZYWAVE | subjectiverefraction_axis | Refraction | Axis (°) |
| ZYWAVE | PPR_sphere | Refraction | Diopter (D) |
| ZYWAVE | PPR_cylinder | Refraction | Diopter (D) |
| ZYWAVE | PPR_axis | Refraction | Axis (°) |
| ZYWAVE | difference_sphere | Refraction | Diopter (D) |
| ZYWAVE | difference_cylinder | Refraction | Diopter (D) |
| ZYWAVE | difference_axis | Refraction | Axis (°) |
| ZYWAVE | PPRexamdiam_sphere | Refraction | Diopter (D) |
| ZYWAVE | PPRexamdiam_cylinder | Refraction | Diopter (D) |
| ZYWAVE | PPRexamdiam_axis | Refraction | Axis (°) |

| | | | |
|---|---|---|---|
| ZYWAVE | wavefront5mm_highorder | Aberrometry | µm RMS |
| ZYWAVE | wavefront5mm_HOwoZ400 | Aberrometry | µm RMS |
| ZYWAVE | wavefront5mm_total | Aberrometry | µm RMS |
| ZYWAVE | wavefront6mm_highorder | Aberrometry | µm RMS |
| ZYWAVE | wavefront6mm_HOwoZ400 | Aberrometry | µm RMS |
| ZYWAVE | wavefront6mm_total | Aberrometry | µm RMS |
