## Supplemental Table 3 for "Machine Learning–based Prediction of LASIK Console Inputs for Aspheric Planning (Q-factor, Defocus, Astigmatism): A Translational Methods Study": SUP TABLE 3.pdf

| Device | Features | Category | Unit |
|---|---|---|---|
| ORBSCAN | measurementtime | Time | Unix datetime |
| ORBSCAN | side | ID | OS/OD |
| ORBSCAN | simkastigmatism_value | Simulated Ks | Diopter (D) |
| ORBSCAN | simkastigmatism_axis | Simulated Ks | Axis (°) |
| ORBSCAN | simkmaximum_value | Simulated Ks | Diopter (D) |
| ORBSCAN | simkamaximum_axis | Simulated Ks | Axis (°) |
| ORBSCAN | simkaminimum_value | Simulated Ks | Diopter (D) |
| ORBSCAN | simkminimum_axis | Simulated Ks | Axis (°) |
| ORBSCAN | 3mm_irregularity_std | 3mm zone | Diopter (D) |
| ORBSCAN | 3mm_meanpower | 3mm zone | Diopter (D) |
| ORBSCAN | 3mm_meanpower_std | 3mm zone | Diopter (D) |
| ORBSCAN | 3mm_astigpower | 3mm zone | Diopter (D) |
| ORBSCAN | 3mm_astigpower_std | 3mm zone | Diopter (D) |
| ORBSCAN | 3mm_steepaxis | 3mm zone | Axis (°) |
| ORBSCAN | 3mm_steepaxis_std | 3mm zone | Axis (°) |
| ORBSCAN | 3mm_flataxis | 3mm zone | Axis (°) |
| ORBSCAN | 3mm_flataxis_std | 3mm zone | Axis (°) |
| ORBSCAN | 5mm_irregularity_std | 5mm zone | Diopter (D) |
| ORBSCAN | 5mm_meanpower | 5mm zone | Diopter (D) |
| ORBSCAN | 5mm_meanpower_std | 5mm zone | Diopter (D) |
| ORBSCAN | 5mm_astigpower | 5mm zone | Diopter (D) |
| ORBSCAN | 5mm_astigpower_std | 5mm zone | Diopter (D) |
| ORBSCAN | 5mm_steepaxis | 5mm zone | Axis (°) |
| ORBSCAN | 5mm_steepaxis_std | 5mm zone | Axis (°) |
| ORBSCAN | 5mm_flataxis | 5mm zone | Axis (°) |
| ORBSCAN | 5mm_flataxis_std | 5mm zone | Axis (°) |
| ORBSCAN | thinnestpoint_pachymetry | Topography | μm |
| ORBSCAN | thinnestpoint_x | Topography | mm |
| ORBSCAN | thinnestpoint_y | Topography | mm |
| ORBSCAN | kappa_module | Topography | Degree (°) |
| ORBSCAN | kappa_argument | Topography | Axis (°) |
| ORBSCAN | kappa_x | Topography | mm |
| ORBSCAN | kappa_y | Topography | mm |
| ORBSCAN | acousticfactor | Topography | No unit |
| ORBSCAN | anteriorchamberdepth | Topography | mm |

| | | | |
|---|---|---|---|
| ORBSCAN | whitetowhite | Topography | mm |
| ORBSCAN | pupildiameter | Topography | mm |
| ORBSCAN | bfsanteriormap_centralpower | Best Fit Values | Diopter (D) |
| ORBSCAN | bfsanteriormap_centralradius | Best Fit Values | mm |
| ORBSCAN | bfsanteriormap_centralastigvalue | Best Fit Values | Diopter (D) |
| ORBSCAN | bfsanteriormap_centralastigaxis | Best Fit Values | Axis (°) |
| ORBSCAN | bfsanteriormap_asphericity | Best Fit Values | No unit |
| ORBSCAN | bfsanteriormap_eccentricity | Best Fit Values | No unit |
| ORBSCAN | bfsanteriormap_shapefactor | Best Fit Values | No unit |
| ORBSCAN | bfsanteriormap_bfsmm | Best Fit Values | mm |
| ORBSCAN | bfsanteriormap_bfsdiopter | Best Fit Values | Diopter (D) |
| ORBSCAN | bfsanteriormap_apicalpower | Best Fit Values | Diopter (D) |
| ORBSCAN | bfsanteriormap_apicalradius | Best Fit Values | mm |
| ORBSCAN | bfsanteriormap_maximumK | Best Fit Values | Diopter (D) |
| ORBSCAN | bfsanteriormap_minimumK | Best Fit Values | Diopter (D) |
| ORBSCAN | bfsanteriormap_apicalastigvalue | Best Fit Values | Diopter (D) |
| ORBSCAN | bfsanteriormap_apicalastigaxis | Best Fit Values | Axis (°) |
| ORBSCAN | bfsposteriormap_centralpower | Best Fit Values | Diopter (D) |
| ORBSCAN | bfsposteriormap_centralradius | Best Fit Values | mm |
| ORBSCAN | bfsposteriormap_centralastigvalue | Best Fit Values | Diopter (D) |
| ORBSCAN | bfsposteriormap_centralastigaxis | Best Fit Values | Axis (°) |
| ORBSCAN | bfsposteriormap_asphericity | Best Fit Values | No unit |
| ORBSCAN | bfsposteriormap_eccentricity | Best Fit Values | No unit |
| ORBSCAN | bfsposteriormap_shapefactor | Best Fit Values | No unit |
| ORBSCAN | bfsposteriormap_bfsmm | Best Fit Values | mm |
| ORBSCAN | bfsposteriormap_bfsdiopter | Best Fit Values | Diopter (D) |
| ORBSCAN | bfsposteriormap_apicalpower | Best Fit Values | Diopter (D) |
| ORBSCAN | bfsposteriormap_apicalradius | Best Fit Values | mm |
| ORBSCAN | bfsposteriormap_maximumK | Best Fit Values | Diopter (D) |
| ORBSCAN | bfsposteriormap_minimumK | Best Fit Values | Diopter (D) |
| ORBSCAN | bfsposteriormap_apicalastigvalue | Best Fit Values | Diopter (D) |
| ORBSCAN | bfsposteriormap_apicalastigaxis | Best Fit Values | Axis (°) |
