## Supplemental Notes 1 for "Machine Learning–based Prediction of LASIK Console Inputs for Aspheric Planning (Q-factor, Defocus, Astigmatism): A Translational Methods Study"

### LINR — Natural-scale coefficients for outcome Defocus ($yZ_2^0$)

| **Predictor** | **Defocus ($yZ_2^0$) (β)** |
|---|---:|
| Intercept | 0.429 |
| Pre-op Defocus ($Z_2^0$) | 0.867 |
| Pre-op Astigm+ ($Z_2^{-2}$) | -0.017 |
| Pre-op Astigmx ($Z_2^{+2}$) | 0.013 |
| Pre-op Asphericity ($Z_4^0$) | -0.019 |
| Post-op Defocus ($Z_2^0$) | -0.580 |
| Post-op Astigm+ ($Z_2^{-2}$) | 0.087 |
| Post-op Astigmx ($Z_2^{+2}$) | -0.008 |
| Post-op Asphericity ($Z_4^0$) | 0.099 |
| Optical Zone (Defocus) | 0.210 |
| Optical Zone (Q-factor) | -0.327 |
| Pre-op Km | 0.009 |
| Pre-op Q | -0.045 |

Coefficients on the original scale. Pipeline: StandardScaler → MultiOutputRegressor(LinearRegression). Transform: β_nat = β_std/σ; Intercept_nat = Intercept_std − Σ(β_std·μ/σ).

### LINR — Natural-scale coefficients for outcome Astigm+ ($yZ_2^{-2}$)

| **Predictor** | **Astigm+ ($yZ_2^{-2}$) (β)** |
|---|---|
| Intercept | -0.392 |
| Pre-op Defocus ($Z_2^0$) | 0.010 |
| Pre-op Astigm+ ($Z_2^{-2}$) | 0.933 |
| Pre-op Astigmx ($Z_2^{+2}$) | 0.039 |
| Pre-op Asphericity ($Z_4^0$) | -0.280 |
| Post-op Defocus ($Z_2^0$) | -0.002 |
| Post-op Astigm+ ($Z_2^{-2}$) | -0.270 |
| Post-op Astigmx ($Z_2^{+2}$) | -0.001 |
| Post-op Asphericity ($Z_4^0$) | 0.093 |
| Optical Zone (Defocus) | 0.010 |
| Optical Zone (Q-factor) | -0.004 |
| Pre-op Km | 0.006 |
| Pre-op Q | 0.066 |

Coefficients on the original scale. Pipeline: StandardScaler → MultiOutputRegressor(LinearRegression). Transform: β_nat = β_std/σ; Intercept_nat = Intercept_std − Σ(β_std·μ/σ).

### LINR — Natural-scale coefficients for outcome Astigmx ($yZ_2^{+2}$)

| **Predictor** | **Astigmx ($yZ_2^{+2}$) (β)** |
|---|---|
| Intercept | 0.025 |
| Pre-op Defocus ($Z_2^0$) | 0.003 |
| Pre-op Astigm+ ($Z_2^{-2}$) | -0.055 |
| Pre-op Astigmx ($Z_2^{+2}$) | 0.834 |
| Pre-op Asphericity ($Z_4^0$) | 0.013 |
| Post-op Defocus ($Z_2^0$) | -0.008 |
| Post-op Astigm+ ($Z_2^{-2}$) | -0.013 |
| Post-op Astigmx ($Z_2^{+2}$) | -0.251 |
| Post-op Asphericity ($Z_4^0$) | -0.009 |
| Optical Zone (Defocus) | -0.005 |
| Optical Zone (Q-factor) | -0.011 |
| Pre-op Km | 0.002 |
| Pre-op Q | -0.021 |

Coefficients on the original scale. Pipeline: StandardScaler → MultiOutputRegressor(LinearRegression). Transform: β_nat = β_std/σ; Intercept_nat = Intercept_std − Σ(β_std·μ/σ).

### LINR — Natural-scale coefficients for outcome Q-factor ($yZ_4^0$)

| **Predictor** | **Q-factor ($yZ_4^0$) (β)** |
|---|---:|
| Intercept | 0.630 |
| Pre-op Defocus ($Z_2^0$) | -0.258 |
| Pre-op Astigm+ ($Z_2^{-2}$) | -0.002 |
| Pre-op Astigmx ($Z_2^{+2}$) | -0.003 |
| Pre-op Asphericity ($Z_4^0$) | 3.527 |
| Post-op Defocus ($Z_2^0$) | 0.069 |
| Post-op Astigm+ ($Z_2^{-2}$) | 0.123 |
| Post-op Astigmx ($Z_2^{+2}$) | 0.015 |
| Post-op Asphericity ($Z_4^0$) | -2.326 |
| Optical Zone (Defocus) | 0.197 |
| Optical Zone (Q-factor) | -0.041 |
| Pre-op Km | -0.031 |
| Pre-op Q | -0.056 |

Coefficients on the original scale. Pipeline: StandardScaler → MultiOutputRegressor(LinearRegression). Transform: β_nat = β_std/σ; Intercept_nat = Intercept_std − Σ(β_std·μ/σ).

### LINR — Partial $R^2$ per predictor — Target: Defocus ($yZ_2^0$)

| Predictor | Partial $R^2$ | $\Delta R^2$ | F(1,df2) | p-value |
|---|---:|---:|---:|---:|
| Pre-op Defocus ($Z_2^0$ | 0.9566 | 0.4482 | 5.368e+04 | 0 |
| Post-op Defocus ($Z$ | 0.5283 | 0.02277 | 2727 | 0 |
| Optical Zone (Q-fa | 0.07212 | 0.001581 | 189.3 | 1.573e-41 |
| Optical Zone (Defo | 0.0233 | 0.000485 | 58.08 | 3.58e-14 |
| Post-op Astigm+ ( | 0.008778 | 0.0001801 | 21.56 | 3.603e-06 |
| Post-op Asphericity | 0.001784 | 3.635e-05 | 4.353 | 0.03705 |
| Pre-op Astigm+ ($Z$ | 0.001595 | 3.247e-05 | 3.889 | 0.04872 |
| Pre-op Km | 0.001232 | 2.508e-05 | 3.003 | 0.08322 |
| Pre-op Q | 0.000555 | 1.129e-05 | 1.352 | 0.245 |
| Pre-op Astigmx ($Z_2$ | 0.0003333 | 6.78e-06 | 0.8119 | 0.3676 |
| Post-op Astigmx ($Z$ | 4.429e-05 | 9.006e-07 | 0.1078 | 0.7426 |
| Pre-op Asphericity | 3.594e-05 | 7.308e-07 | 0.08751 | 0.7674 |

Nested OLS on scaled X. For predictor x_j, reduced model drops x_j.
Partial $R^2$ = (SSE_reduced − SSE_full) / SSE_reduced. $\Delta R^2 = R^2_{full} - R^2_{reduced}$.
F(1,df2) = ((SSE_reduced − SSE_full)/1) / (SSE_full/df2), with df2 = n − p − 1.

LINR — Partial $R^2$ per predictor — Target: Astigm+ ($yZ_2^{-2}$)

| **Predictor** | **Partial $R^2$** | **$\Delta R^2$** | **F(1,df2)** | **p-value** |
|---|---|---|---|---|
| Pre-op Astigm+ ($Z$ | 0.9187 | 0.844 | 2.751e+04 | 0 |
| Post-op Astigm+ ( | 0.1595 | 0.01418 | 462.1 | 5.315e-94 |
| Pre-op Asphericity | 0.01701 | 0.001293 | 42.15 | 1.022e-10 |
| Pre-op Astigmx ($Z_2$ | 0.006669 | 0.0005016 | 16.35 | 5.437e-05 |
| Pre-op Defocus ($Z_2^0$ | 0.006637 | 0.0004992 | 16.27 | 5.664e-05 |
| Post-op Asphericity | 0.003502 | 0.0002626 | 8.558 | 0.003472 |
| Pre-op Q | 0.002689 | 0.0002015 | 6.566 | 0.01045 |
| Pre-op Km | 0.001462 | 0.0001094 | 3.564 | 0.05915 |
| Optical Zone (Defo | 0.0001306 | 9.761e-06 | 0.3181 | 0.5728 |
| Optical Zone (Q-fa | 2.378e-05 | 1.777e-06 | 0.05791 | 0.8098 |
| Post-op Defocus ($Z$ | 1.983e-05 | 1.482e-06 | 0.04829 | 0.8261 |
| Post-op Astigmx ($Z$ | 8.927e-07 | 6.67e-08 | 0.002174 | 0.9628 |

Nested OLS on scaled X. For predictor x_j, reduced model drops x_j.
Partial $R^2$ = (SSE_reduced − SSE_full) / SSE_reduced. $\Delta R^2 = R^2_{full} - R^2_{reduced}$.
F(1,df2) = ((SSE_reduced − SSE_full)/1) / (SSE_full/df2), with df2 = n − p − 1.

### LINR — Partial $R^2$ per predictor — Target: Astigmx ($yZ_2^{+2}$)

| **Predictor** | **Partial $R^2$** | **Δ$R^2$** | **F(1,df2)** | **p-value** |
|---|---:|---:|---:|---:|
| Pre-op Astigmx ($Z_2$ | 0.8567 | 0.8089 | 1.456e+04 | 0 |
| Post-op Astigmx ($Z$ | 0.165 | 0.02674 | 481.3 | 1.678e-97 |
| Pre-op Astigm+ ($Z$ | 0.07333 | 0.01071 | 192.7 | 3.217e-42 |
| Pre-op Defocus ($Z_2^0$ | 0.001503 | 0.0002036 | 3.665 | 0.05568 |
| Post-op Defocus ($Z$ | 0.001009 | 0.0001367 | 2.46 | 0.1169 |
| Post-op Astigm+ ( | 0.0008522 | 0.0001154 | 2.077 | 0.1497 |
| Pre-op Q | 0.0005314 | 7.193e-05 | 1.295 | 0.2553 |
| Optical Zone (Q-fa | 0.0004131 | 5.591e-05 | 1.006 | 0.3159 |
| Pre-op Km | 0.0003781 | 5.118e-05 | 0.9211 | 0.3373 |
| Pre-op Asphericity | 7.108e-05 | 9.617e-06 | 0.1731 | 0.6774 |
| Optical Zone (Defo | 6.973e-05 | 9.435e-06 | 0.1698 | 0.6803 |
| Post-op Asphericity | 5.809e-05 | 7.859e-06 | 0.1415 | 0.7069 |

Nested OLS on scaled X. For predictor x_j, reduced model drops x_j.
Partial $R^2$ = (SSE_reduced − SSE_full) / SSE_reduced.  $\Delta R^2 = R^2_{full} - R^2_{reduced}$.
F(1,df2) = ((SSE_reduced − SSE_full)/1) / (SSE_full/df2), with df2 = n − p − 1.

### LINR — Partial $R^2$ per predictor — Target: Q-factor ($yZ_4^0$)

| **Predictor** | **Partial $R^2$** | **$\Delta R^2$** | **F(1,df2)** | **p-value** |
|---|---|---|---|---|
| Pre-op Defocus ($Z_2^0$ | 0.3924 | 0.3366 | 1573 | 8.983e-266 |
| Pre-op Asphericity | 0.2906 | 0.2135 | 997.7 | 8.069e-184 |
| Post-op Asphericity | 0.2459 | 0.17 | 794.1 | 1.869e-151 |
| Optical Zone (Defo | 0.006927 | 0.003635 | 16.99 | 3.893e-05 |
| Post-op Astigm+ (Z | 0.00581 | 0.003046 | 14.23 | 0.0001655 |
| Pre-op Km | 0.005351 | 0.002804 | 13.1 | 0.0003014 |
| Post-op Defocus ($Z$ | 0.005195 | 0.002721 | 12.71 | 0.0003698 |
| Optical Zone (Q-fa | 0.0004026 | 0.0002099 | 0.9808 | 0.3221 |
| Pre-op Q | 0.0002933 | 0.0001529 | 0.7143 | 0.3981 |
| Post-op Astigmx ($Z$ | 5.557e-05 | 2.896e-05 | 0.1353 | 0.713 |
| Pre-op Astigmx ($Z_2$ | 7.422e-06 | 3.868e-06 | 0.01807 | 0.8931 |
| Pre-op Astigm+ ($Z$ | 5.467e-06 | 2.849e-06 | 0.01331 | 0.9082 |

Nested OLS on scaled X. For predictor x_j, reduced model drops x_j.
Partial $R^2$ = (SSE_reduced − SSE_full) / SSE_reduced. $\Delta R^2 = R^2_{full} - R^2_{reduced}$.
F(1,df2) = ((SSE_reduced − SSE_full)/1) / (SSE_full/df2), with df2 = n − p − 1.

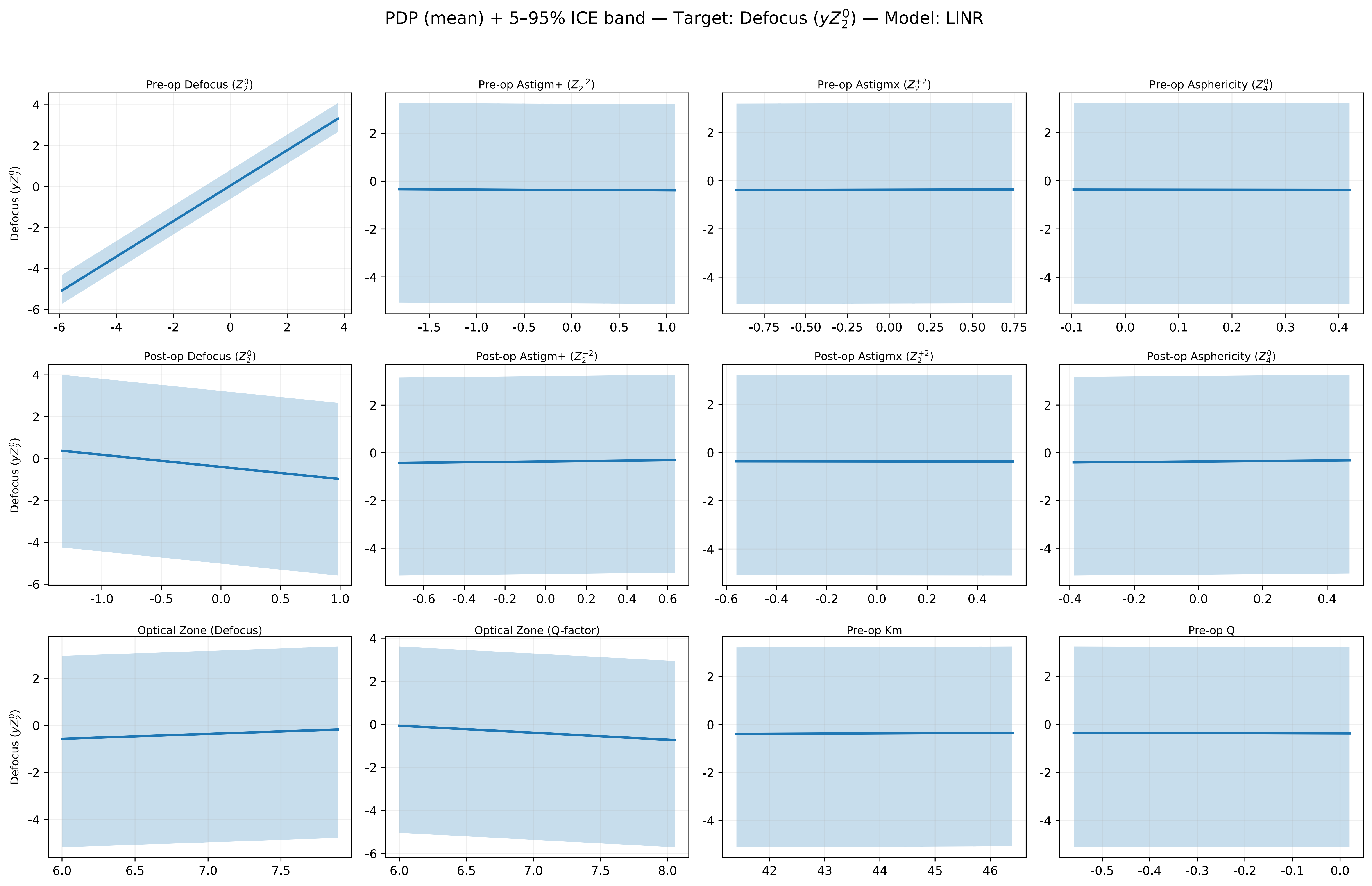

PDP (mean) + 5–95% ICE band — Target: Defocus ($yZ_2^0$) — Model: LINR
Pre-op Defocus ($Z_2^0$)
Pre-op Astigm+ ($Z_2^{-2}$)
Pre-op Astigmx ($Z_2^{+2}$)
Pre-op Asphericity ($Z_4^0$)
Post-op Defocus ($Z_2^0$)
Post-op Astigm+ ($Z_2^{-2}$)
Post-op Astigmx ($Z_2^{+2}$)
Post-op Asphericity ($Z_4^0$)
Optical Zone (Defocus)
Optical Zone (Q-factor)
Pre-op Km
Pre-op Q
Defocus ($yZ_2^0$)

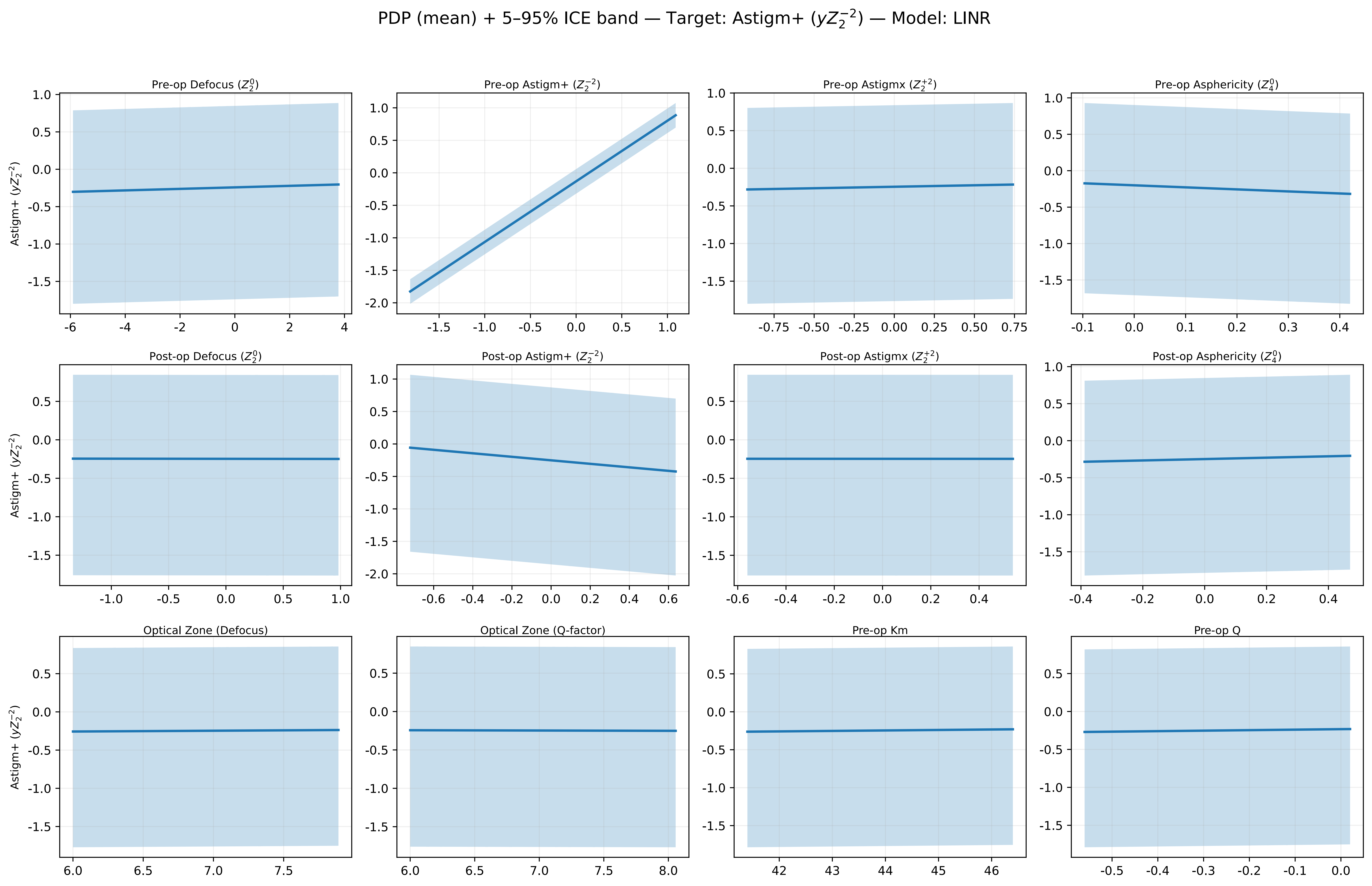

PDP (mean) + 5–95% ICE band — Target: Astigm+ ($yZ_2^{-2}$) — Model: LINR
Pre-op Defocus ($Z_2^0$)
Pre-op Astigm+ ($Z_2^{-2}$)
Pre-op Astigmx ($Z_2^{+2}$)
Pre-op Asphericity ($Z_4^0$)
Post-op Defocus ($Z_2^0$)
Post-op Astigm+ ($Z_2^{-2}$)
Post-op Astigmx ($Z_2^{+2}$)
Post-op Asphericity ($Z_4^0$)
Optical Zone (Defocus)
Optical Zone (Q-factor)
Pre-op Km
Pre-op Q
Astigm+ ($yZ_2^{-2}$)

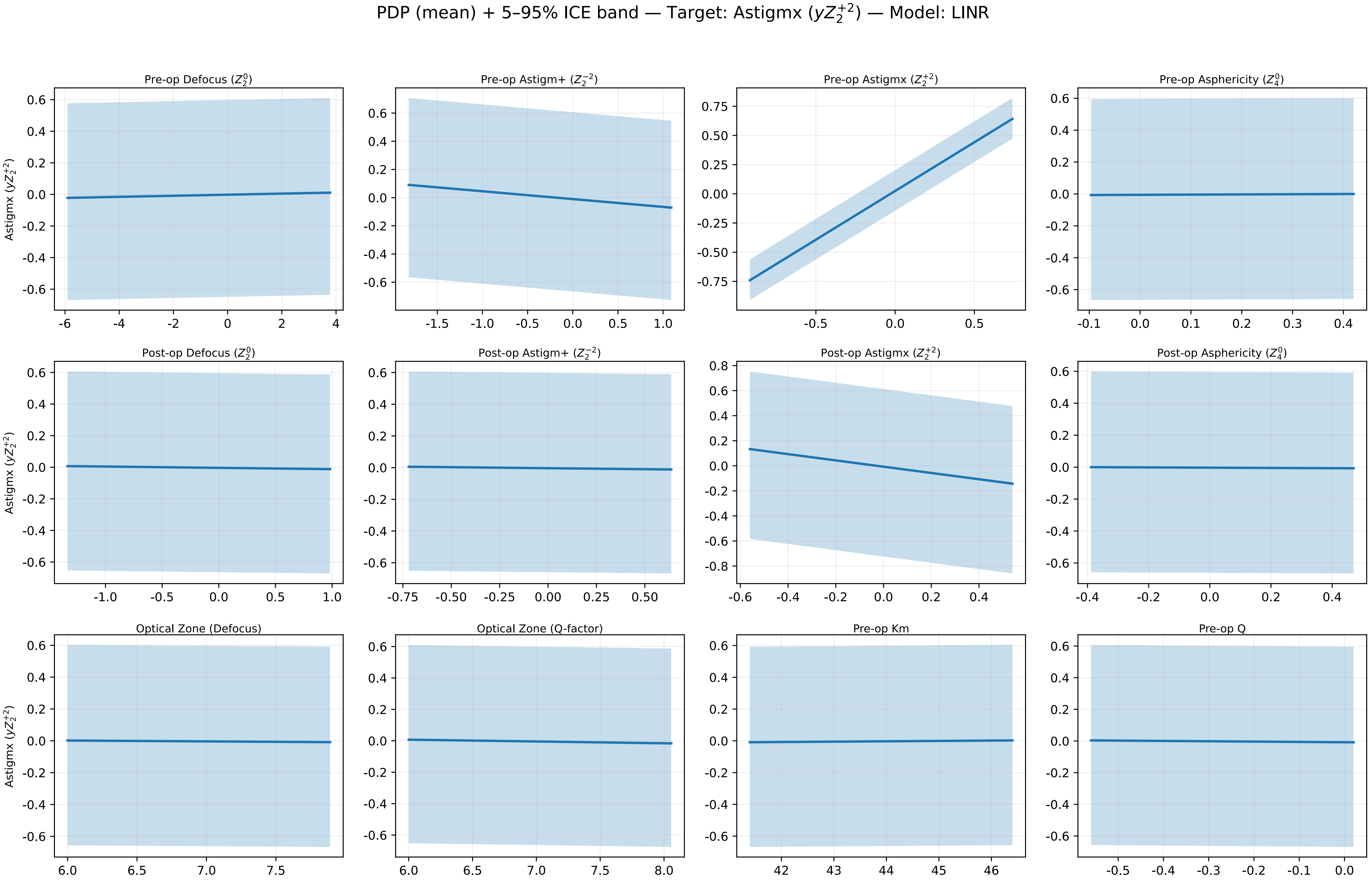
PDP (mean) + 5–95% ICE band — Target: Astigmx ($yZ_2^{+2}$) — Model: LINR
Pre-op Defocus ($Z_2^0$)
Pre-op Astigm+ ($Z_2^{-2}$)
Pre-op Astigmx ($Z_2^{+2}$)
Pre-op Asphericity ($Z_4^0$)
Post-op Defocus ($Z_2^0$)
Post-op Astigm+ ($Z_2^{-2}$)
Post-op Astigmx ($Z_2^{+2}$)
Post-op Asphericity ($Z_4^0$)
Optical Zone (Defocus)
Optical Zone (Q-factor)
Pre-op Km
Pre-op Q
Astigmx ($yZ_2^{+2}$)

PDP (mean) + 5–95% ICE band — Target: Q-factor ($yZ_4^0$) — Model: LINR

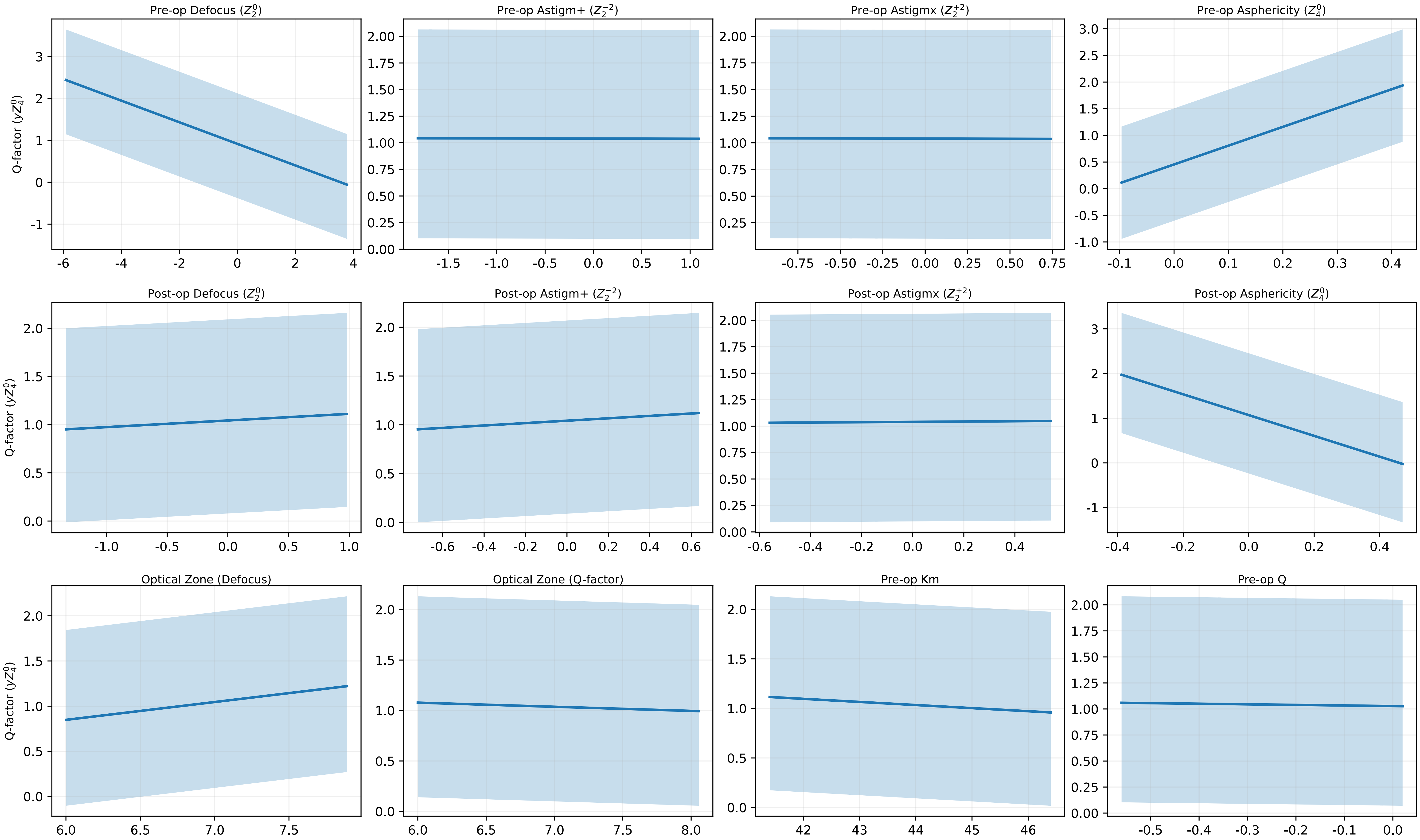

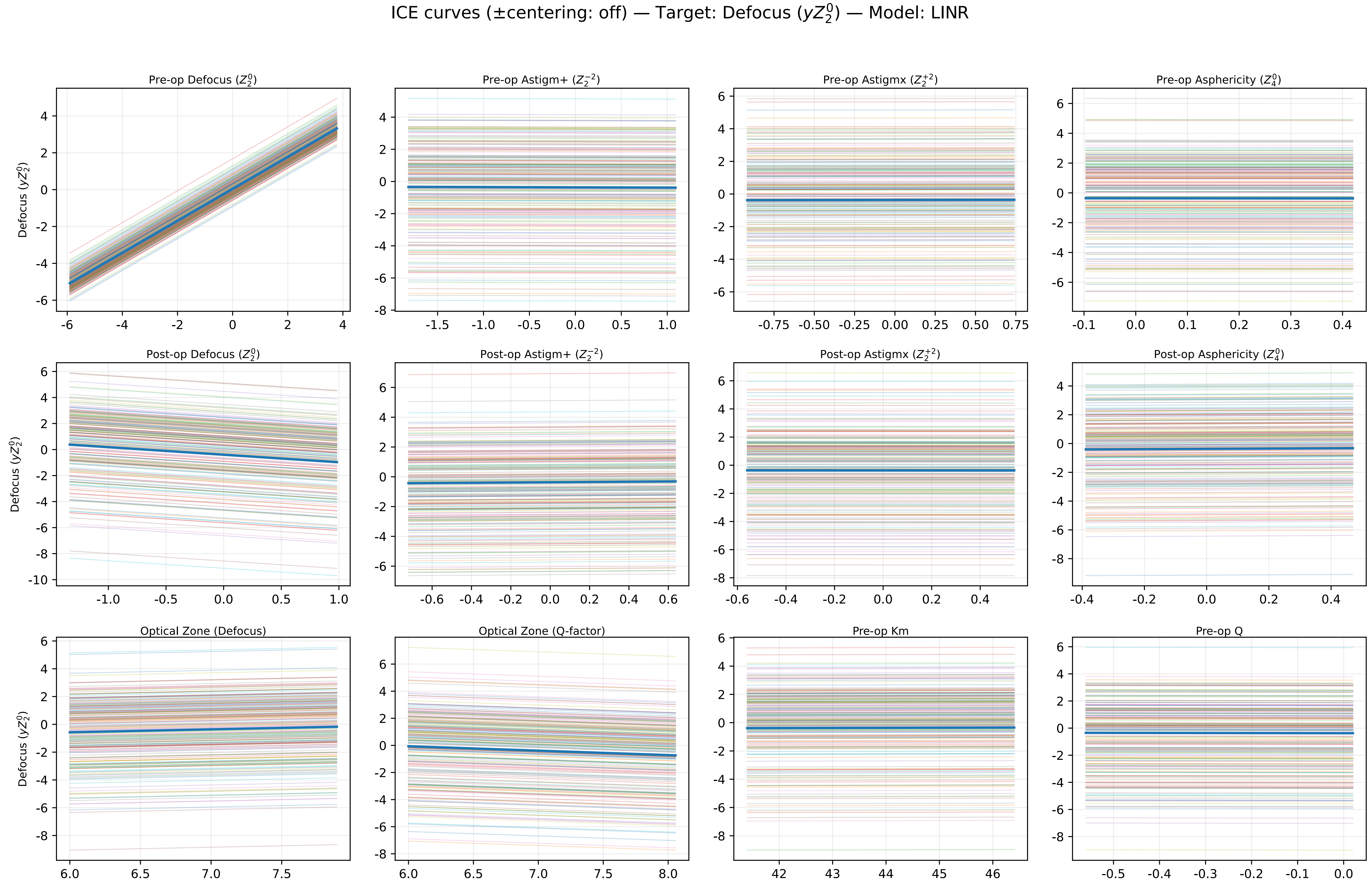

ICE curves (±centering: off) — Target: Defocus ($yZ_2^0$) — Model: LINR
Pre-op Defocus ($Z_2^0$)
Pre-op Astigm+ ($Z_2^{-2}$)
Pre-op Astigmx ($Z_2^{+2}$)
Pre-op Asphericity ($Z_4^0$)
Post-op Defocus ($Z_2^0$)
Post-op Astigm+ ($Z_2^{-2}$)
Post-op Astigmx ($Z_2^{+2}$)
Post-op Asphericity ($Z_4^0$)
Optical Zone (Defocus)
Optical Zone (Q-factor)
Pre-op Km
Pre-op Q
Defocus ($yZ_2^0$)

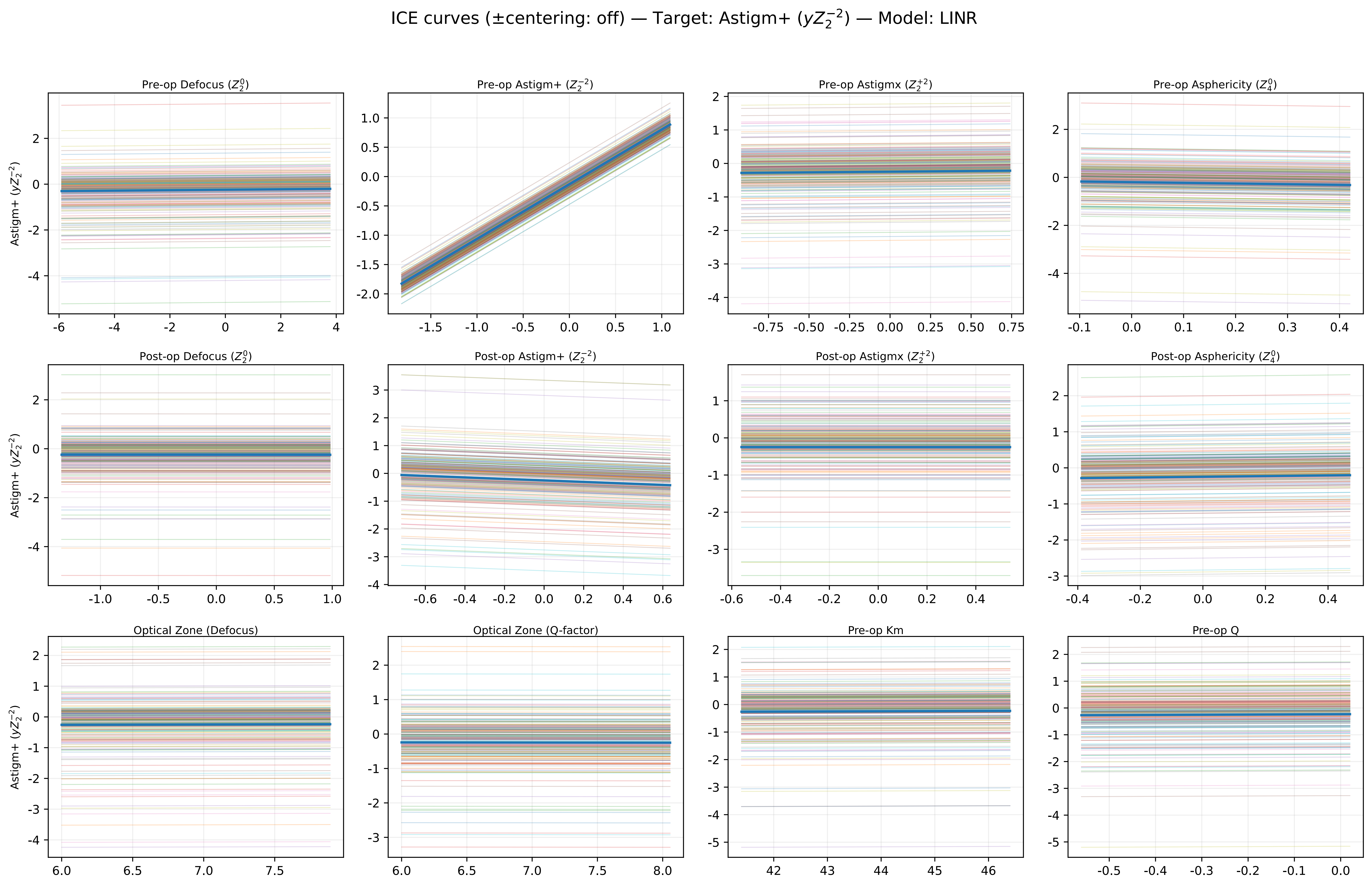

ICE curves (±centering: off) — Target: Astigm+ ($yZ_2^{-2}$) — Model: LINR
Pre-op Defocus ($Z_2^0$)
Pre-op Astigm+ ($Z_2^{-2}$)
Pre-op Astigmx ($Z_2^{+2}$)
Pre-op Asphericity ($Z_4^0$)
Post-op Defocus ($Z_2^0$)
Post-op Astigm+ ($Z_2^{-2}$)
Post-op Astigmx ($Z_2^{+2}$)
Post-op Asphericity ($Z_4^0$)
Optical Zone (Defocus)
Optical Zone (Q-factor)
Pre-op Km
Pre-op Q
Astigm+ ($yZ_2^{-2}$)

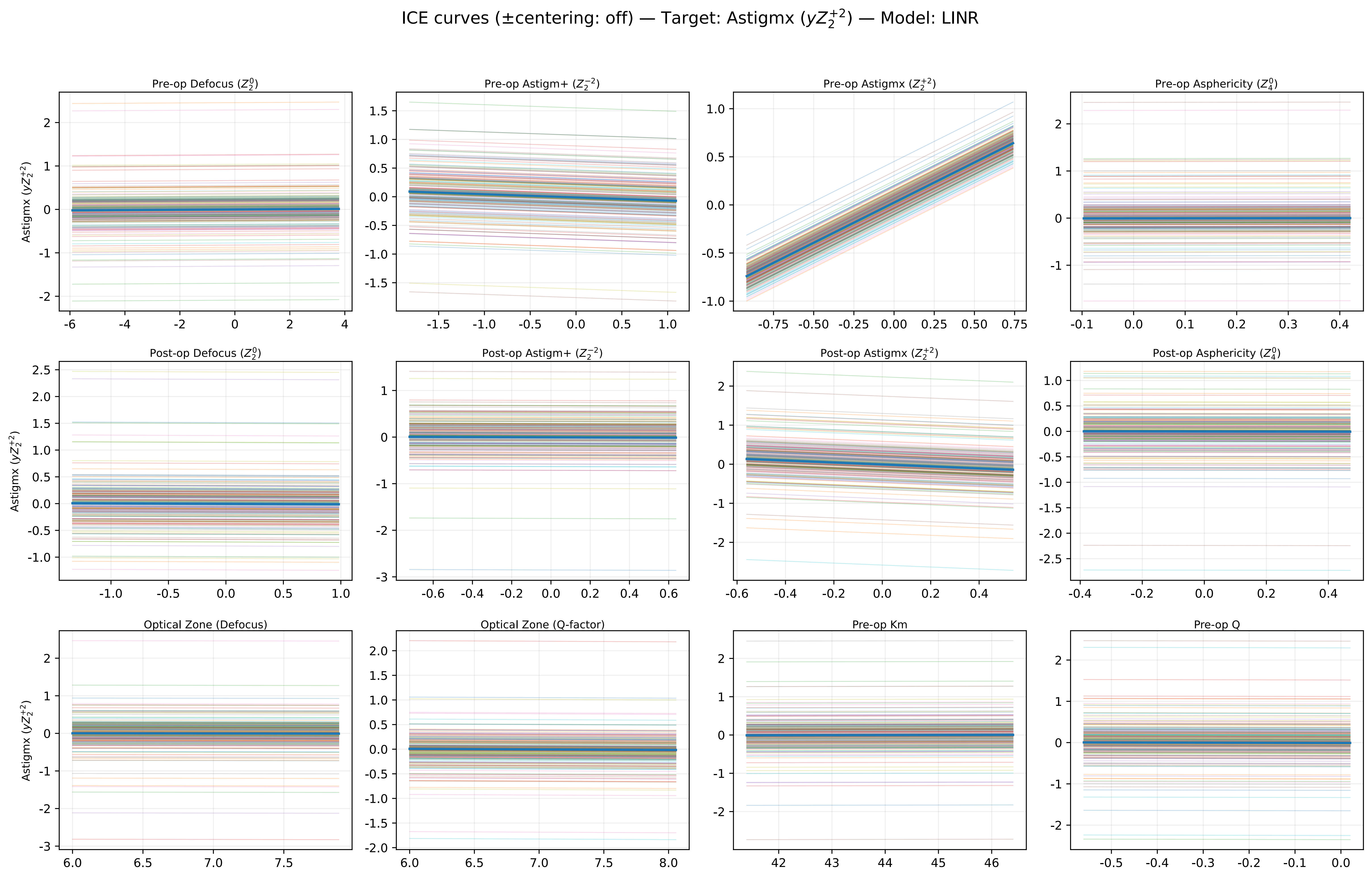

ICE curves (±centering: off) — Target: Astigmx ($yZ_2^{+2}$) — Model: LINR
Pre-op Defocus ($Z_2^0$)
Pre-op Astigm+ ($Z_2^{-2}$)
Pre-op Astigmx ($Z_2^{+2}$)
Pre-op Asphericity ($Z_4^0$)
Post-op Defocus ($Z_2^0$)
Post-op Astigm+ ($Z_2^{-2}$)
Post-op Astigmx ($Z_2^{+2}$)
Post-op Asphericity ($Z_4^0$)
Optical Zone (Defocus)
Optical Zone (Q-factor)
Pre-op Km
Pre-op Q
Astigmx ($yZ_2^{+2}$)

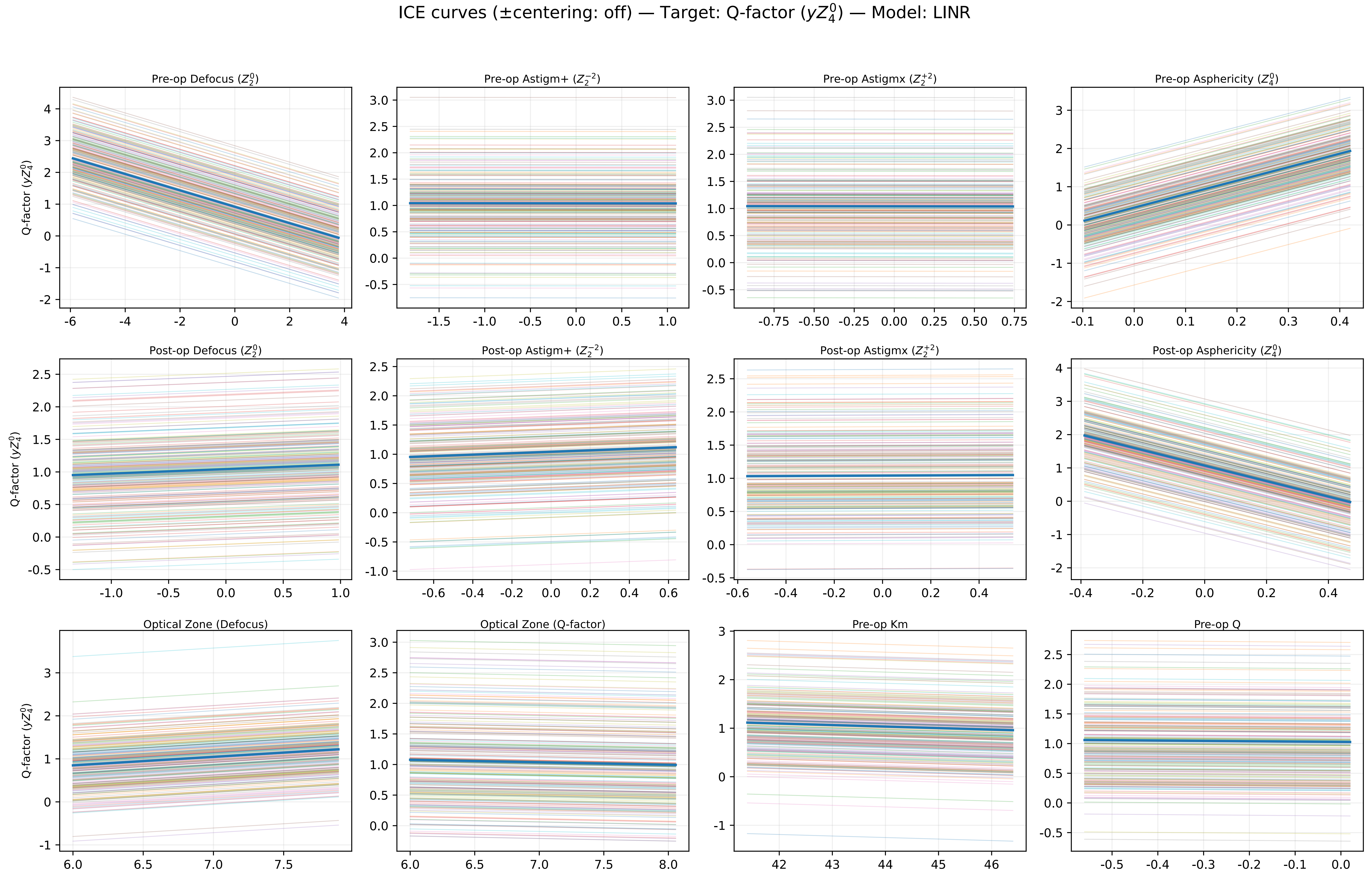
ICE curves (±centering: off) — Target: Q-factor ($yZ_4^0$) — Model: LINR
Pre-op Defocus ($Z_2^0$)
Pre-op Astigm+ ($Z_2^{-2}$)
Pre-op Astigmx ($Z_2^{+2}$)
Pre-op Asphericity ($Z_4^0$)
Post-op Defocus ($Z_2^0$)
Post-op Astigm+ ($Z_2^{-2}$)
Post-op Astigmx ($Z_2^{+2}$)
Post-op Asphericity ($Z_4^0$)
Optical Zone (Defocus)
Optical Zone (Q-factor)
Pre-op Km
Pre-op Q
Q-factor ($yZ_4^0$)

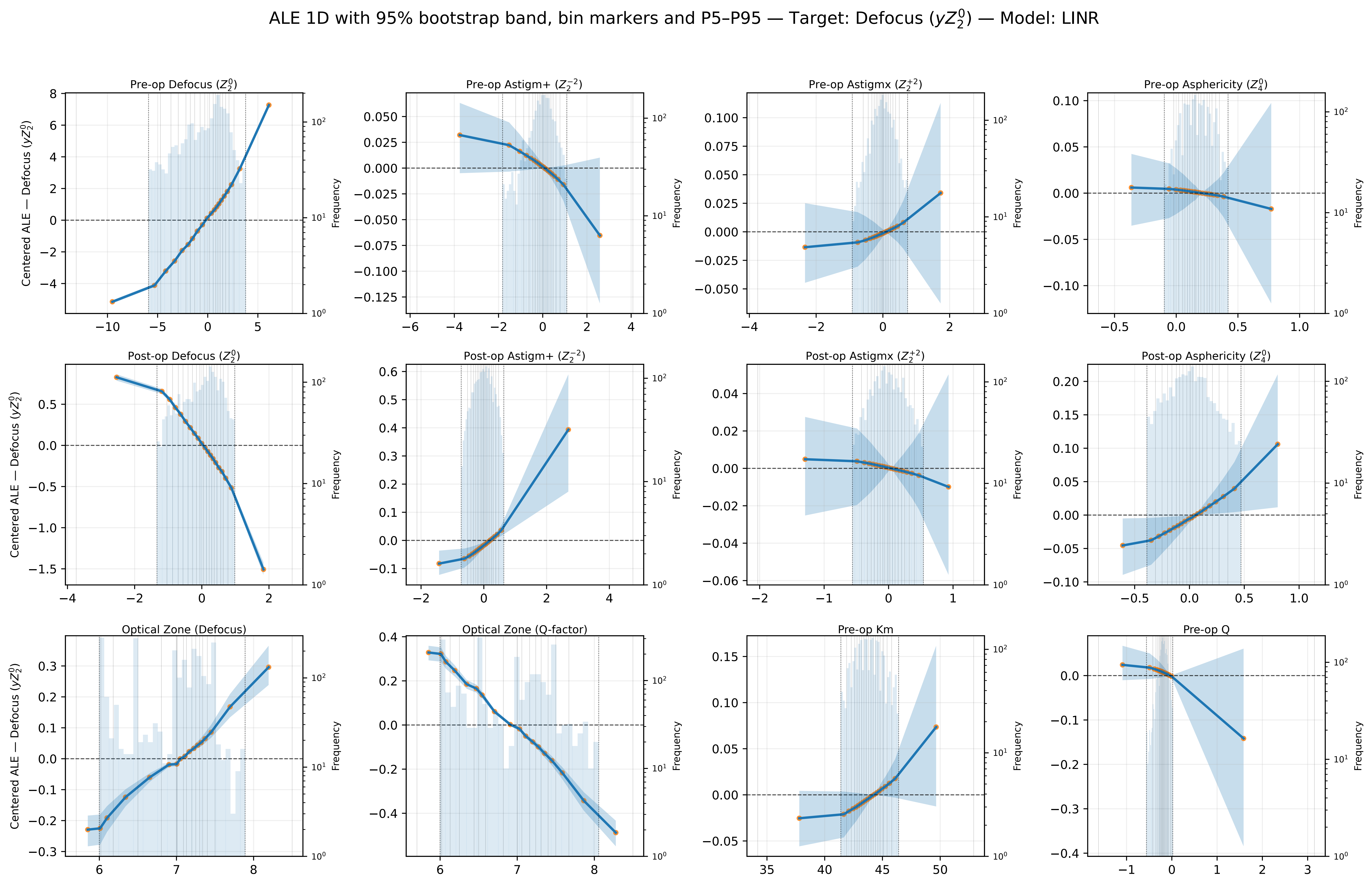

ALE 1D with 95% bootstrap band, bin markers and P5–P95 — Target: Defocus ($yZ_2^0$) — Model: LINR
Pre-op Defocus ($Z_2^0$)
Pre-op Astigm+ ($Z_2^{-2}$)
Pre-op Astigmx ($Z_2^{+2}$)
Pre-op Asphericity ($Z_4^0$)
Post-op Defocus ($Z_2^0$)
Post-op Astigm+ ($Z_2^{-2}$)
Post-op Astigmx ($Z_2^{+2}$)
Post-op Asphericity ($Z_4^0$)
Optical Zone (Defocus)
Optical Zone (Q-factor)
Pre-op Km
Pre-op Q
Centered ALE — Defocus ($yZ_2^0$)
Frequency

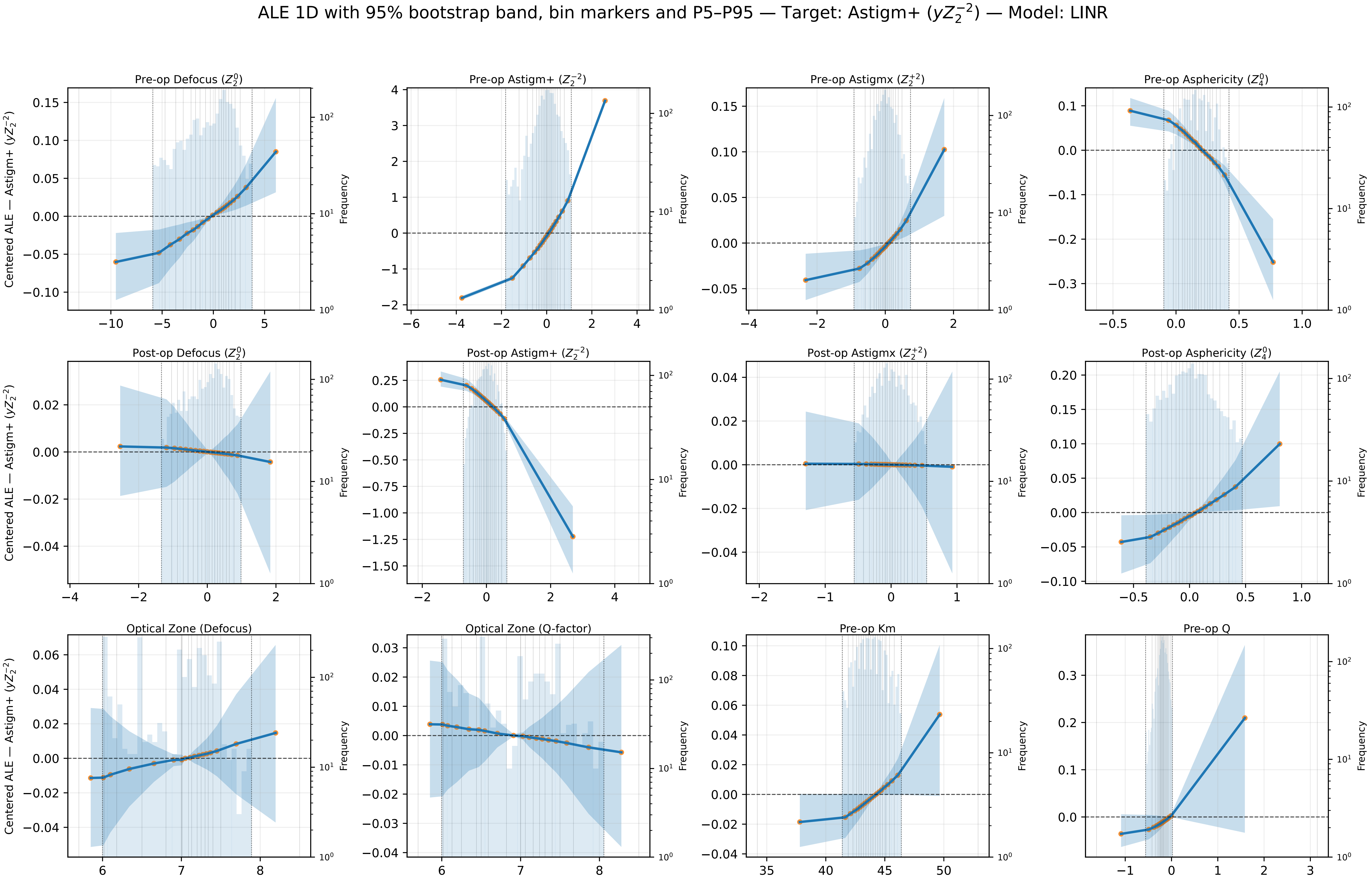
ALE 1D with 95% bootstrap band, bin markers and P5–P95 — Target: Astigm+ ($yZ_2^{-2}$) — Model: LINR
Pre-op Defocus ($Z_2^0$)
Pre-op Astigm+ ($Z_2^{-2}$)
Pre-op Astigmx ($Z_2^{+2}$)
Pre-op Asphericity ($Z_4^0$)
Post-op Defocus ($Z_2^0$)
Post-op Astigm+ ($Z_2^{-2}$)
Post-op Astigmx ($Z_2^{+2}$)
Post-op Asphericity ($Z_4^0$)
Optical Zone (Defocus)
Optical Zone (Q-factor)
Pre-op Km
Pre-op Q
Centered ALE — Astigm+ ($yZ_2^{-2}$)
Frequency

ALE 1D with 95% bootstrap band, bin markers and P5–P95 — Target: Astigmx ($yZ_2^{+2}$) — Model: LINR

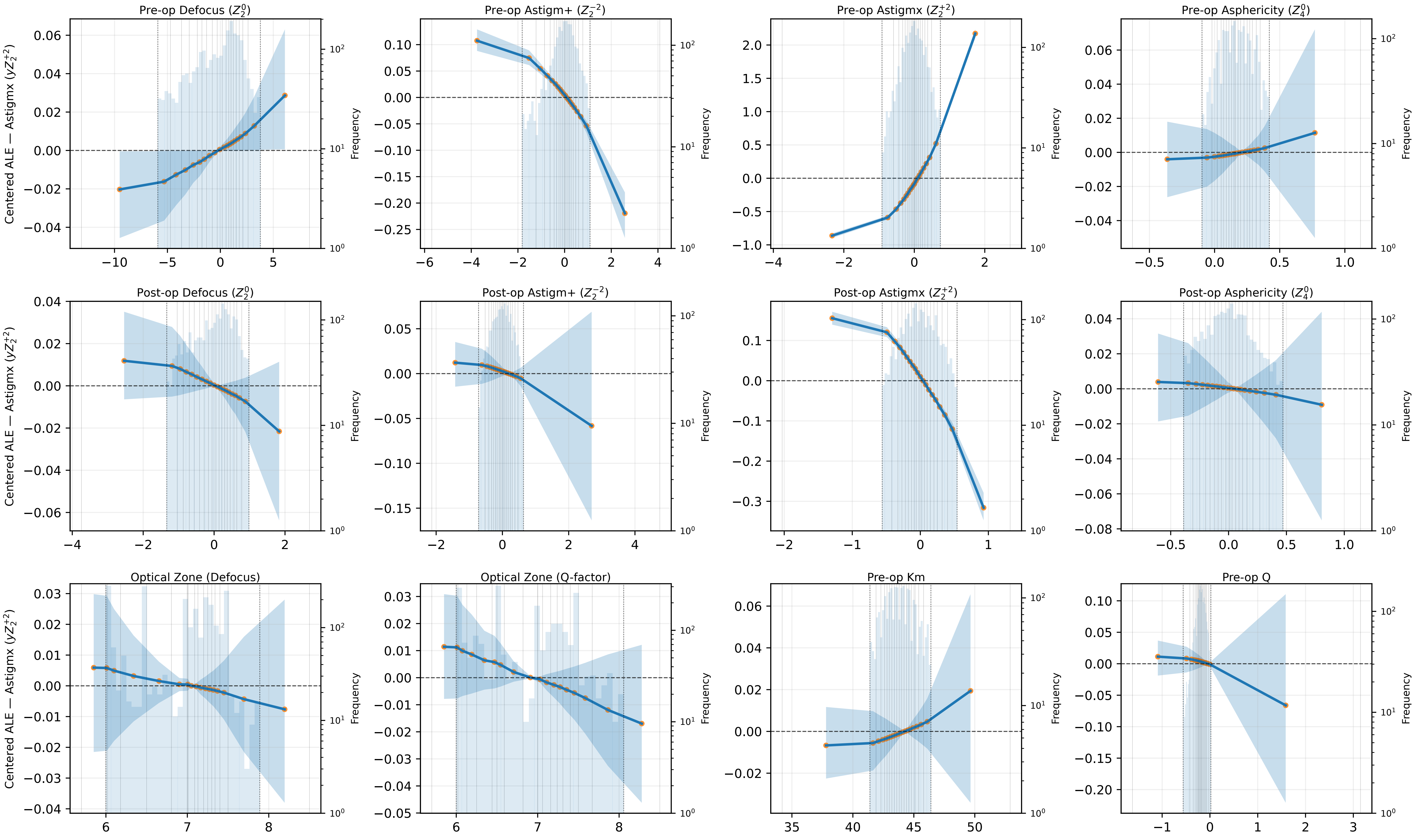

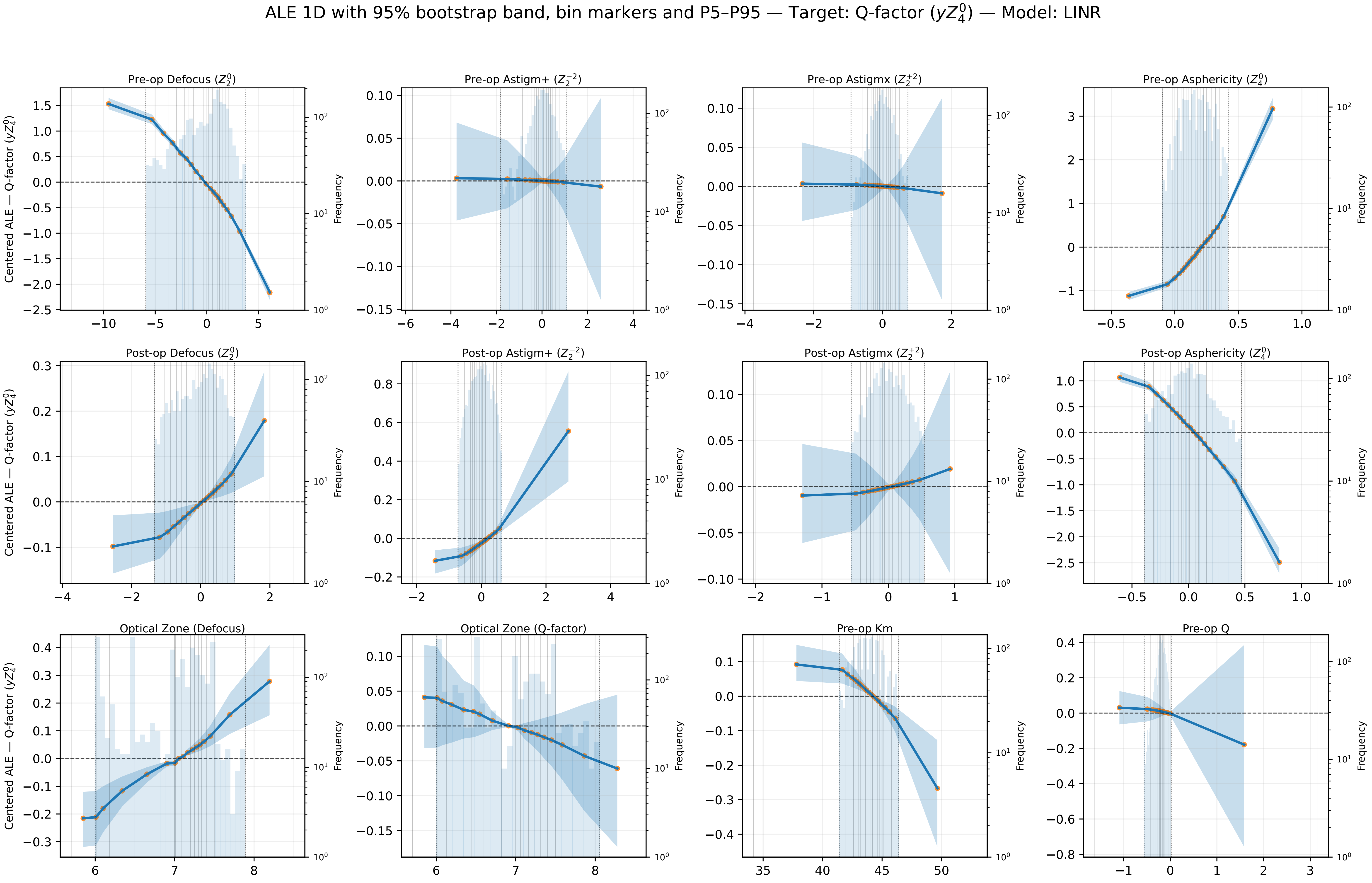

ALE 1D with 95% bootstrap band, bin markers and P5–P95 — Target: Q-factor ($yZ_4^0$) — Model: LINR
Pre-op Defocus ($Z_2^0$)
Pre-op Astigm+ ($Z_2^{-2}$)
Pre-op Astigmx ($Z_2^{+2}$)
Pre-op Asphericity ($Z_4^0$)
Post-op Defocus ($Z_2^0$)
Post-op Astigm+ ($Z_2^{-2}$)
Post-op Astigmx ($Z_2^{+2}$)
Post-op Asphericity ($Z_4^0$)
Optical Zone (Defocus)
Optical Zone (Q-factor)
Pre-op Km
Pre-op Q
Centered ALE — Q-factor ($yZ_4^0$)
Frequency
