## Supplemental Notes 2 for "Machine Learning–based Prediction of LASIK Console Inputs for Aspheric Planning (Q-factor, Defocus, Astigmatism): A Translational Methods Study"

Supplementary Figure S2. External-set permutation importance
Target: Defocus ($yZ_2^0$)

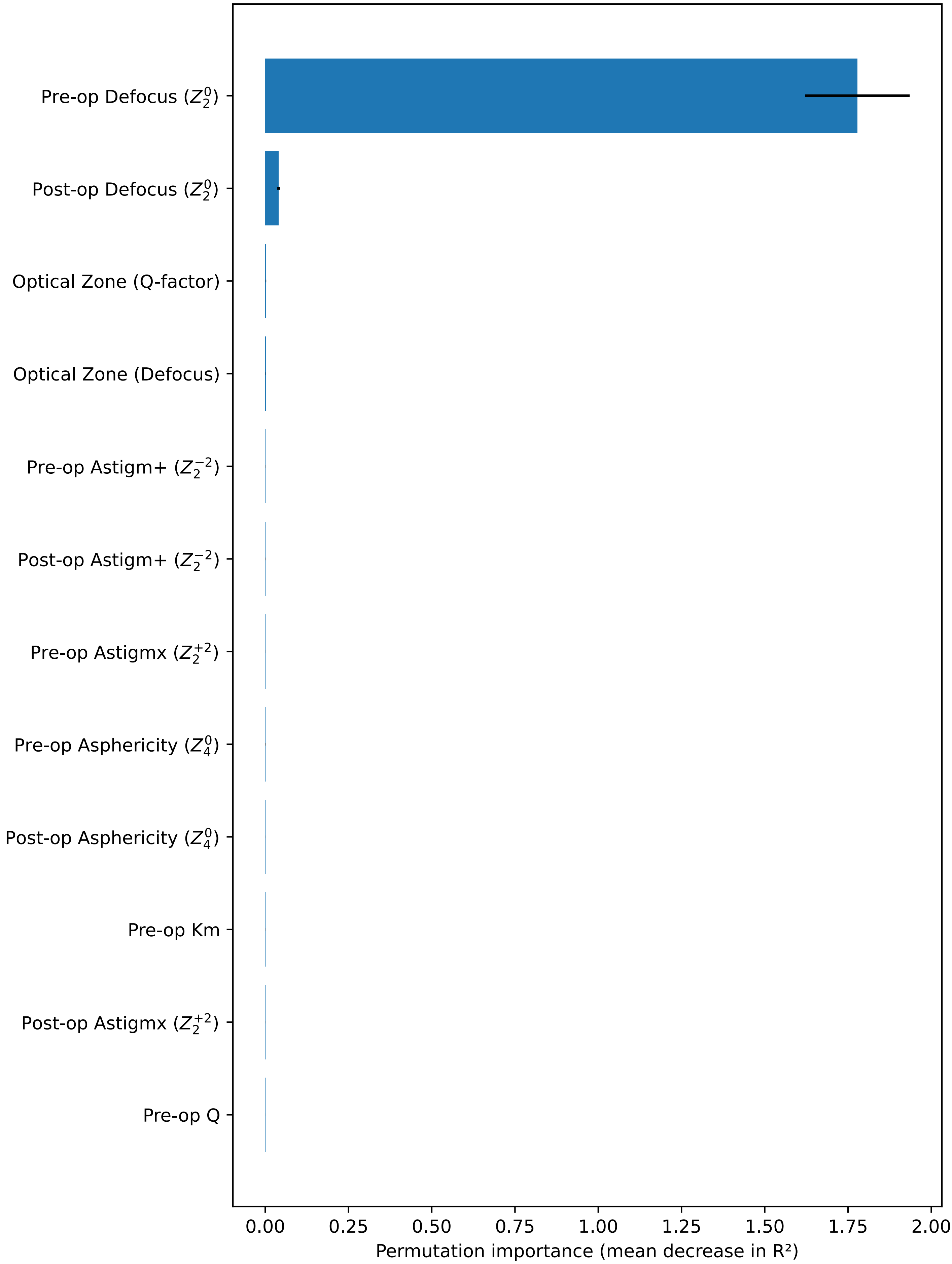


Model: StandardScaler + MultiOutputRegressor(BaggingRegressor(GradientBoostingRegressor)). External performance (test set): $R^2$ = 0.979, MSE = 0.131558, MAE = 0.255349, MedAE = 0.171808. Bars: mean decrease over 50 permutations; error bars: SD.

Supplementary Figure S3. External-set permutation importance
Target: Astigm+ ($yZ_2^{-2}$)

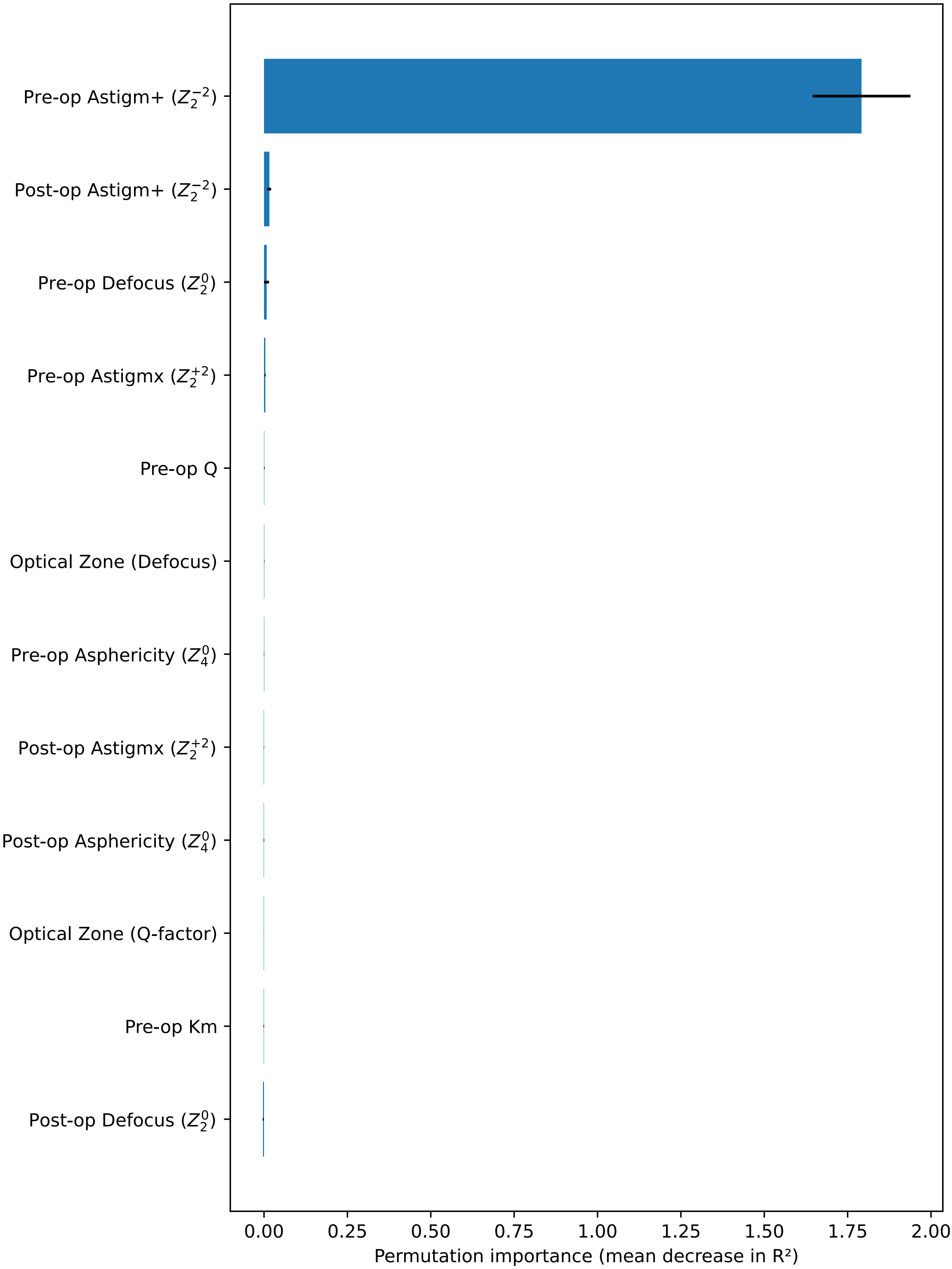


Model: StandardScaler + MultiOutputRegressor(BaggingRegressor(GradientBoostingRegressor)). External performance (test set): $R^2$ = 0.874, MSE = 0.124581, MAE = 0.183650, MedAE = 0.127022. Bars: mean decrease over 50 permutations; error bars: SD.

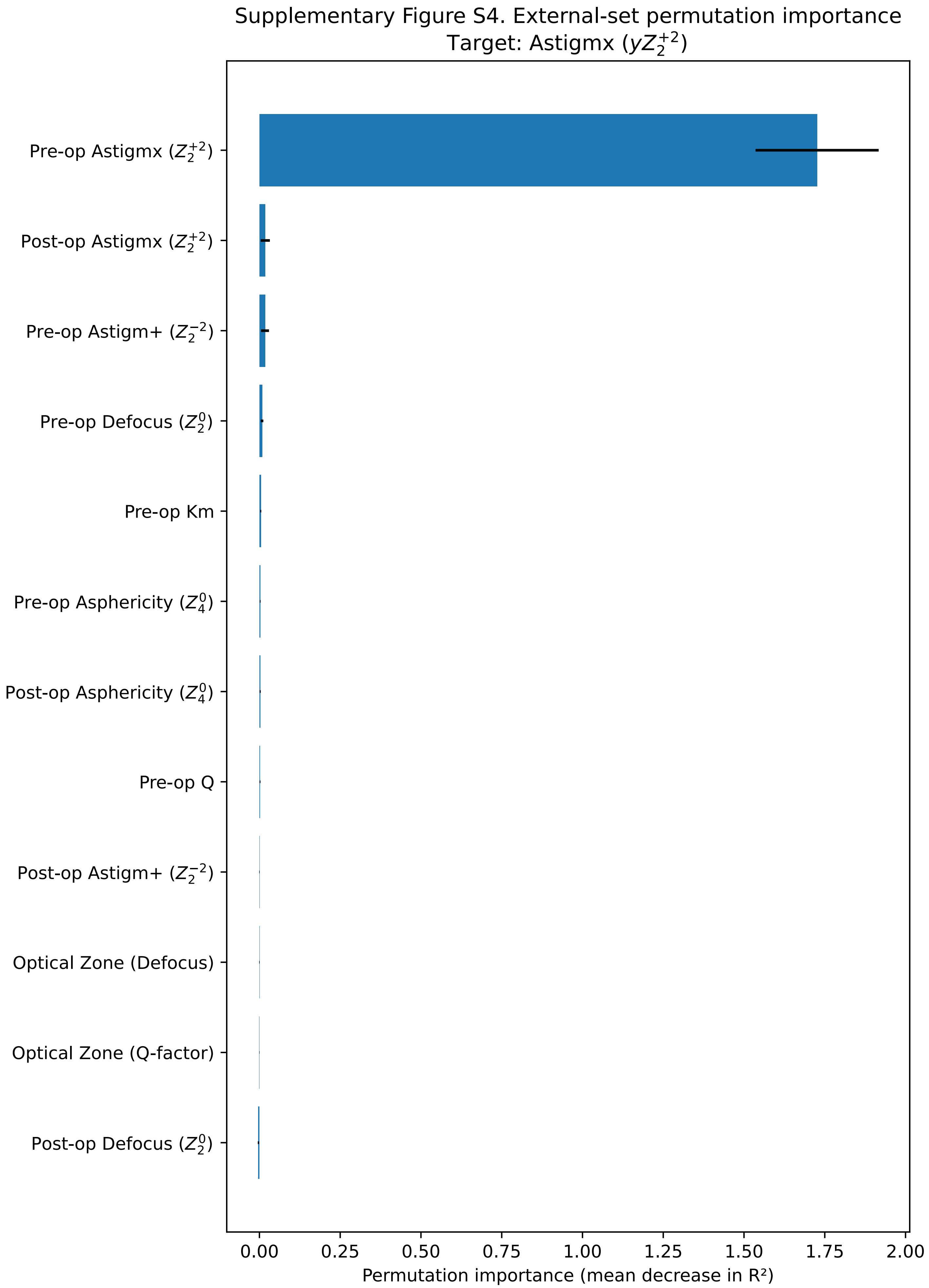


Model: StandardScaler + MultiOutputRegressor(BaggingRegressor(GradientBoostingRegressor)). External performance (test set): $R^2$ = 0.684, MSE = 0.069668, MAE = 0.148310, MedAE = 0.089264. Bars: mean decrease over 50 permutations; error bars: SD.

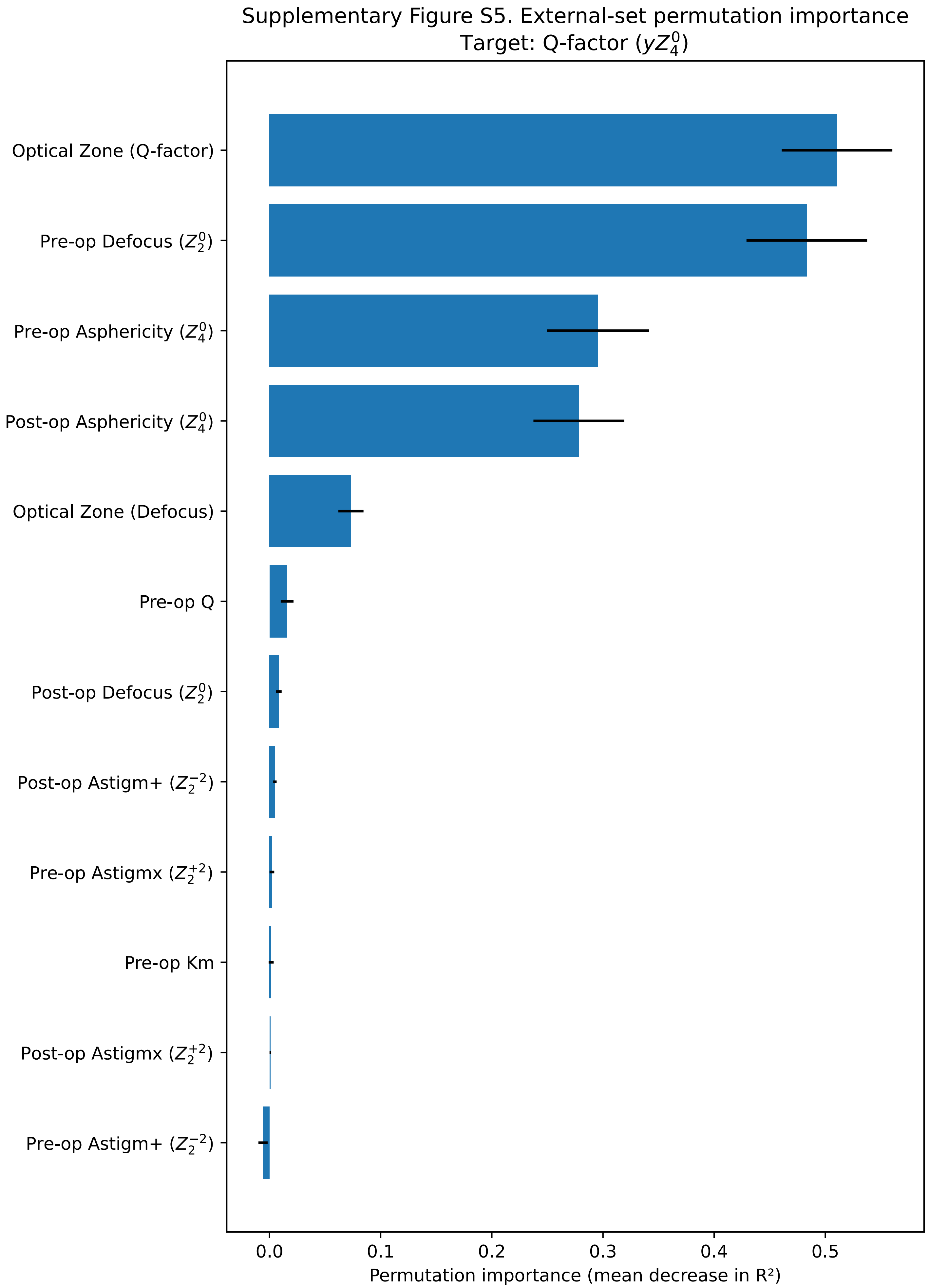


Model: StandardScaler + MultiOutputRegressor(BaggingRegressor(GradientBoostingRegressor)). External performance (test set): $R^2$ = 0.710, MSE = 0.148008, MAE = 0.305308, MedAE = 0.262417. Bars: mean decrease over 50 permutations; error bars: SD.

### Supplementary Table S1. External-set permutation importance (top 8 per target)

#### Target: Defocus ($yZ_2^0$)

| Feature | Mean $\Delta R^2$ | SD |
|---|---|---|
| Pre-op Defocus ($Z_2^0$) | 1.7782 | 0.1570 |
| Post-op Defocus ($Z_2^0$) | 0.0404 | 0.0048 |
| Optical Zone (Q-factor) | 0.0022 | 0.0008 |
| Optical Zone (Defocus) | 0.0019 | 0.0005 |
| Pre-op Astigm+ ($Z_2^{-2}$) | 0.0005 | 0.0002 |
| Post-op Astigm+ ($Z_2^{-2}$) | 0.0003 | 0.0002 |
| Pre-op Astigmx ($Z_2^{+2}$) | 0.0002 | 0.0001 |
| Pre-op Asphericity ($Z_4^0$) | 0.0002 | 0.0003 |

#### Target: Astigm+ ($yZ_2^{-2}$)

| Feature | Mean $\Delta R^2$ | SD |
|---|---|---|
| Pre-op Astigm+ ($Z_2^{-2}$) | 1.7916 | 0.1467 |
| Post-op Astigm+ ($Z_2^{-2}$) | 0.0157 | 0.0057 |
| Pre-op Defocus ($Z_2^0$) | 0.0075 | 0.0075 |
| Pre-op Astigmx ($Z_2^{+2}$) | 0.0035 | 0.0015 |
| Pre-op Q | 0.0008 | 0.0010 |
| Optical Zone (Defocus) | 0.0006 | 0.0006 |
| Pre-op Asphericity ($Z_4^0$) | 0.0001 | 0.0006 |
| Post-op Astigmx ($Z_2^{+2}$) | -0.0002 | 0.0005 |

#### Target: Astigmx ($yZ_2^{+2}$)

| Feature | Mean $\Delta R^2$ | SD |
|---|---|---|
| Pre-op Astigmx ($Z_2^{+2}$) | 1.7263 | 0.1906 |
| Post-op Astigmx ($Z_2^{+2}$) | 0.0181 | 0.0142 |
| Pre-op Astigm+ ($Z_2^{-2}$) | 0.0173 | 0.0122 |
| Pre-op Defocus ($Z_2^0$) | 0.0082 | 0.0040 |
| Pre-op Km | 0.0041 | 0.0016 |
| Pre-op Asphericity ($Z_4^0$) | 0.0024 | 0.0011 |
| Post-op Asphericity ($Z_4^0$) | 0.0023 | 0.0016 |
| Pre-op Q | 0.0019 | 0.0013 |

#### Target: Q-factor ($yZ_4^0$)

| Feature | Mean $\Delta R^2$ | SD |
|---|---|---|
| Optical Zone (Q-factor) | 0.5106 | 0.0498 |
| Pre-op Defocus ($Z_2^0$) | 0.4834 | 0.0543 |
| Pre-op Asphericity ($Z_4^0$) | 0.2954 | 0.0460 |
| Post-op Asphericity ($Z_4^0$) | 0.2783 | 0.0409 |
| Optical Zone (Defocus) | 0.0733 | 0.0113 |
| Pre-op Q | 0.0159 | 0.0058 |
| Post-op Defocus ($Z_2^0$) | 0.0083 | 0.0026 |
| Post-op Astigm+ ($Z_2^{-2}$) | 0.0049 | 0.0015 |

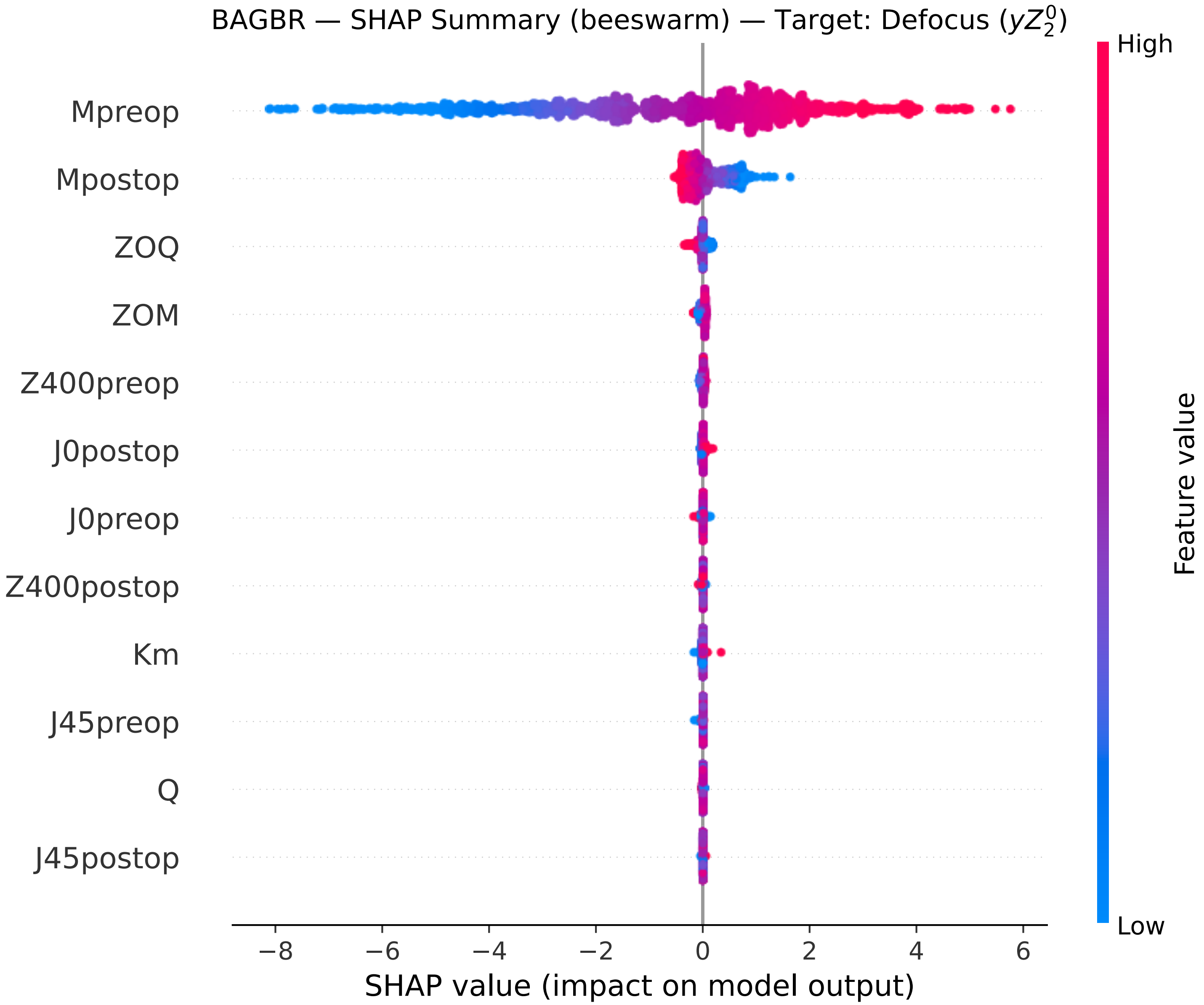
BAGBR — SHAP Summary (beeswarm) — Target: Defocus ($yZ_2^0$)
Mpreop
Mpostop
ZOQ
ZOM
Z400preop
J0postop
J0preop
Z400postop
Km
J45preop
Q
J45postop
−8
−6
−4
−2
0
2
4
6
SHAP value (impact on model output)
High
Low
Feature value

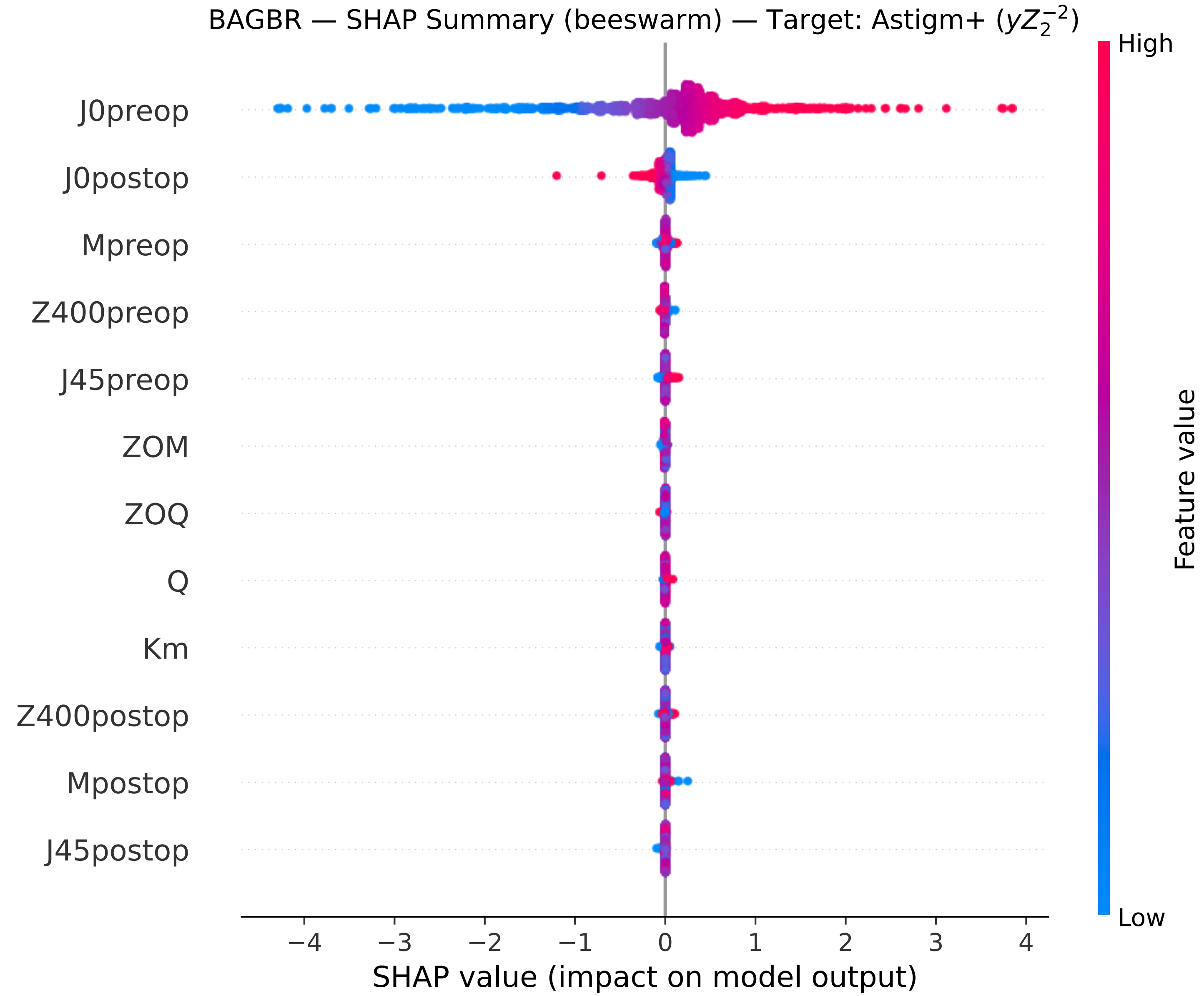

BAGBR — SHAP Summary (beeswarm) — Target: Astigm+ ($yZ_2^{-2}$)
J0preop
J0postop
Mpreop
Z400preop
J45preop
ZOM
ZOQ
Q
Km
Z400postop
Mpostop
J45postop
−4
−3
−2
−1
0
1
2
3
4
SHAP value (impact on model output)
High
Low
Feature value

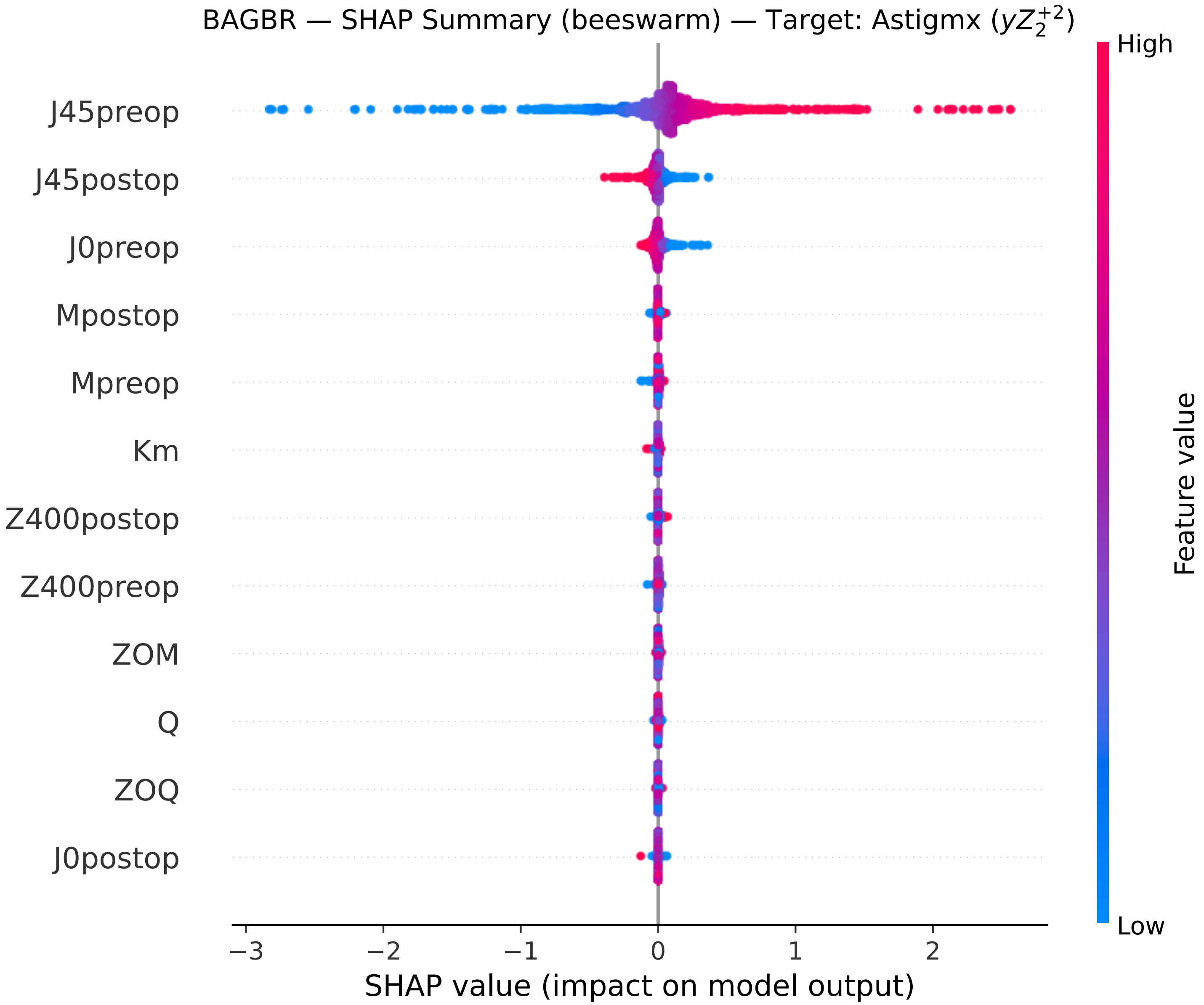

BAGBR — SHAP Summary (beeswarm) — Target: Astigmx ($yZ_2^{+2}$)
J45preop
J45postop
J0preop
Mpostop
Mpreop
Km
Z400postop
Z400preop
ZOM
Q
ZOQ
J0postop
−3
−2
−1
0
1
2
SHAP value (impact on model output)
High
Low
Feature value

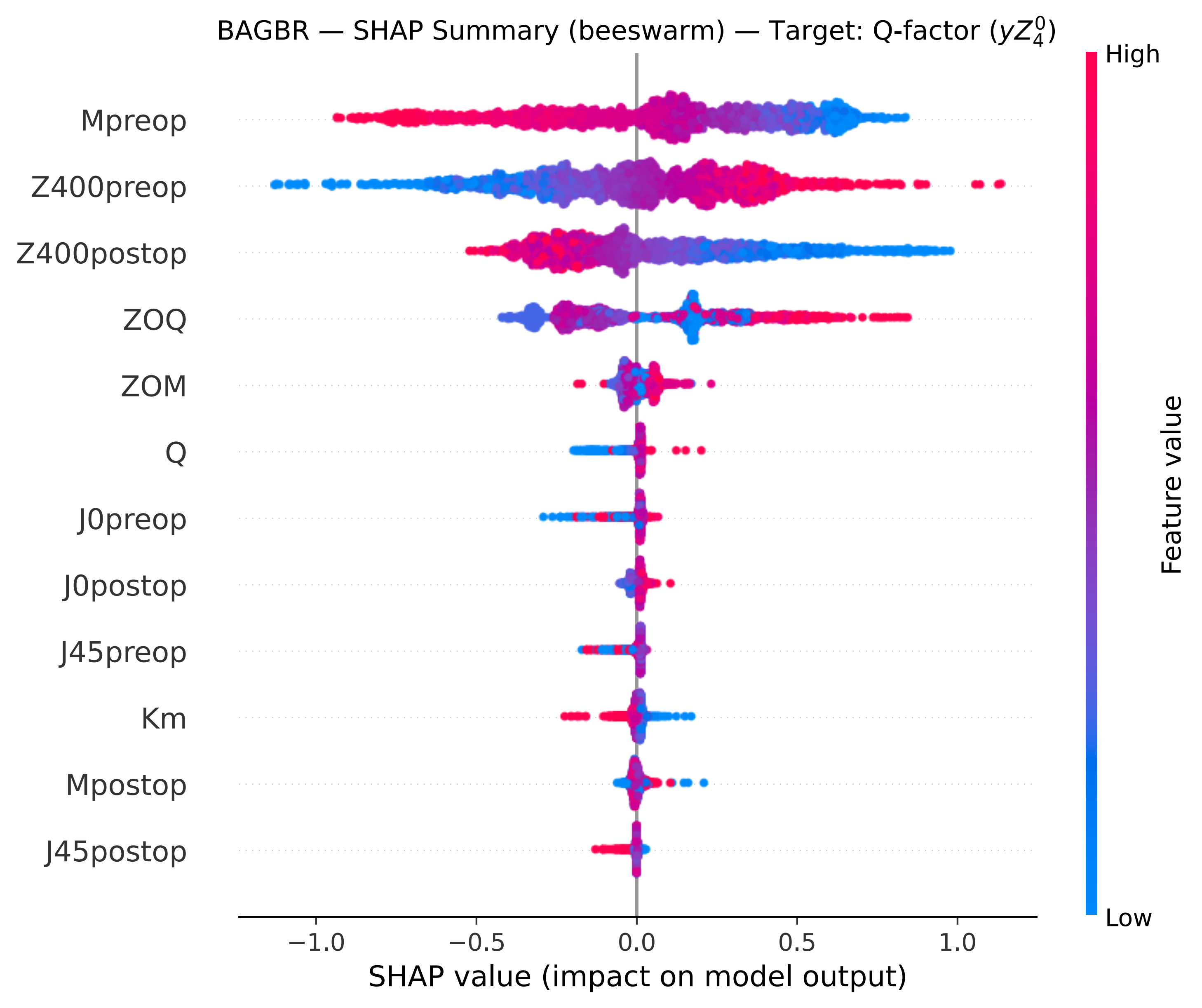

BAGBR — SHAP Summary (beeswarm) — Target: Q-factor ($yZ_4^0$)
Mpreop
Z400preop
Z400postop
ZOQ
ZOM
Q
J0preop
J0postop
J45preop
Km
Mpostop
J45postop
−1.0
−0.5
0.0
0.5
1.0
SHAP value (impact on model output)
High
Low
Feature value

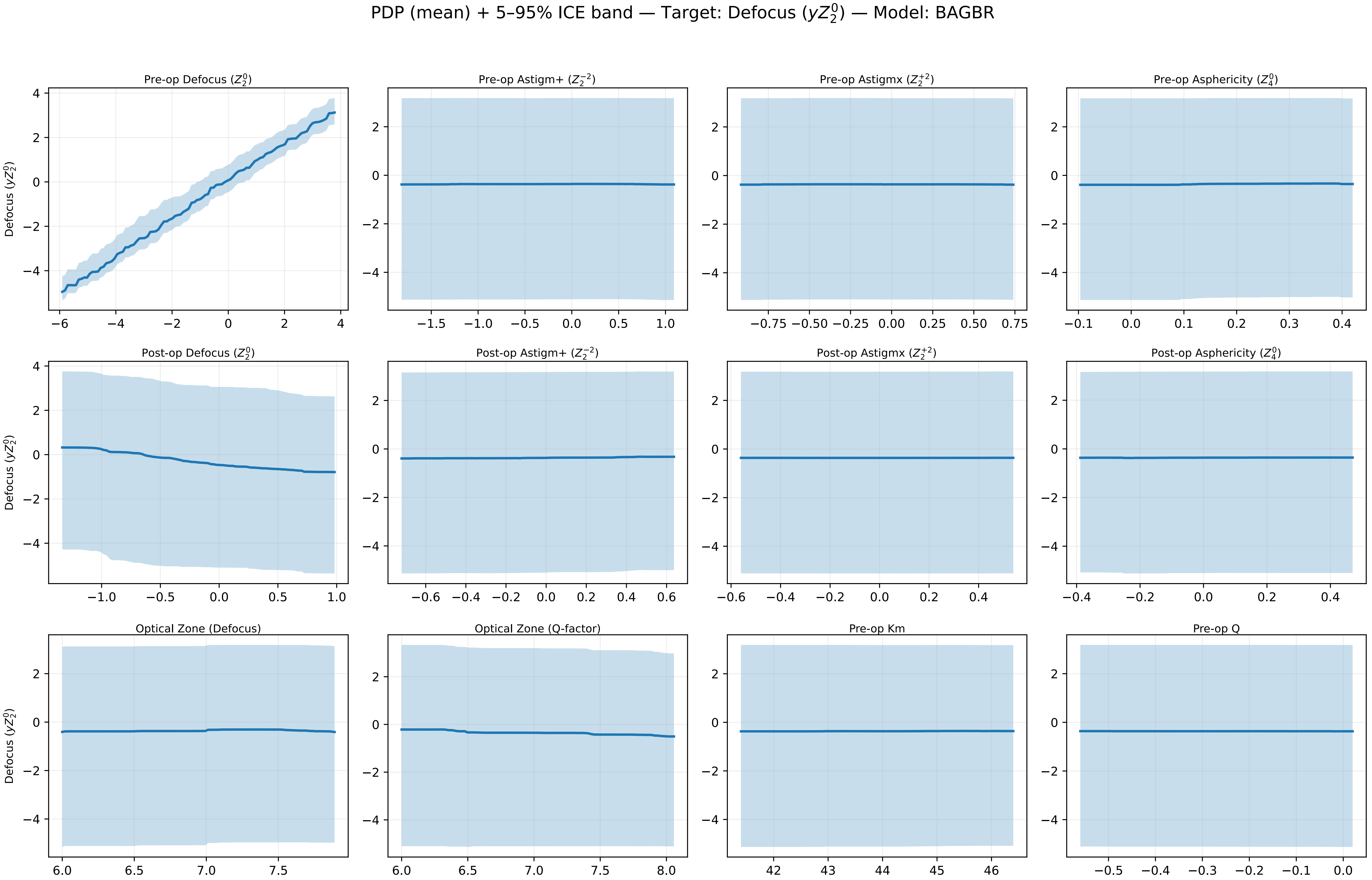
PDP (mean) + 5–95% ICE band — Target: Defocus ($yZ_2^0$) — Model: BAGBR
Pre-op Defocus ($Z_2^0$)
Pre-op Astigm+ ($Z_2^{-2}$)
Pre-op Astigmx ($Z_2^{+2}$)
Pre-op Asphericity ($Z_4^0$)
Post-op Defocus ($Z_2^0$)
Post-op Astigm+ ($Z_2^{-2}$)
Post-op Astigmx ($Z_2^{+2}$)
Post-op Asphericity ($Z_4^0$)
Optical Zone (Defocus)
Optical Zone (Q-factor)
Pre-op Km
Pre-op Q
Defocus ($yZ_2^0$)

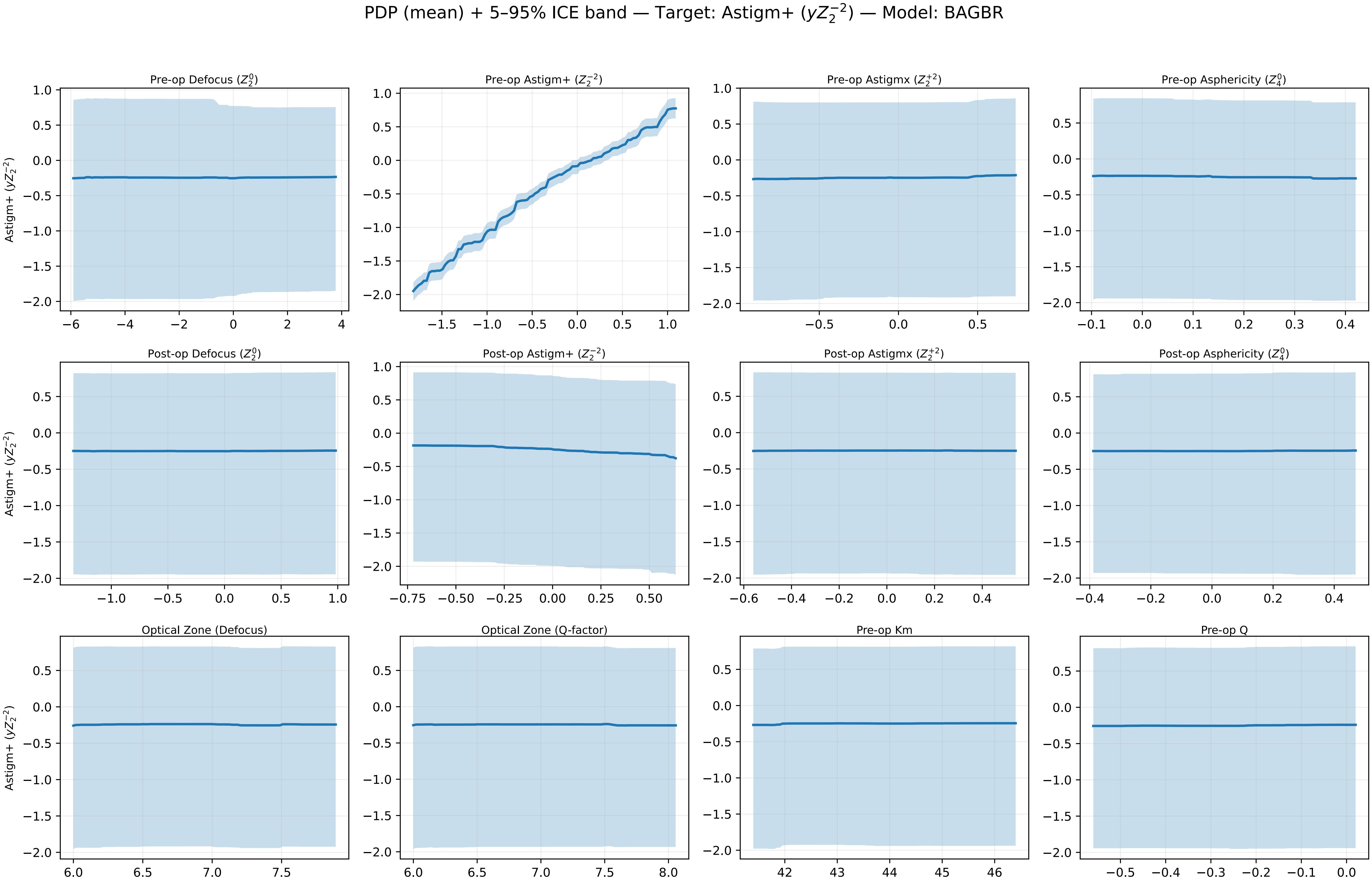
PDP (mean) + 5–95% ICE band — Target: Astigm+ ($yZ_2^{-2}$) — Model: BAGBR
Pre-op Defocus ($Z_2^0$)
Pre-op Astigm+ ($Z_2^{-2}$)
Pre-op Astigmx ($Z_2^{+2}$)
Pre-op Asphericity ($Z_4^0$)
Post-op Defocus ($Z_2^0$)
Post-op Astigm+ ($Z_2^{-2}$)
Post-op Astigmx ($Z_2^{+2}$)
Post-op Asphericity ($Z_4^0$)
Optical Zone (Defocus)
Optical Zone (Q-factor)
Pre-op Km
Pre-op Q
Astigm+ ($yZ_2^{-2}$)

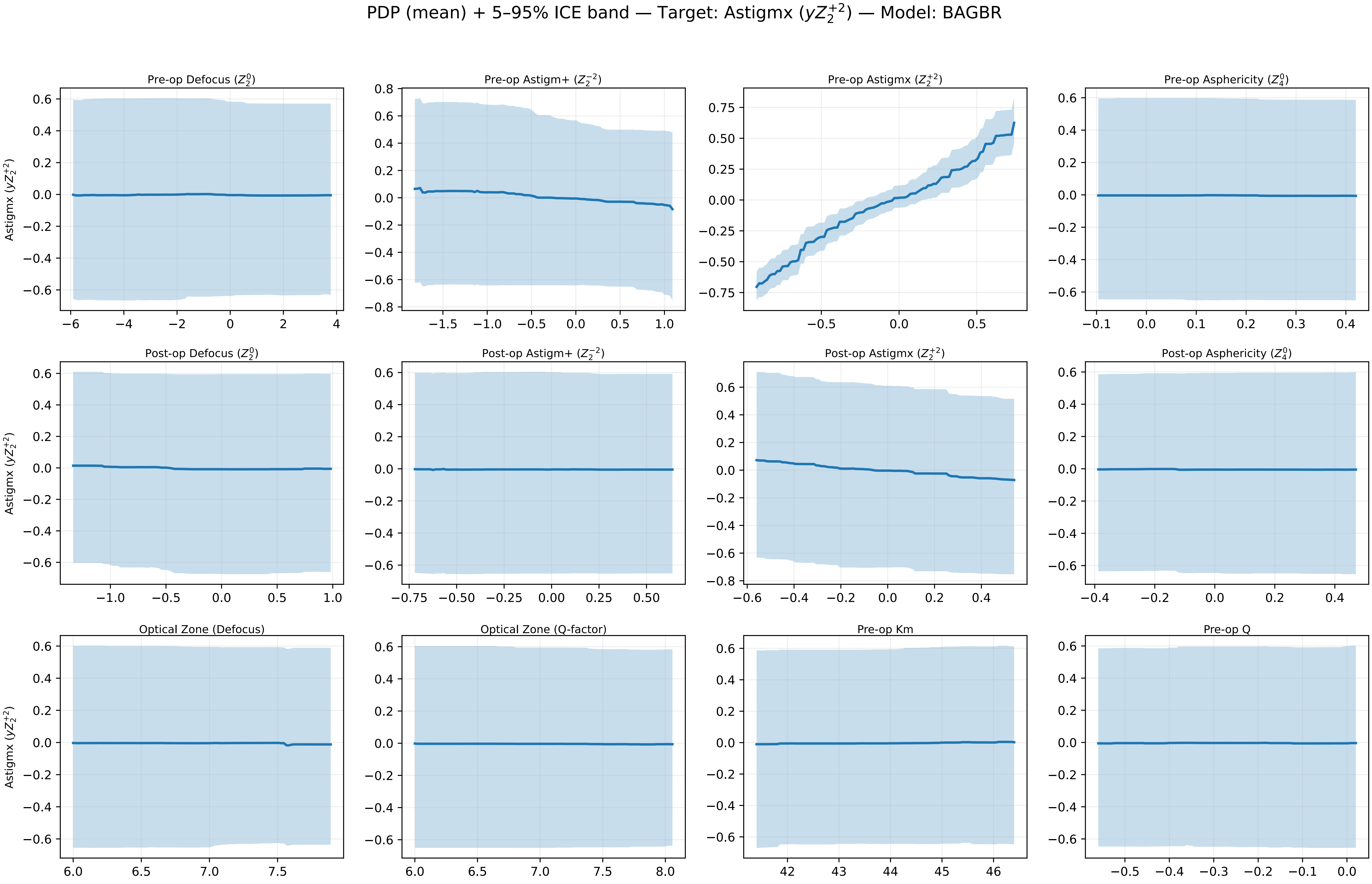

PDP (mean) + 5–95% ICE band — Target: Astigmx ($yZ_2^{+2}$) — Model: BAGBR
Pre-op Defocus ($Z_2^0$)
Pre-op Astigm+ ($Z_2^{-2}$)
Pre-op Astigmx ($Z_2^{+2}$)
Pre-op Asphericity ($Z_4^0$)
Post-op Defocus ($Z_2^0$)
Post-op Astigm+ ($Z_2^{-2}$)
Post-op Astigmx ($Z_2^{+2}$)
Post-op Asphericity ($Z_4^0$)
Optical Zone (Defocus)
Optical Zone (Q-factor)
Pre-op Km
Pre-op Q
Astigmx ($yZ_2^{+2}$)
0.6
0.4
0.2
0.0
−0.2
−0.4
−0.6
0.8
−0.8
0.75
0.50
0.25
0.00
−0.25
−0.50
−0.75
−6
−4
−2
0
2
4
−1.5
−1.0
−0.5
0.0
0.5
1.0
−0.1
0.1
0.3
−0.50
−0.25
−0.6
6.0
6.5
7.0
7.5
8.0
42
43
44
45
46
−0.3
0.0

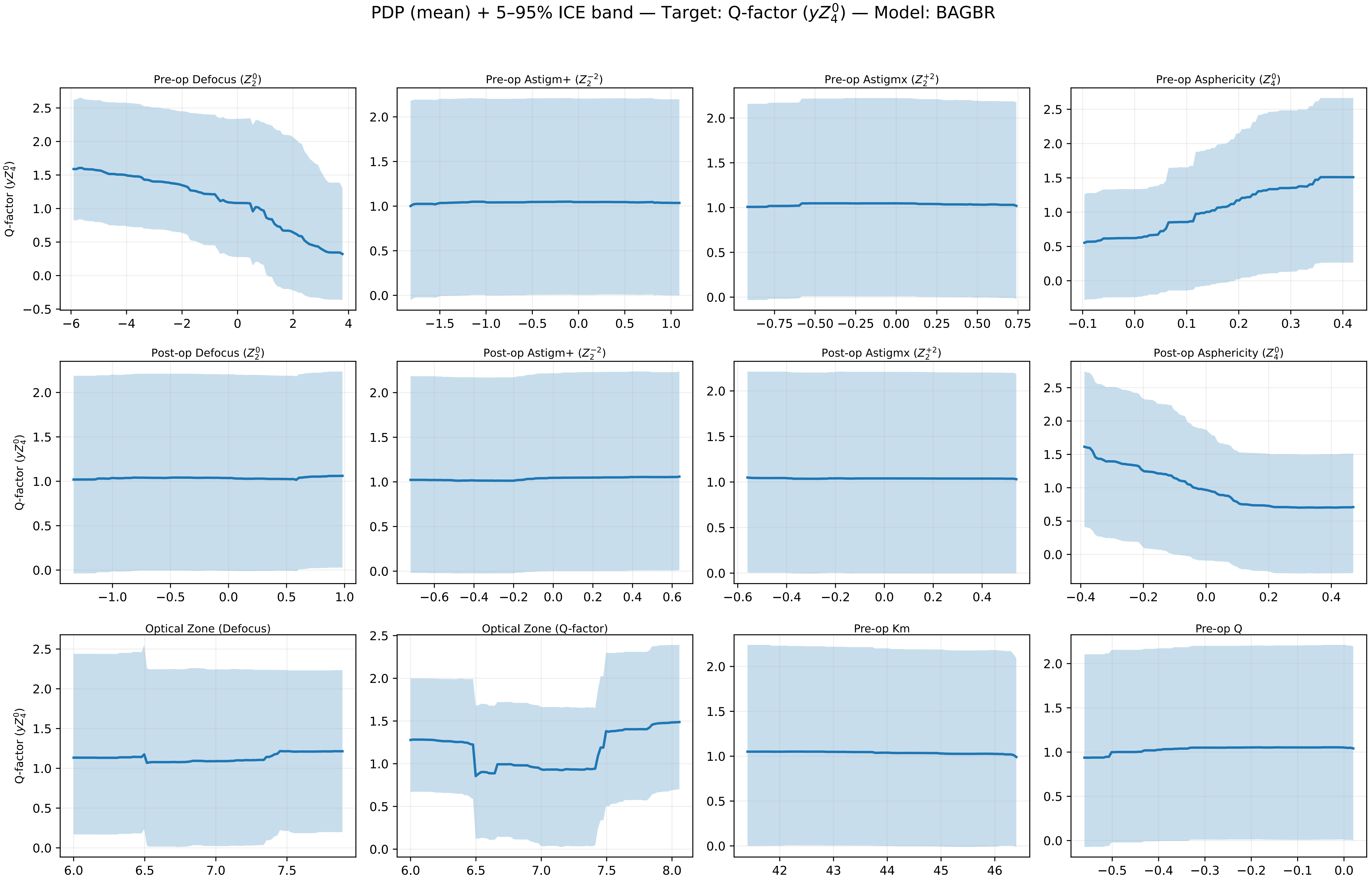
PDP (mean) + 5–95% ICE band — Target: Q-factor ($yZ_4^0$) — Model: BAGBR
Pre-op Defocus ($Z_2^0$)
Pre-op Astigm+ ($Z_2^{-2}$)
Pre-op Astigmx ($Z_2^{+2}$)
Pre-op Asphericity ($Z_4^0$)
Post-op Defocus ($Z_2^0$)
Post-op Astigm+ ($Z_2^{-2}$)
Post-op Astigmx ($Z_2^{+2}$)
Post-op Asphericity ($Z_4^0$)
Optical Zone (Defocus)
Optical Zone (Q-factor)
Pre-op Km
Pre-op Q
Q-factor ($yZ_4^0$)

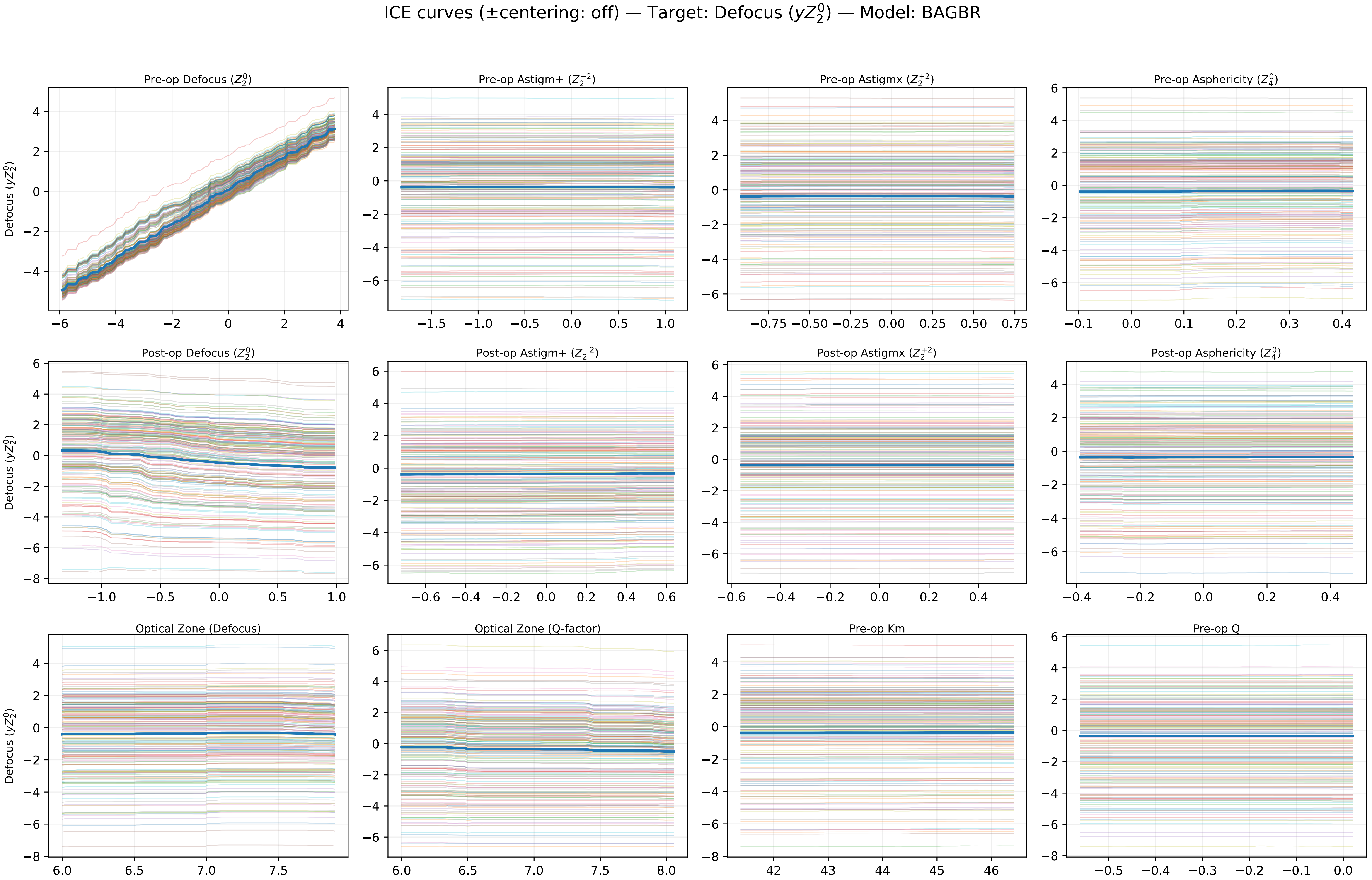

ICE curves (±centering: off) — Target: Defocus ($yZ_2^0$) — Model: BAGBR
Pre-op Defocus ($Z_2^0$)
Pre-op Astigm+ ($Z_2^{-2}$)
Pre-op Astigmx ($Z_2^{+2}$)
Pre-op Asphericity ($Z_4^0$)
Post-op Defocus ($Z_2^0$)
Post-op Astigm+ ($Z_2^{-2}$)
Post-op Astigmx ($Z_2^{+2}$)
Post-op Asphericity ($Z_4^0$)
Optical Zone (Defocus)
Optical Zone (Q-factor)
Pre-op Km
Pre-op Q
Defocus ($yZ_2^0$)

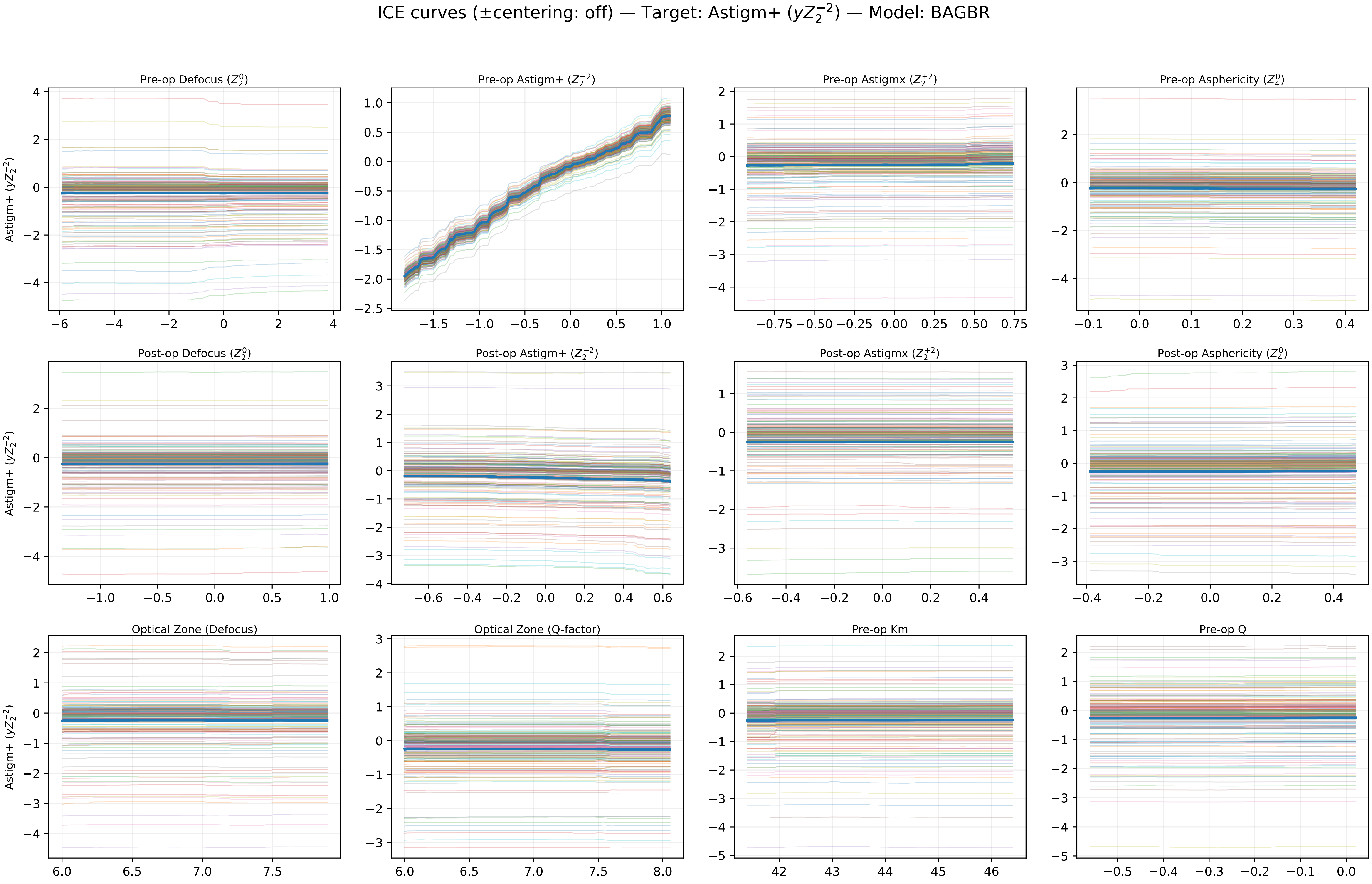

ICE curves (±centering: off) — Target: Astigm+ ($yZ_2^{-2}$) — Model: BAGBR
Pre-op Defocus ($Z_2^0$)
Pre-op Astigm+ ($Z_2^{-2}$)
Pre-op Astigmx ($Z_2^{+2}$)
Pre-op Asphericity ($Z_4^0$)
Post-op Defocus ($Z_2^0$)
Post-op Astigm+ ($Z_2^{-2}$)
Post-op Astigmx ($Z_2^{+2}$)
Post-op Asphericity ($Z_4^0$)
Optical Zone (Defocus)
Optical Zone (Q-factor)
Pre-op Km
Pre-op Q
Astigm+ ($yZ_2^{-2}$)

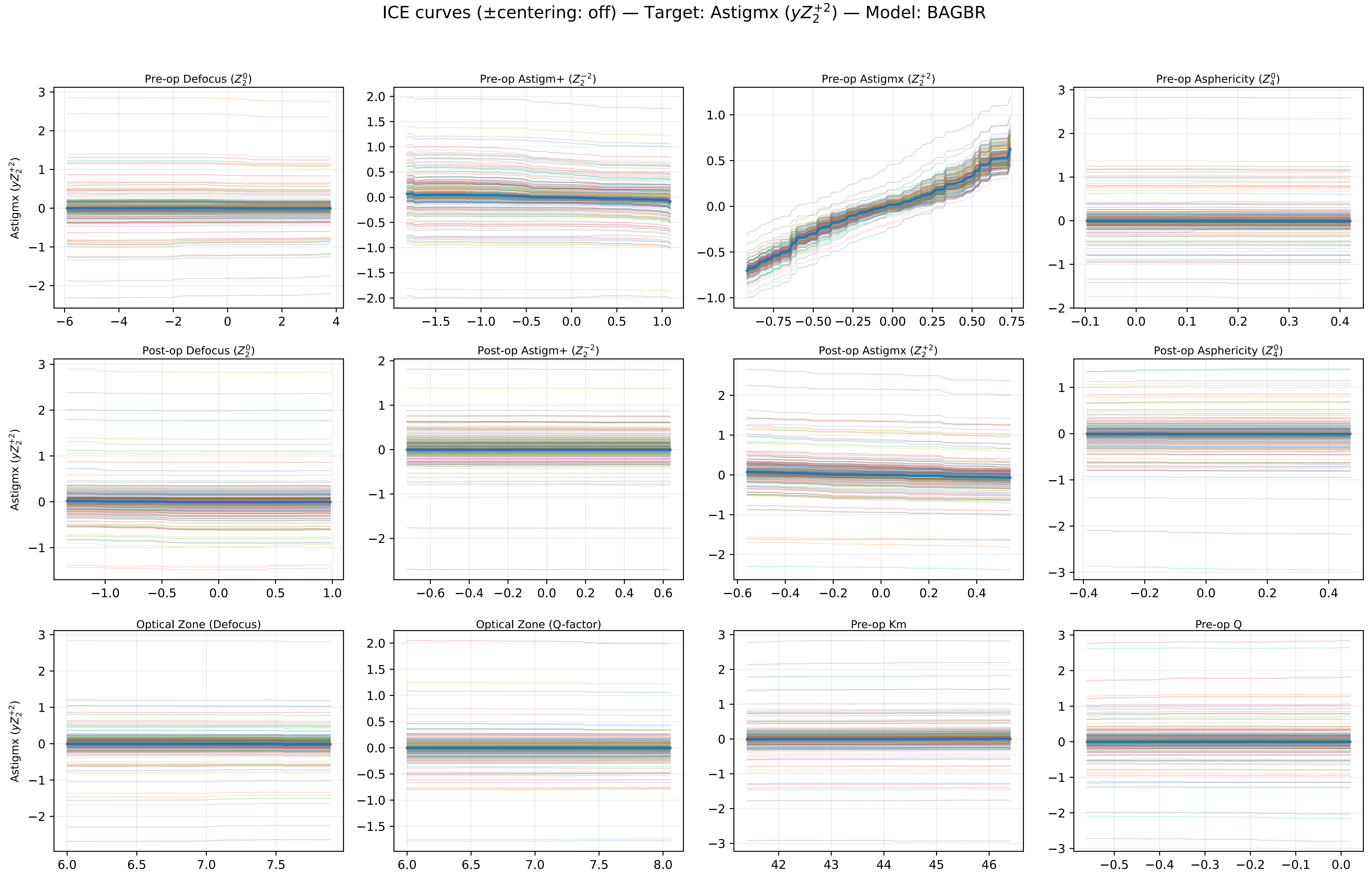

ICE curves (±centering: off) — Target: Astigmx ($yZ_2^{+2}$) — Model: BAGBR
Pre-op Defocus ($Z_2^0$)
Pre-op Astigm+ ($Z_2^{-2}$)
Pre-op Astigmx ($Z_2^{+2}$)
Pre-op Asphericity ($Z_4^0$)
Post-op Defocus ($Z_2^0$)
Post-op Astigm+ ($Z_2^{-2}$)
Post-op Astigmx ($Z_2^{+2}$)
Post-op Asphericity ($Z_4^0$)
Optical Zone (Defocus)
Optical Zone (Q-factor)
Pre-op Km
Pre-op Q
Astigmx ($yZ_2^{+2}$)

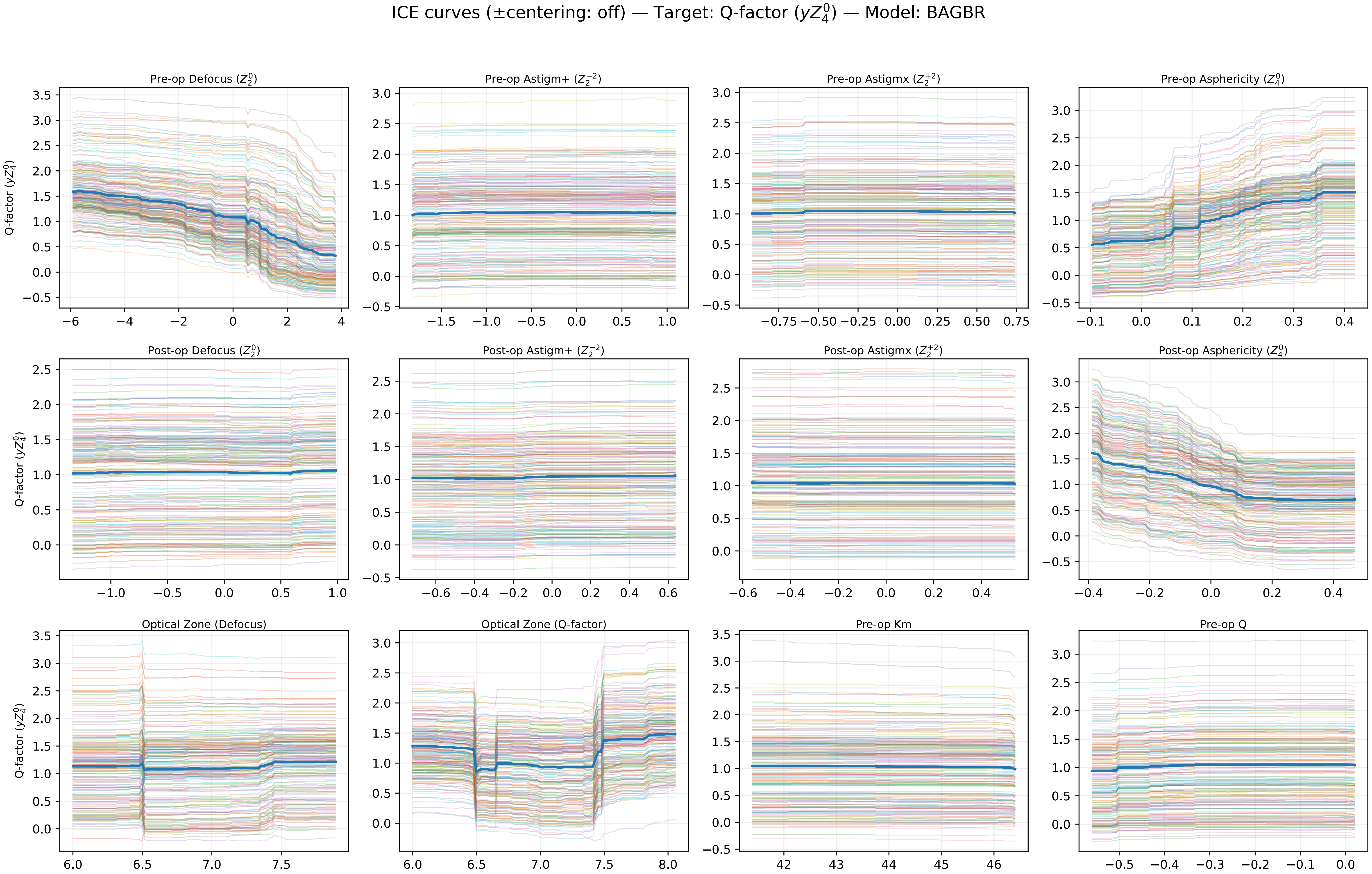
ICE curves (±centering: off) — Target: Q-factor ($yZ_4^0$) — Model: BAGBR
Pre-op Defocus ($Z_2^0$)
Pre-op Astigm+ ($Z_2^{-2}$)
Pre-op Astigmx ($Z_2^{+2}$)
Pre-op Asphericity ($Z_4^0$)
Post-op Defocus ($Z_2^0$)
Post-op Astigm+ ($Z_2^{-2}$)
Post-op Astigmx ($Z_2^{+2}$)
Post-op Asphericity ($Z_4^0$)
Optical Zone (Defocus)
Optical Zone (Q-factor)
Pre-op Km
Pre-op Q
Q-factor ($yZ_4^0$)

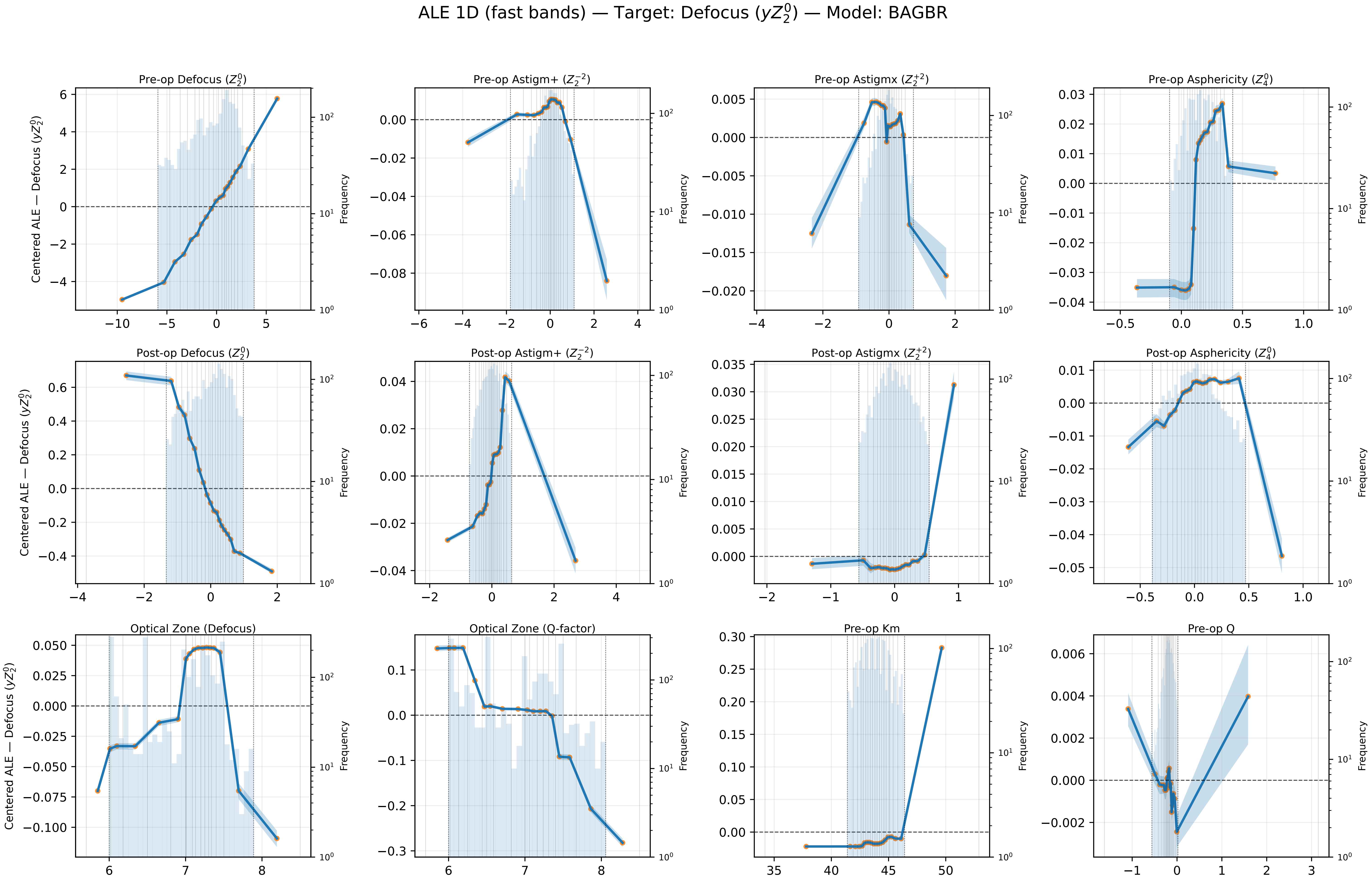

ALE 1D (fast bands) — Target: Defocus ($yZ_2^0$) — Model: BAGBR
Pre-op Defocus ($Z_2^0$)
Pre-op Astigm+ ($Z_2^{-2}$)
Pre-op Astigmx ($Z_2^{+2}$)
Pre-op Asphericity ($Z_4^0$)
Post-op Defocus ($Z_2^0$)
Post-op Astigm+ ($Z_2^{-2}$)
Post-op Astigmx ($Z_2^{+2}$)
Post-op Asphericity ($Z_4^0$)
Optical Zone (Defocus)
Optical Zone (Q-factor)
Pre-op Km
Pre-op Q
Centered ALE — Defocus ($yZ_2^0$)
Frequency

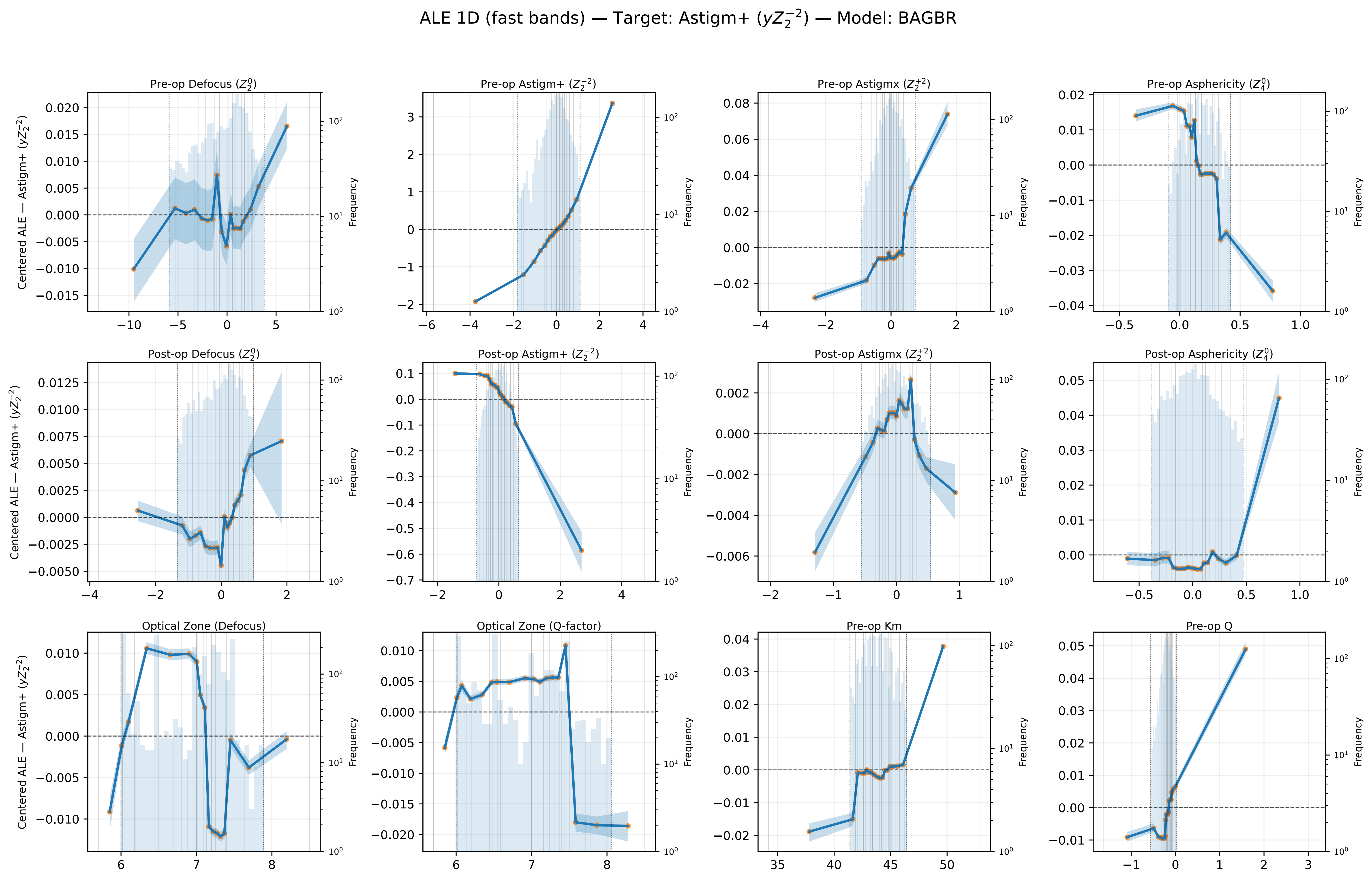
ALE 1D (fast bands) — Target: Astigm+ ($yZ_2^{-2}$) — Model: BAGBR
Pre-op Defocus ($Z_2^0$)
Pre-op Astigm+ ($Z_2^{-2}$)
Pre-op Astigmx ($Z_2^{+2}$)
Pre-op Asphericity ($Z_4^0$)
Post-op Defocus ($Z_2^0$)
Post-op Astigm+ ($Z_2^{-2}$)
Post-op Astigmx ($Z_2^{+2}$)
Post-op Asphericity ($Z_4^0$)
Optical Zone (Defocus)
Optical Zone (Q-factor)
Pre-op Km
Pre-op Q
Centered ALE — Astigm+ ($yZ_2^{-2}$)
Frequency

ALE 1D (fast bands) — Target: Astigmx ($yZ_2^{+2}$) — Model: BAGBR

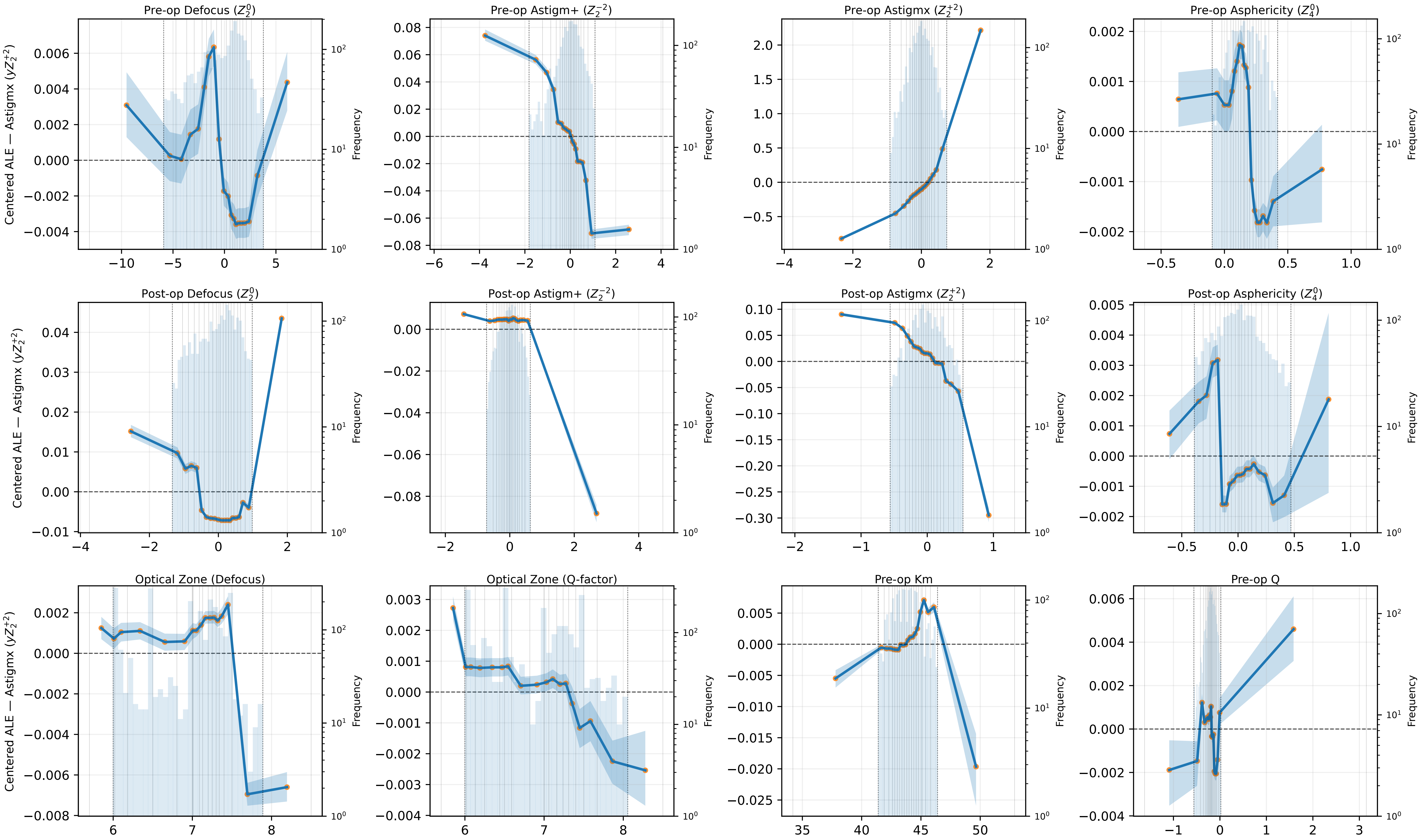

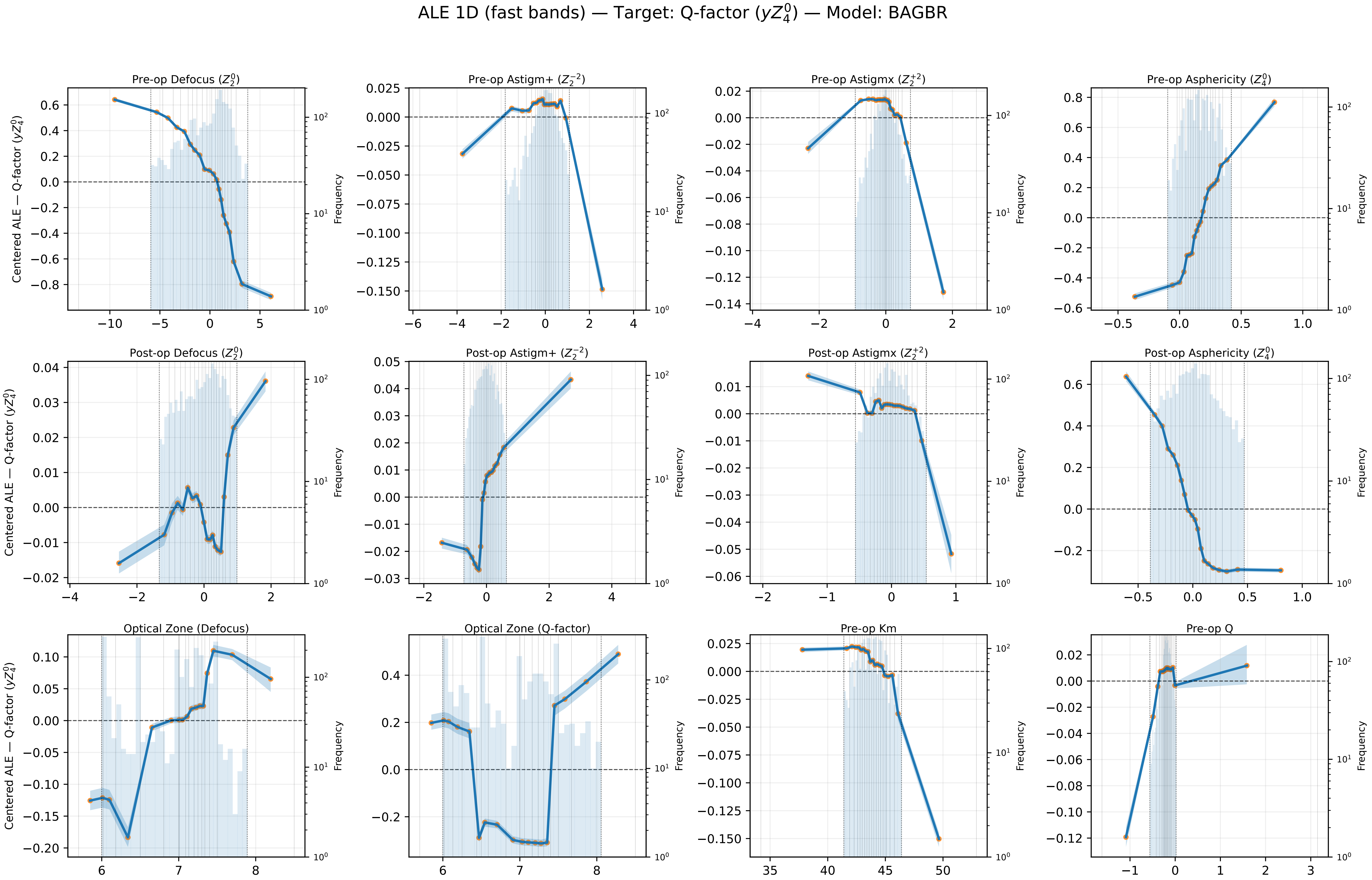

ALE 1D (fast bands) — Target: Q-factor ($yZ_4^0$) — Model: BAGBR
Pre-op Defocus ($Z_2^0$)
Pre-op Astigm+ ($Z_2^{-2}$)
Pre-op Astigmx ($Z_2^{+2}$)
Pre-op Asphericity ($Z_4^0$)
Post-op Defocus ($Z_2^0$)
Post-op Astigm+ ($Z_2^{-2}$)
Post-op Astigmx ($Z_2^{+2}$)
Post-op Asphericity ($Z_4^0$)
Optical Zone (Defocus)
Optical Zone (Q-factor)
Pre-op Km
Pre-op Q
Centered ALE — Q-factor ($yZ_4^0$)
Frequency
