## Supplemental Notes 3 for "Machine Learning–based Prediction of LASIK Console Inputs for Aspheric Planning (Q-factor, Defocus, Astigmatism): A Translational Methods Study"

Supplementary Figure S2. External-set permutation importance
Target: Defocus ($yZ_2^0$)

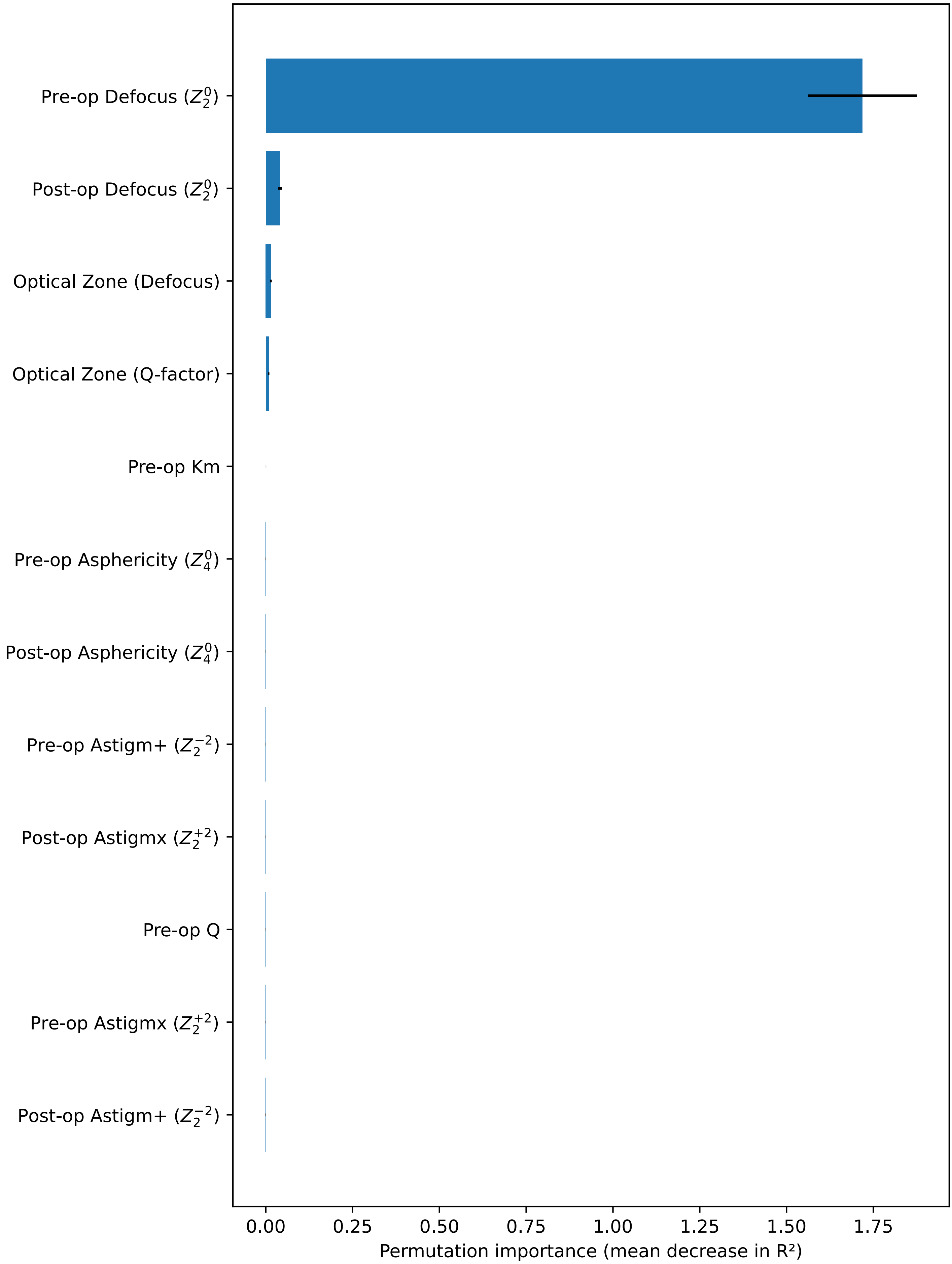


Model: StandardScaler + MultiOutputRegressor(MLP). External performance (test set): $R^2$ = 0.985, MSE = 0.094814, MAE = 0.234681, MedAE = 0.181119. Bars: mean decrease over 50 permutations; error bars: SD.

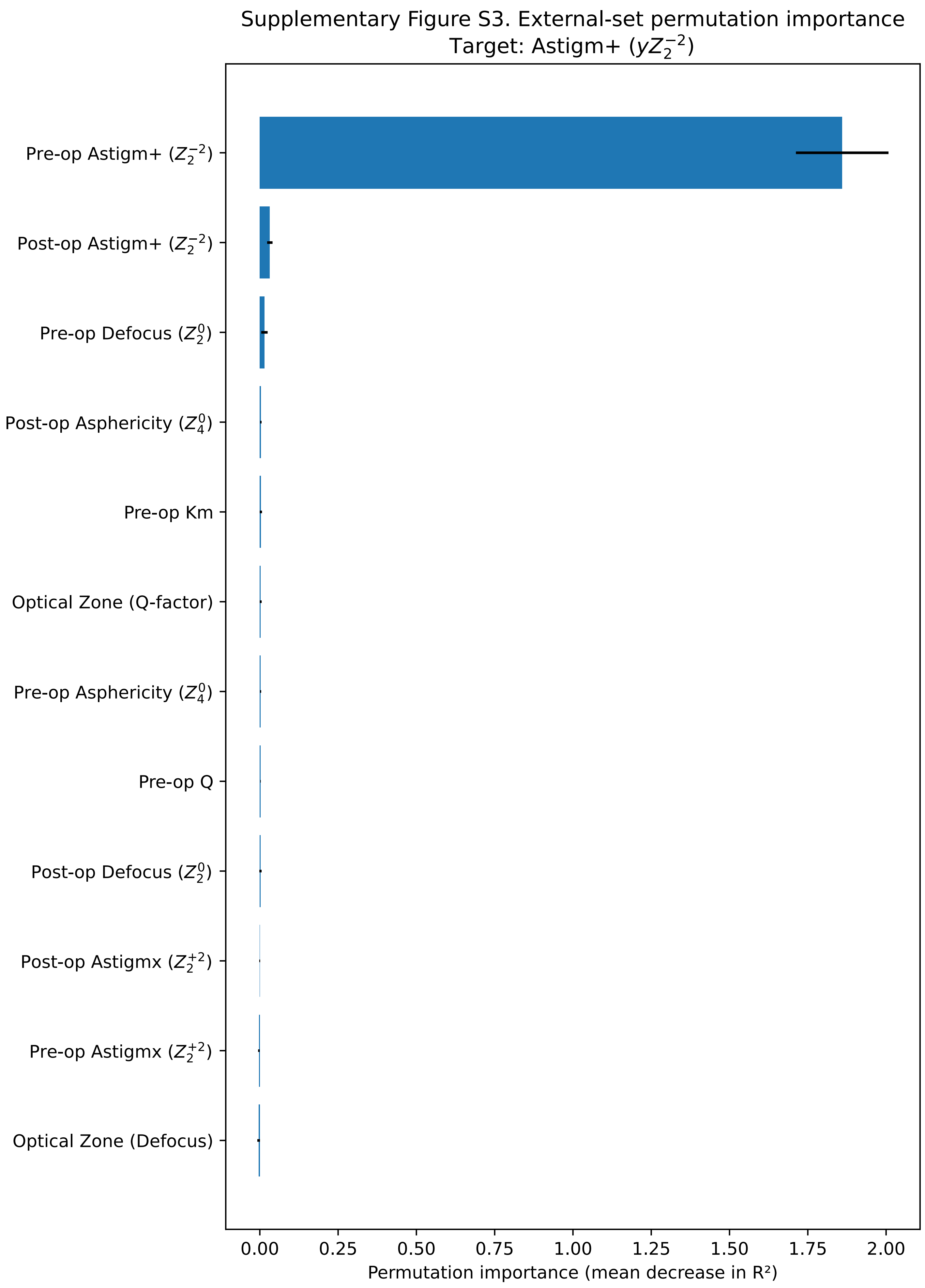


Model: StandardScaler + MultiOutputRegressor(MLP). External performance (test set): $R^2$ = 0.874, MSE = 0.124960, MAE = 0.181028, MedAE = 0.114932. Bars: mean decrease over 50 permutations; error bars: SD.

Supplementary Figure S4. External-set permutation importance
Target: Astigmx ($yZ_2^{+2}$)

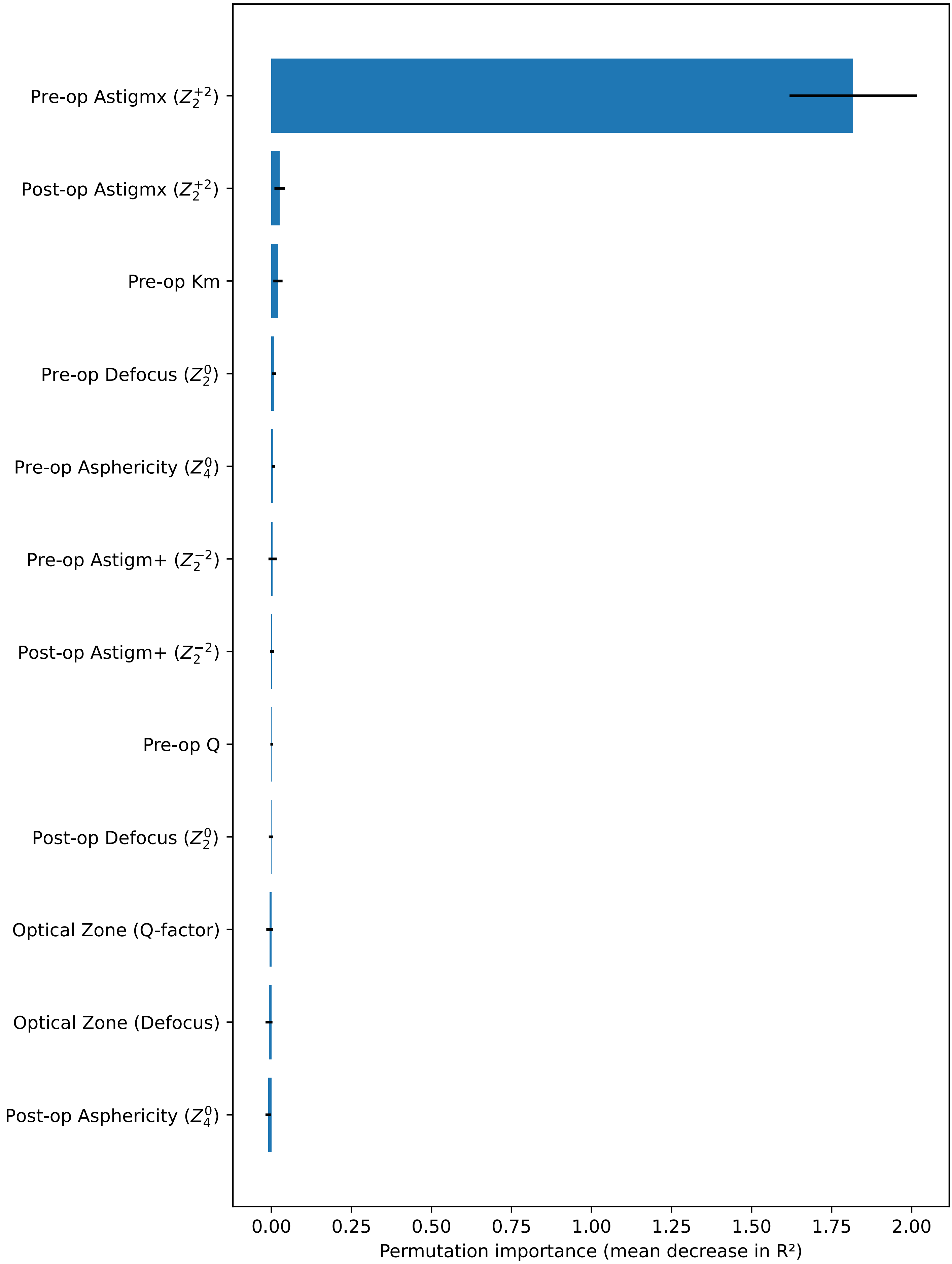


Model: StandardScaler + MultiOutputRegressor(MLP). External performance (test set): $R^2$ = 0.660, MSE = 0.074970, MAE = 0.149685, MedAE = 0.102489. Bars: mean decrease over 50 permutations; error bars: SD.

Supplementary Figure S5. External-set permutation importance
Target: Q-factor ($yZ_4^0$)

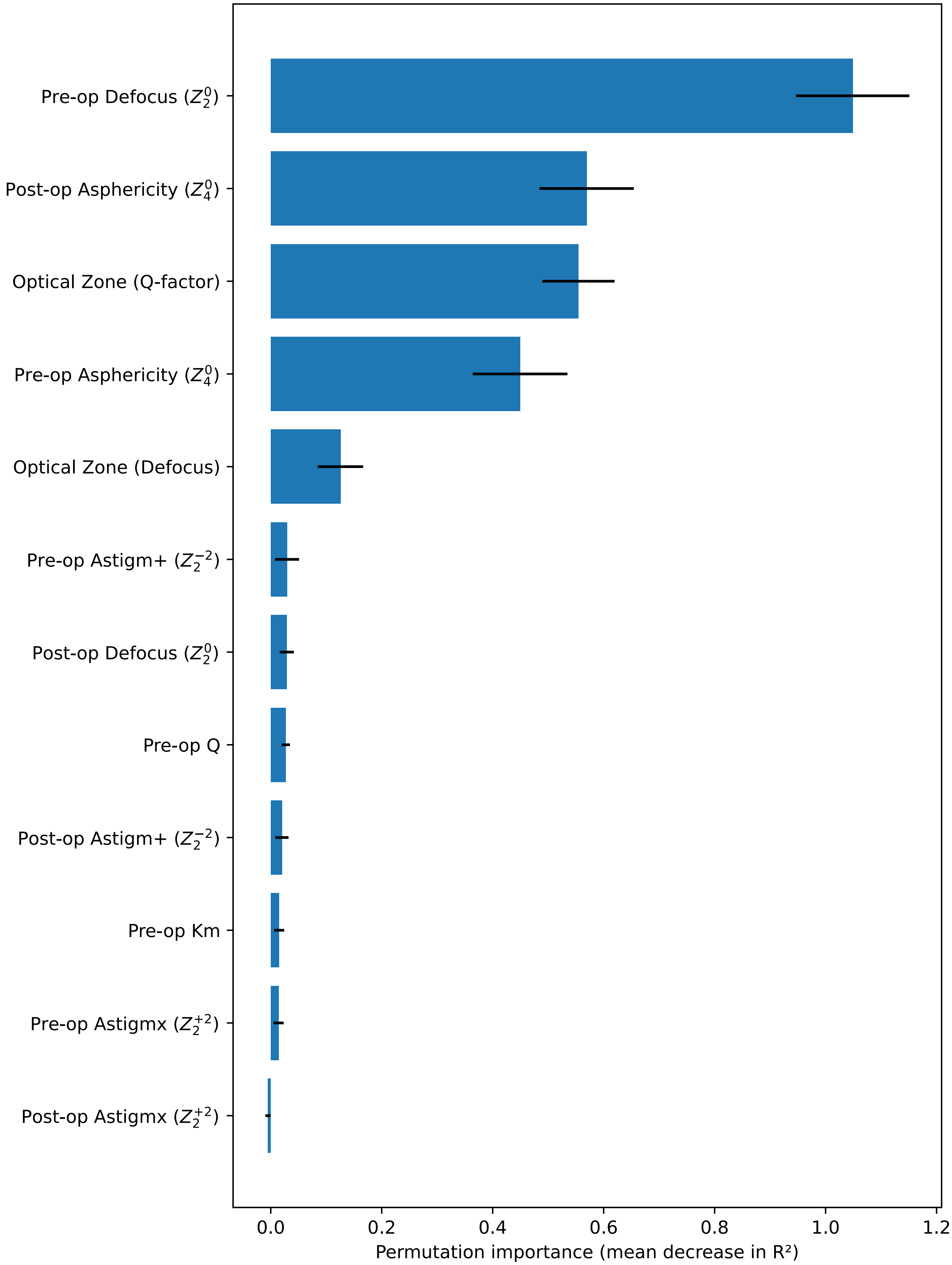


Model: StandardScaler + MultiOutputRegressor(MLP). External performance (test set): $R^2$ = 0.734, MSE = 0.135773, MAE = 0.265580, MedAE = 0.172159. Bars: mean decrease over 50 permutations; error bars: SD.

### Supplementary Table S1. External-set permutation importance (top 8 per target)

#### Target: Defocus ($yZ_2^0$)

| Feature | Mean ΔR² | SD |
|---|---|---|
| Pre-op Defocus ($Z_2^0$) | 1.7186 | 0.1563 |
| Post-op Defocus ($Z_2^0$) | 0.0412 | 0.0053 |
| Optical Zone (Defocus) | 0.0147 | 0.0024 |
| Optical Zone (Q-factor) | 0.0087 | 0.0017 |
| Pre-op Km | 0.0006 | 0.0006 |
| Pre-op Asphericity ($Z_4^0$) | 0.0001 | 0.0008 |
| Post-op Asphericity ($Z_4^0$) | 0.0001 | 0.0006 |
| Pre-op Astigm+ ($Z_2^{-2}$) | 0.0001 | 0.0006 |

#### Target: Astigm+ ($yZ_2^{-2}$)

| Feature | Mean ΔR² | SD |
|---|---|---|
| Pre-op Astigm+ ($Z_2^{-2}$) | 1.8599 | 0.1478 |
| Post-op Astigm+ ($Z_2^{-2}$) | 0.0320 | 0.0086 |
| Pre-op Defocus ($Z_2^0$) | 0.0147 | 0.0102 |
| Post-op Asphericity ($Z_4^0$) | 0.0033 | 0.0022 |
| Pre-op Km | 0.0033 | 0.0036 |
| Optical Zone (Q-factor) | 0.0028 | 0.0031 |
| Pre-op Asphericity ($Z_4^0$) | 0.0024 | 0.0019 |
| Pre-op Q | 0.0022 | 0.0008 |

#### Target: Astigmx ($yZ_2^{+2}$)

| Feature | Mean ΔR² | SD |
|---|---|---|
| Pre-op Astigmx ($Z_2^{+2}$) | 1.8172 | 0.1986 |
| Post-op Astigmx ($Z_2^{+2}$) | 0.0261 | 0.0166 |
| Pre-op Km | 0.0204 | 0.0144 |
| Pre-op Defocus ($Z_2^0$) | 0.0089 | 0.0062 |
| Pre-op Asphericity ($Z_4^0$) | 0.0061 | 0.0048 |
| Pre-op Astigm+ ($Z_2^{-2}$) | 0.0038 | 0.0129 |
| Post-op Astigm+ ($Z_2^{-2}$) | 0.0025 | 0.0063 |
| Pre-op Q | 0.0009 | 0.0041 |

#### Target: Q-factor ($yZ_4^0$)

| Feature | Mean ΔR² | SD |
|---|---|---|
| Pre-op Defocus ($Z_2^0$) | 1.0489 | 0.1022 |
| Post-op Asphericity ($Z_4^0$) | 0.5693 | 0.0850 |
| Optical Zone (Q-factor) | 0.5546 | 0.0651 |
| Pre-op Asphericity ($Z_4^0$) | 0.4494 | 0.0853 |
| Optical Zone (Defocus) | 0.1257 | 0.0409 |
| Pre-op Astigm+ ($Z_2^{-2}$) | 0.0293 | 0.0217 |
| Post-op Defocus ($Z_2^0$) | 0.0290 | 0.0127 |
| Pre-op Q | 0.0271 | 0.0075 |

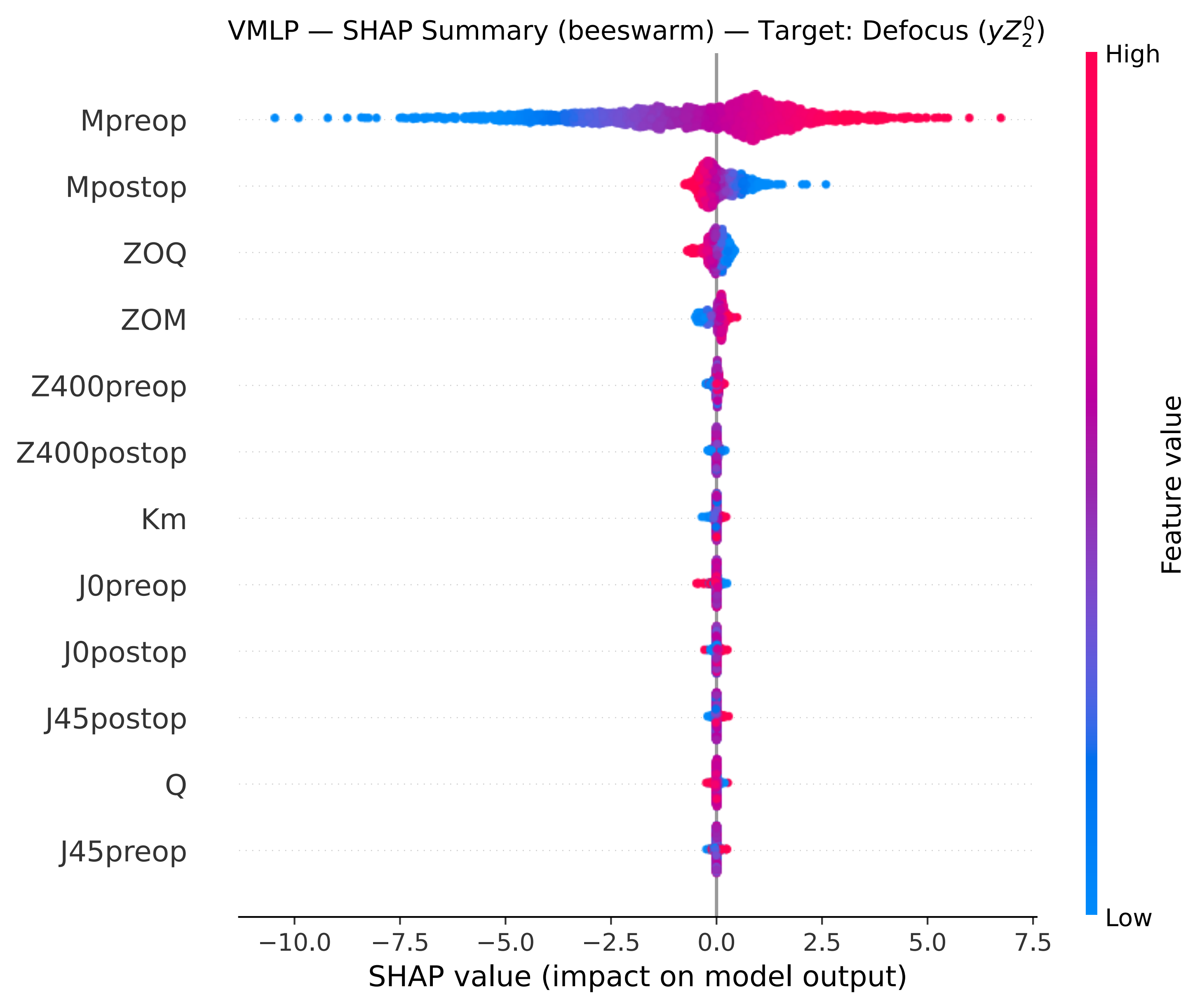

VMLP — SHAP Summary (beeswarm) — Target: Defocus ($yZ_2^0$)
Mpreop
Mpostop
ZOQ
ZOM
Z400preop
Z400postop
Km
J0preop
J0postop
J45postop
Q
J45preop
−10.0
−7.5
−5.0
−2.5
0.0
2.5
5.0
7.5
SHAP value (impact on model output)
High
Low
Feature value

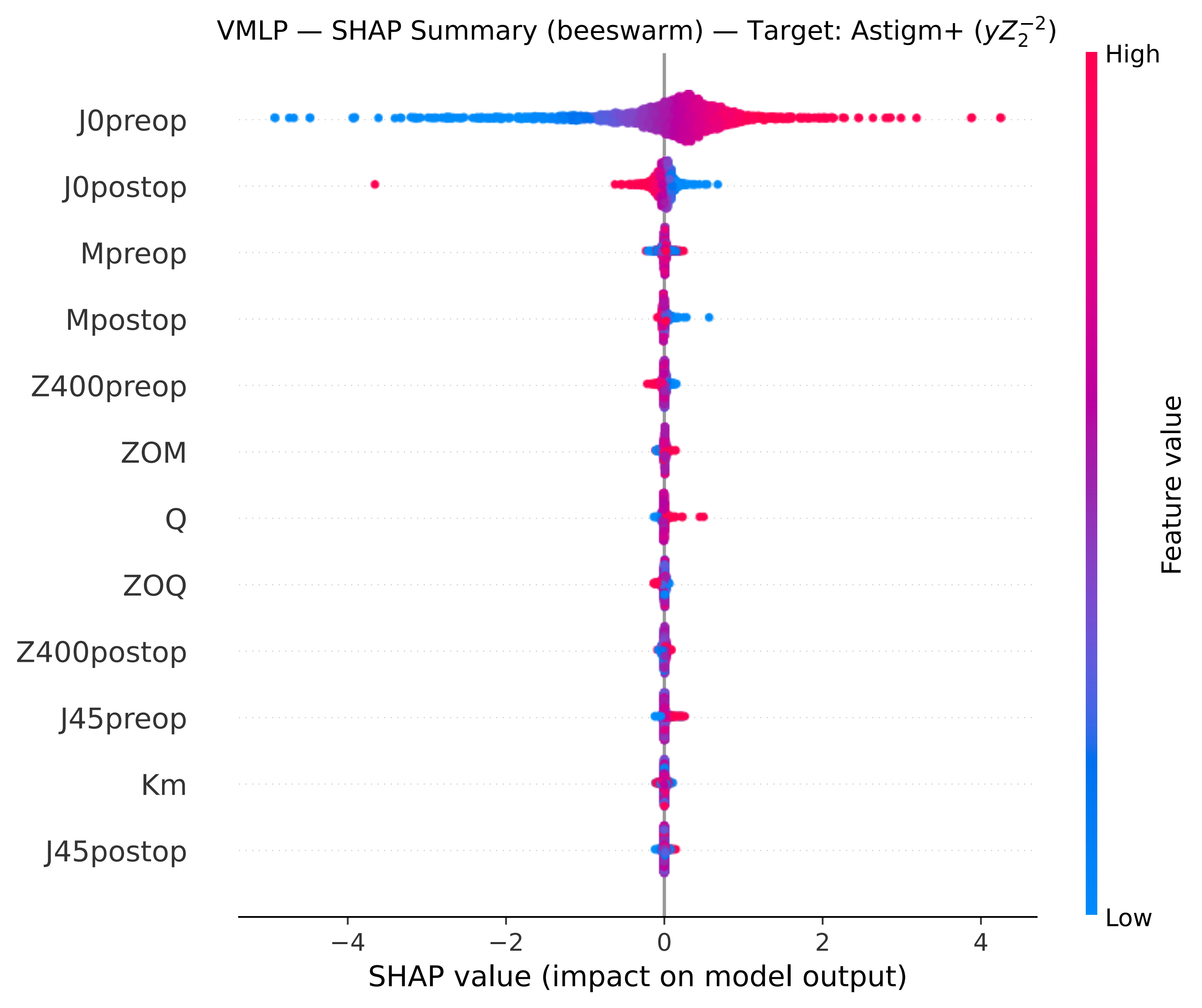
VMLP — SHAP Summary (beeswarm) — Target: Astigm+ ($yZ_2^{-2}$)
J0preop
J0postop
Mpreop
Mpostop
Z400preop
ZOM
Q
ZOQ
Z400postop
J45preop
Km
J45postop
−4
−2
0
2
4
SHAP value (impact on model output)
High
Low
Feature value

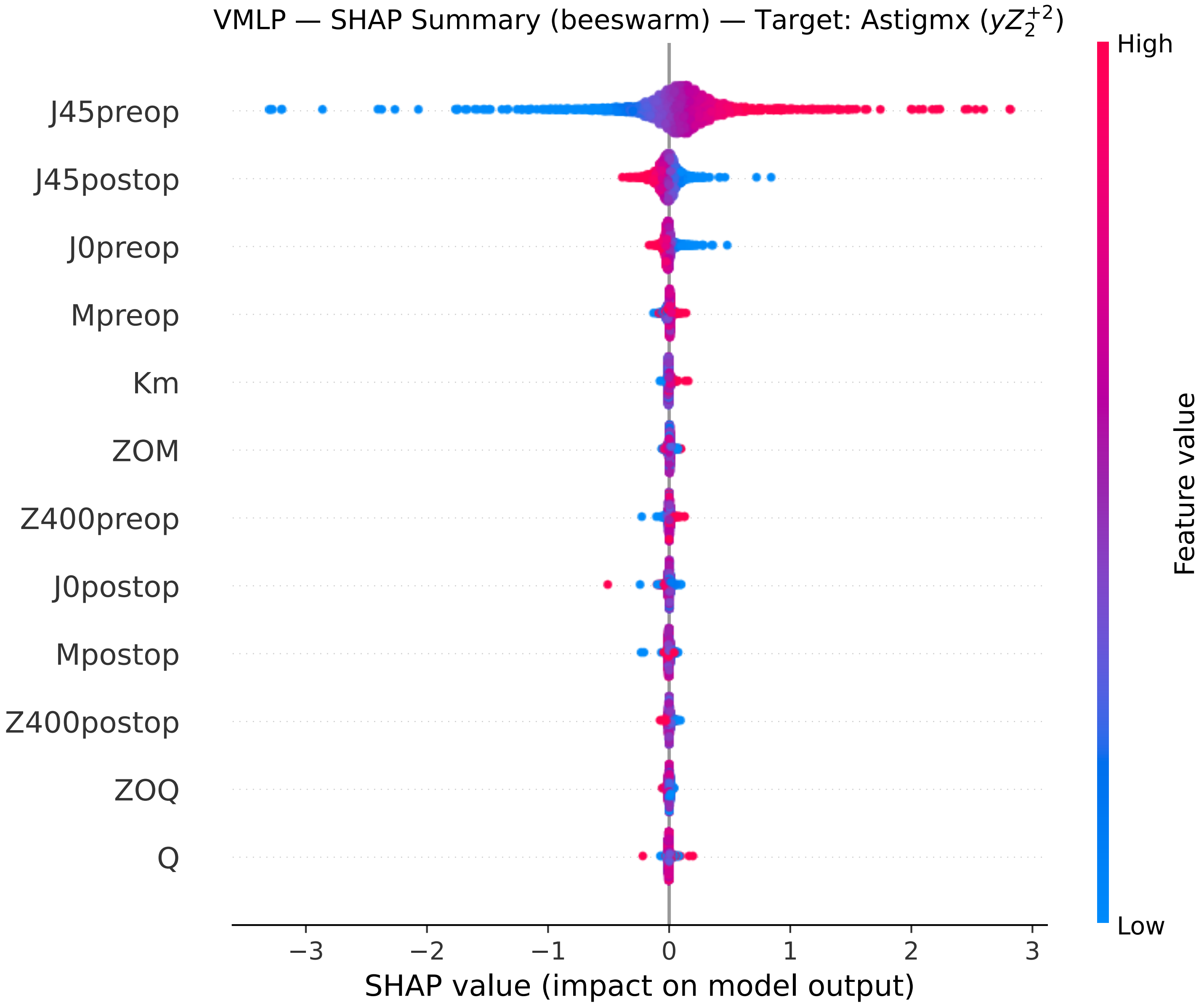
VMLP — SHAP Summary (beeswarm) — Target: Astigmx ($yZ_2^{+2}$)
J45preop
J45postop
J0preop
Mpreop
Km
ZOM
Z400preop
J0postop
Mpostop
Z400postop
ZOQ
Q
−3
−2
−1
0
1
2
3
SHAP value (impact on model output)
High
Low
Feature value

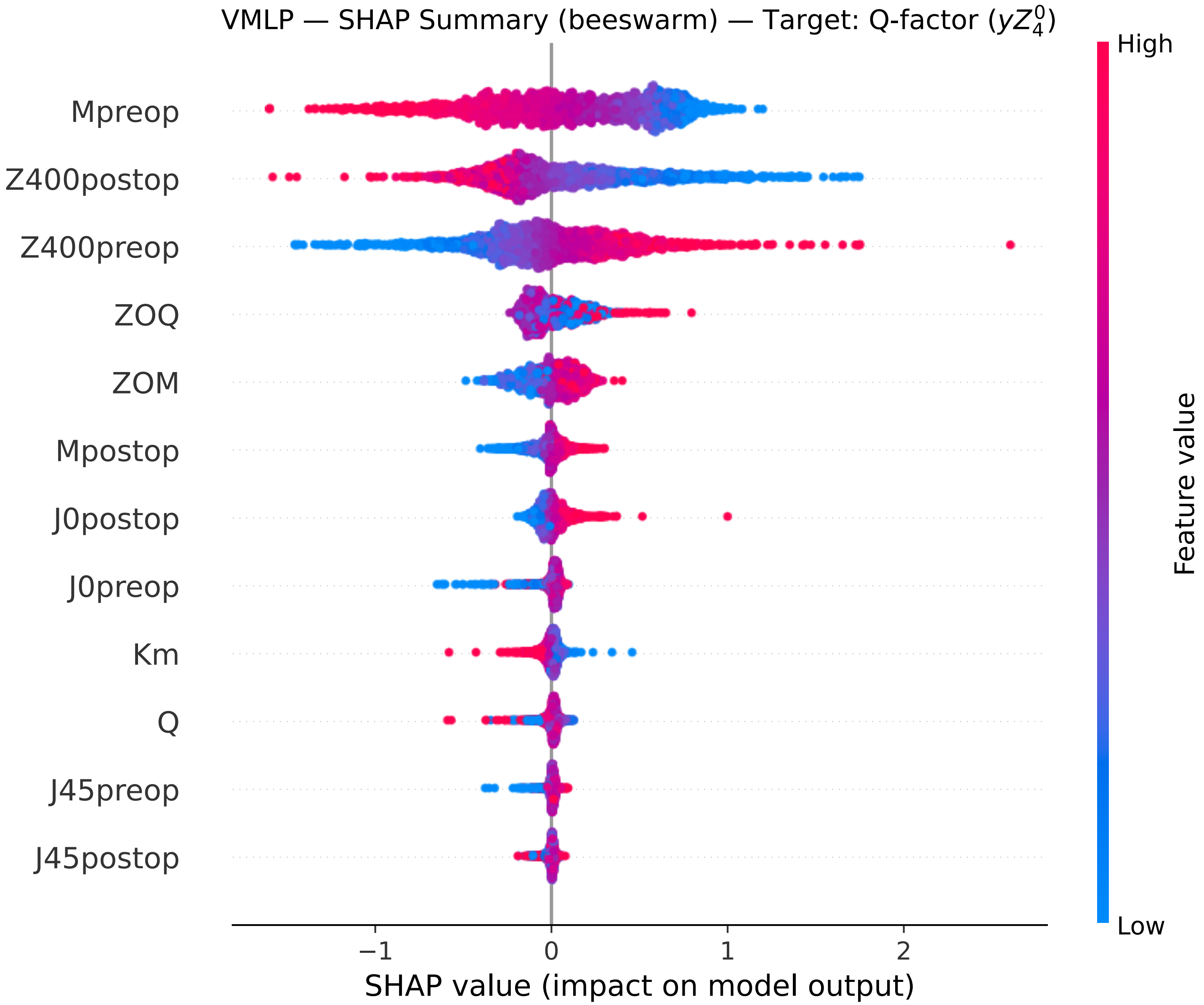
VMLP — SHAP Summary (beeswarm) — Target: Q-factor ($yZ_4^0$)
Mpreop
Z400postop
Z400preop
ZOQ
ZOM
Mpostop
J0postop
J0preop
Km
Q
J45preop
J45postop
−1
0
1
2
SHAP value (impact on model output)
High
Low
Feature value

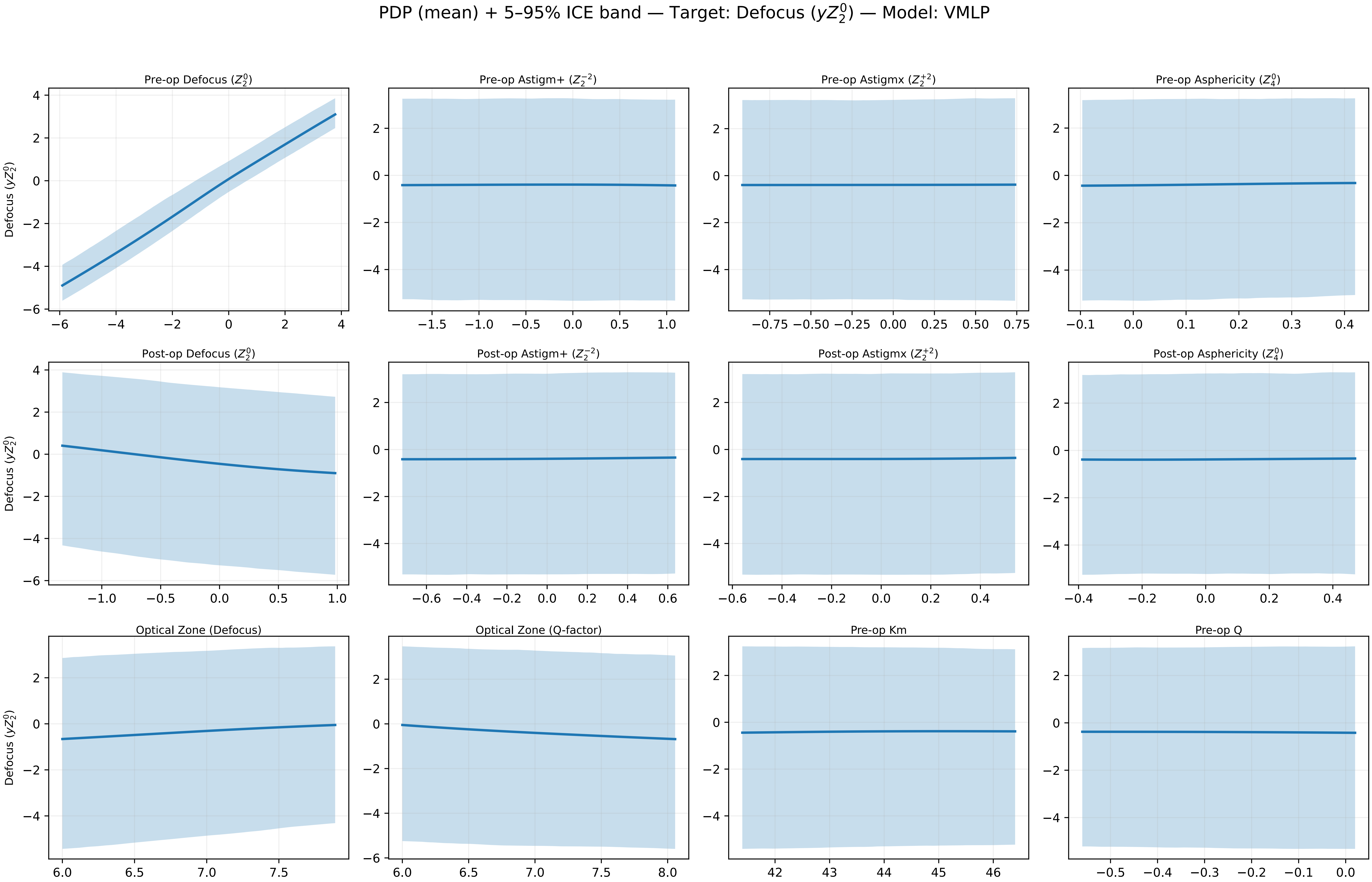
PDP (mean) + 5–95% ICE band — Target: Defocus ($yZ_2^0$) — Model: VMLP
Pre-op Defocus ($Z_2^0$)
Pre-op Astigm+ ($Z_2^{-2}$)
Pre-op Astigmx ($Z_2^{+2}$)
Pre-op Asphericity ($Z_4^0$)
Post-op Defocus ($Z_2^0$)
Post-op Astigm+ ($Z_2^{-2}$)
Post-op Astigmx ($Z_2^{+2}$)
Post-op Asphericity ($Z_4^0$)
Optical Zone (Defocus)
Optical Zone (Q-factor)
Pre-op Km
Pre-op Q
Defocus ($yZ_2^0$)

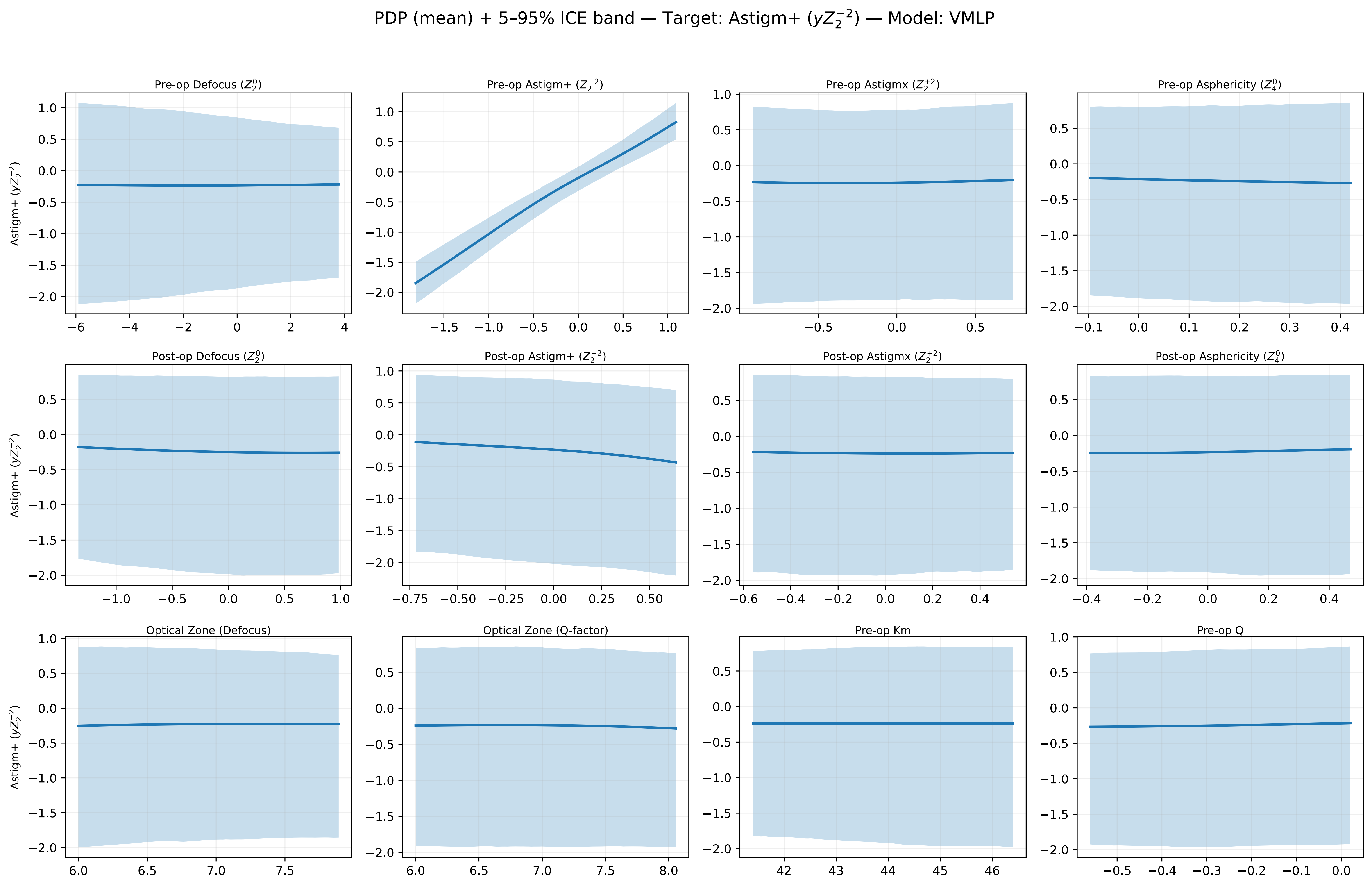
PDP (mean) + 5–95% ICE band — Target: Astigm+ ($yZ_2^{-2}$) — Model: VMLP
Pre-op Defocus ($Z_2^0$)
Pre-op Astigm+ ($Z_2^{-2}$)
Pre-op Astigmx ($Z_2^{+2}$)
Pre-op Asphericity ($Z_4^0$)
Post-op Defocus ($Z_2^0$)
Post-op Astigm+ ($Z_2^{-2}$)
Post-op Astigmx ($Z_2^{+2}$)
Post-op Asphericity ($Z_4^0$)
Optical Zone (Defocus)
Optical Zone (Q-factor)
Pre-op Km
Pre-op Q
Astigm+ ($yZ_2^{-2}$)

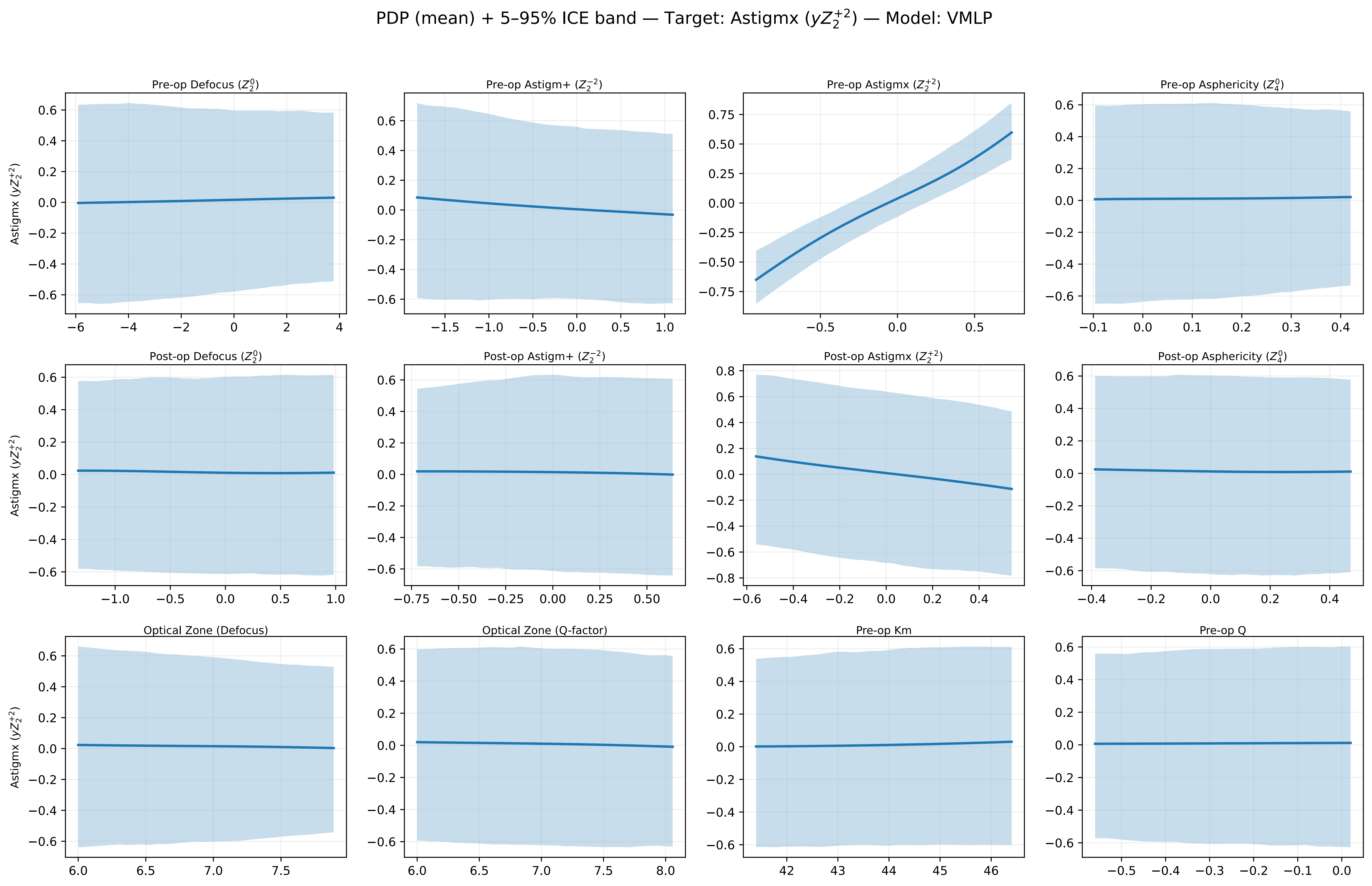

PDP (mean) + 5–95% ICE band — Target: Astigmx ($yZ_2^{+2}$) — Model: VMLP
Pre-op Defocus ($Z_2^0$)
Pre-op Astigm+ ($Z_2^{-2}$)
Pre-op Astigmx ($Z_2^{+2}$)
Pre-op Asphericity ($Z_4^0$)
Post-op Defocus ($Z_2^0$)
Post-op Astigm+ ($Z_2^{-2}$)
Post-op Astigmx ($Z_2^{+2}$)
Post-op Asphericity ($Z_4^0$)
Optical Zone (Defocus)
Optical Zone (Q-factor)
Pre-op Km
Pre-op Q
Astigmx ($yZ_2^{+2}$)

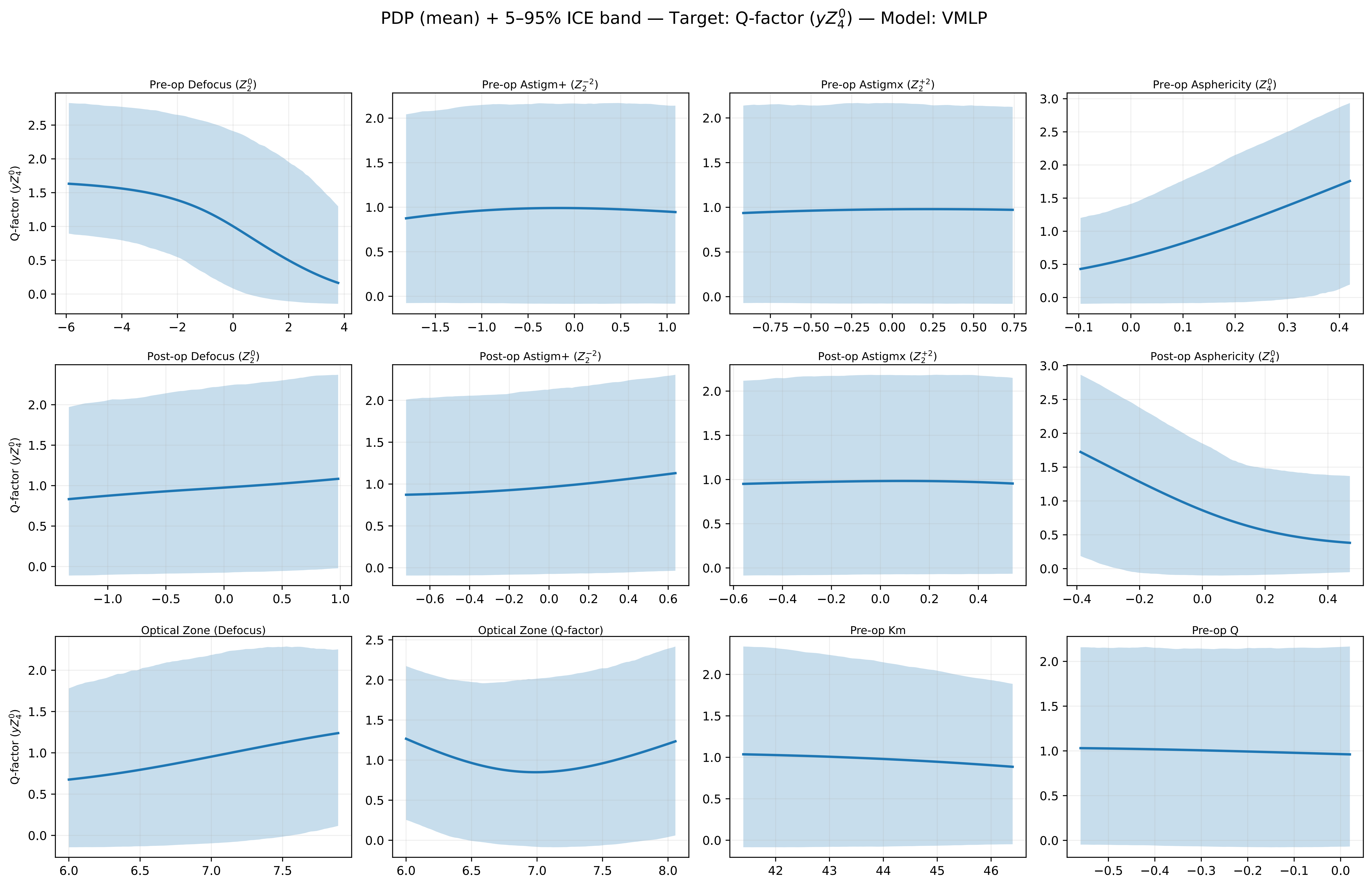
PDP (mean) + 5–95% ICE band — Target: Q-factor ($yZ_4^0$) — Model: VMLP
Pre-op Defocus ($Z_2^0$)
Pre-op Astigm+ ($Z_2^{-2}$)
Pre-op Astigmx ($Z_2^{+2}$)
Pre-op Asphericity ($Z_4^0$)
Post-op Defocus ($Z_2^0$)
Post-op Astigm+ ($Z_2^{-2}$)
Post-op Astigmx ($Z_2^{+2}$)
Post-op Asphericity ($Z_4^0$)
Optical Zone (Defocus)
Optical Zone (Q-factor)
Pre-op Km
Pre-op Q
Q-factor ($yZ_4^0$)

ICE curves (±centering: off) — Target: Defocus ($yZ_2^0$) — Model: VMLP

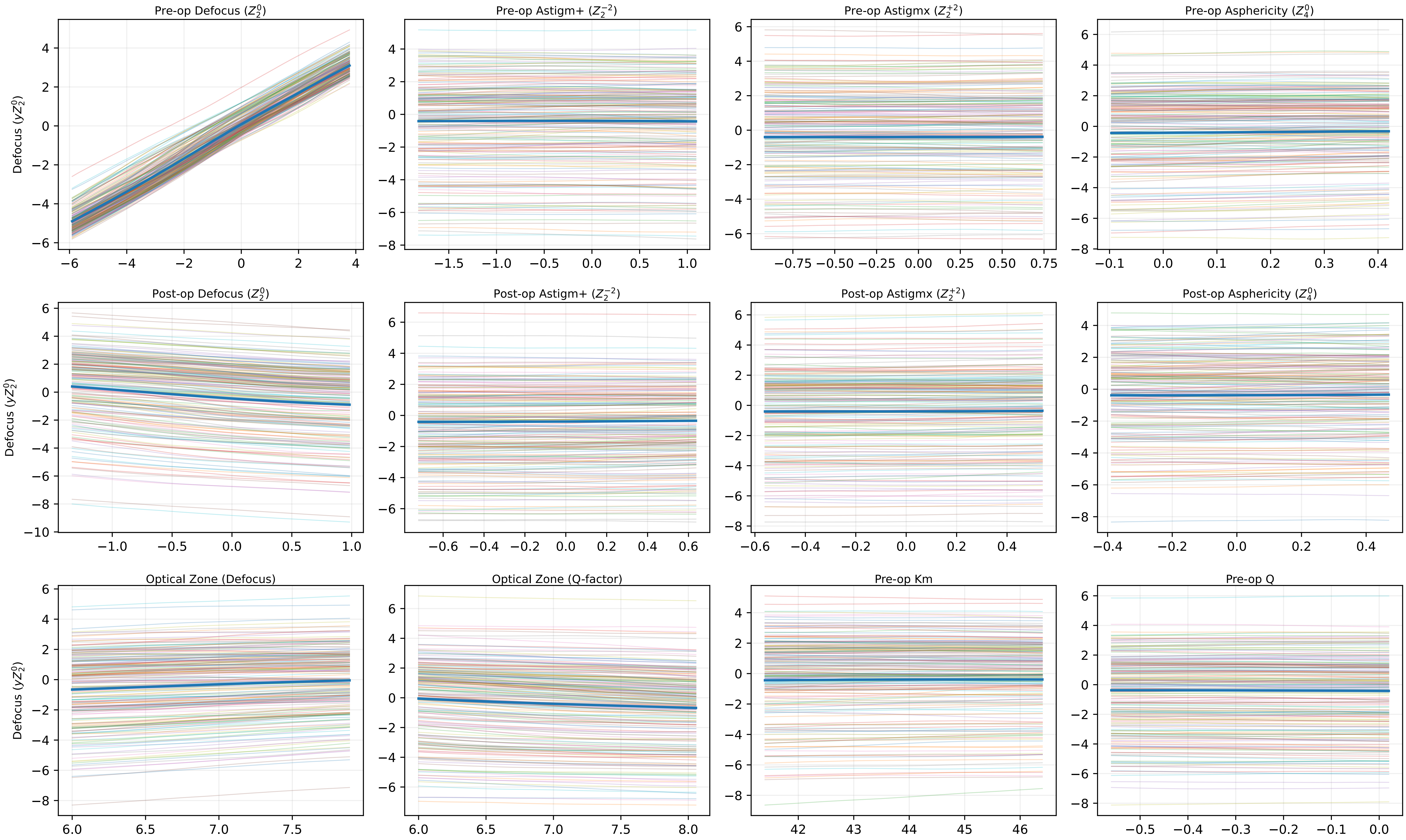

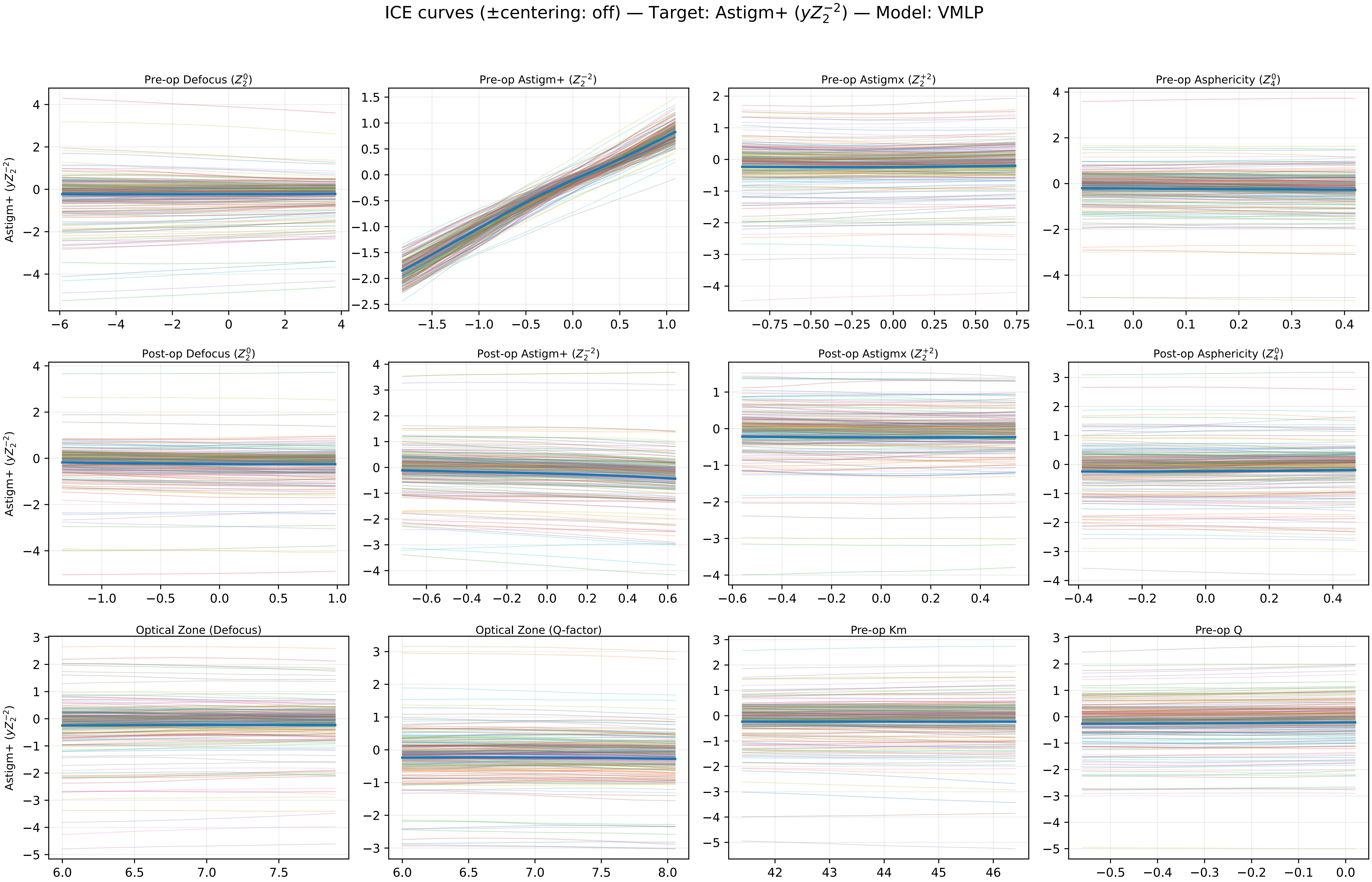

ICE curves (±centering: off) — Target: Astigm+ ($yZ_2^{-2}$) — Model: VMLP
Pre-op Defocus ($Z_2^0$)
Pre-op Astigm+ ($Z_2^{-2}$)
Pre-op Astigmx ($Z_2^{+2}$)
Pre-op Asphericity ($Z_4^0$)
Post-op Defocus ($Z_2^0$)
Post-op Astigm+ ($Z_2^{-2}$)
Post-op Astigmx ($Z_2^{+2}$)
Post-op Asphericity ($Z_4^0$)
Optical Zone (Defocus)
Optical Zone (Q-factor)
Pre-op Km
Pre-op Q
Astigm+ ($yZ_2^{-2}$)

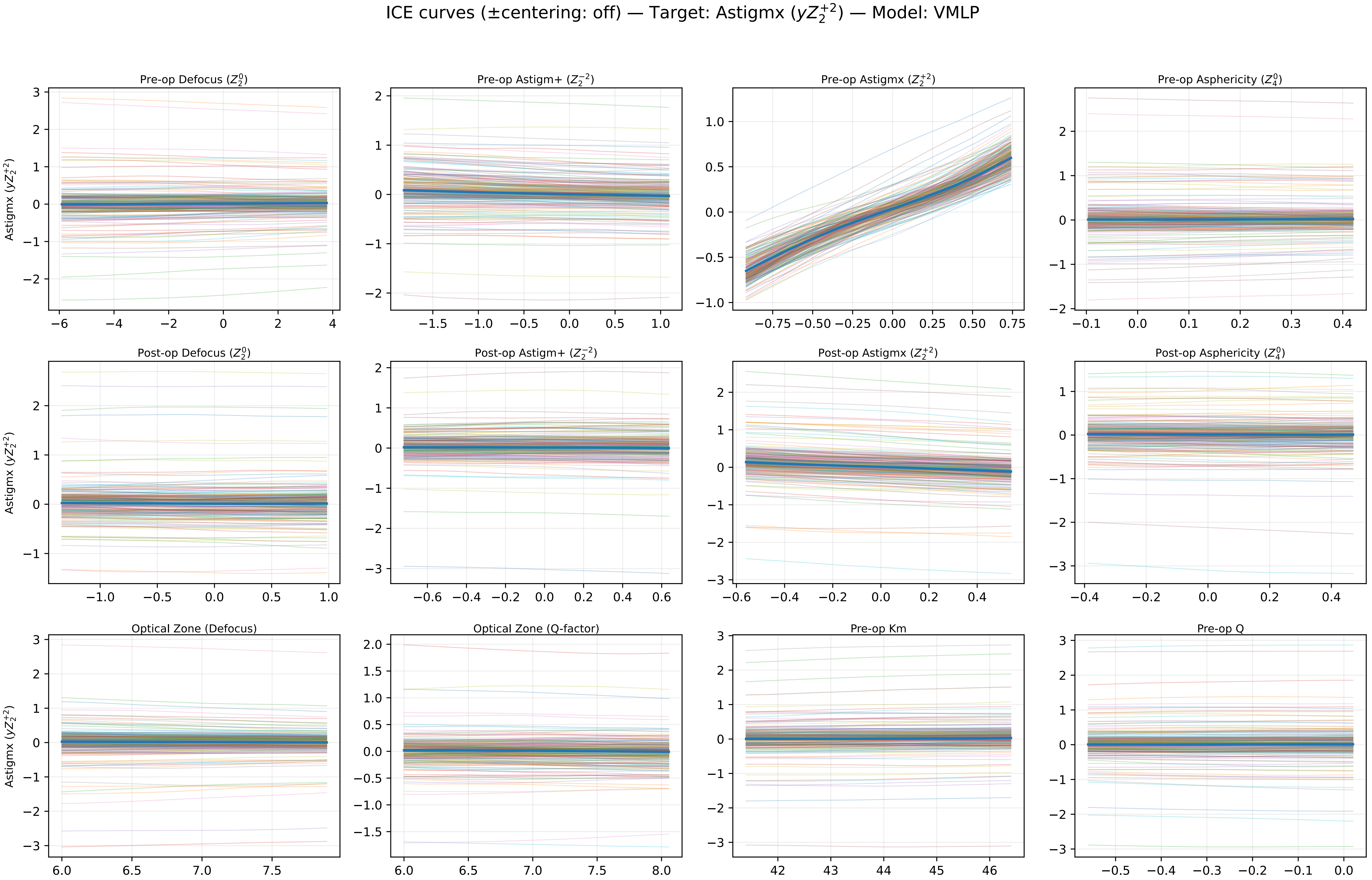
ICE curves (±centering: off) — Target: Astigmx ($yZ_2^{+2}$) — Model: VMLP
Pre-op Defocus ($Z_2^0$)
Pre-op Astigm+ ($Z_2^{-2}$)
Pre-op Astigmx ($Z_2^{+2}$)
Pre-op Asphericity ($Z_4^0$)
Post-op Defocus ($Z_2^0$)
Post-op Astigm+ ($Z_2^{-2}$)
Post-op Astigmx ($Z_2^{+2}$)
Post-op Asphericity ($Z_4^0$)
Optical Zone (Defocus)
Optical Zone (Q-factor)
Pre-op Km
Pre-op Q
Astigmx ($yZ_2^{+2}$)

ICE curves (±centering: off) — Target: Q-factor ($yZ_4^0$) — Model: VMLP

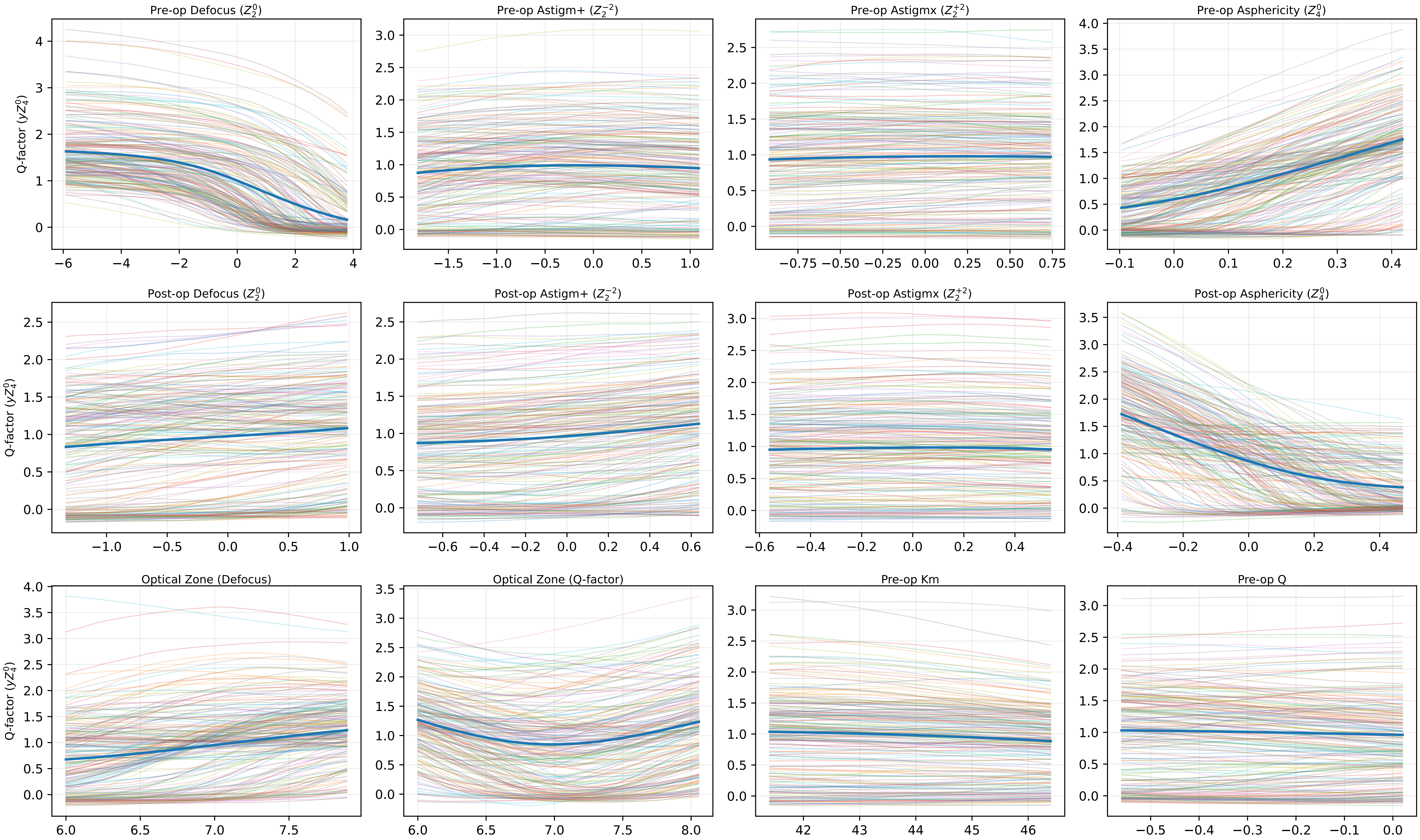

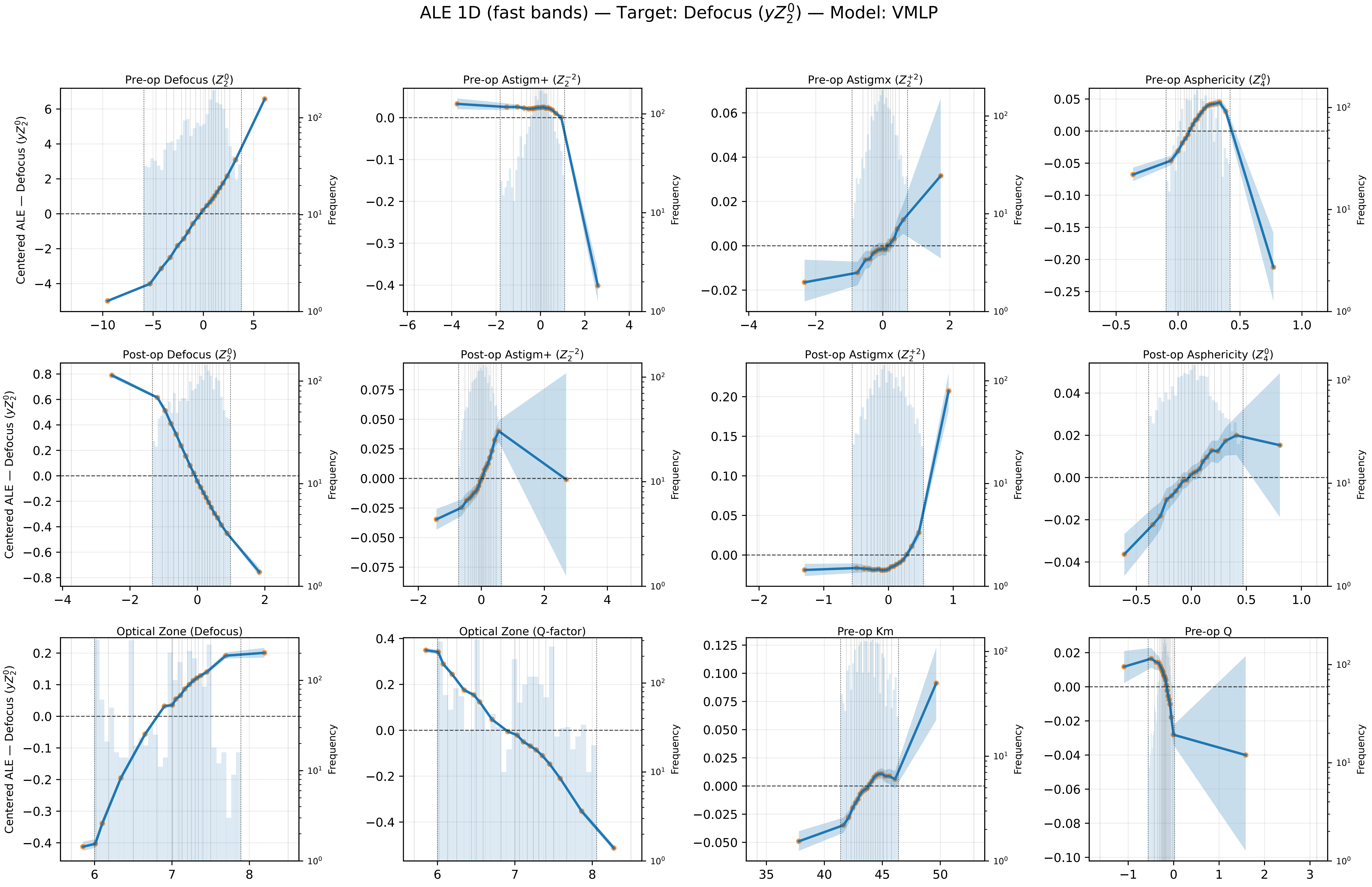
ALE 1D (fast bands) — Target: Defocus ($yZ_2^0$) — Model: VMLP
Pre-op Defocus ($Z_2^0$)
Pre-op Astigm+ ($Z_2^{-2}$)
Pre-op Astigmx ($Z_2^{+2}$)
Pre-op Asphericity ($Z_4^0$)
Post-op Defocus ($Z_2^0$)
Post-op Astigm+ ($Z_2^{-2}$)
Post-op Astigmx ($Z_2^{+2}$)
Post-op Asphericity ($Z_4^0$)
Optical Zone (Defocus)
Optical Zone (Q-factor)
Pre-op Km
Pre-op Q
Centered ALE — Defocus ($yZ_2^0$)
Frequency

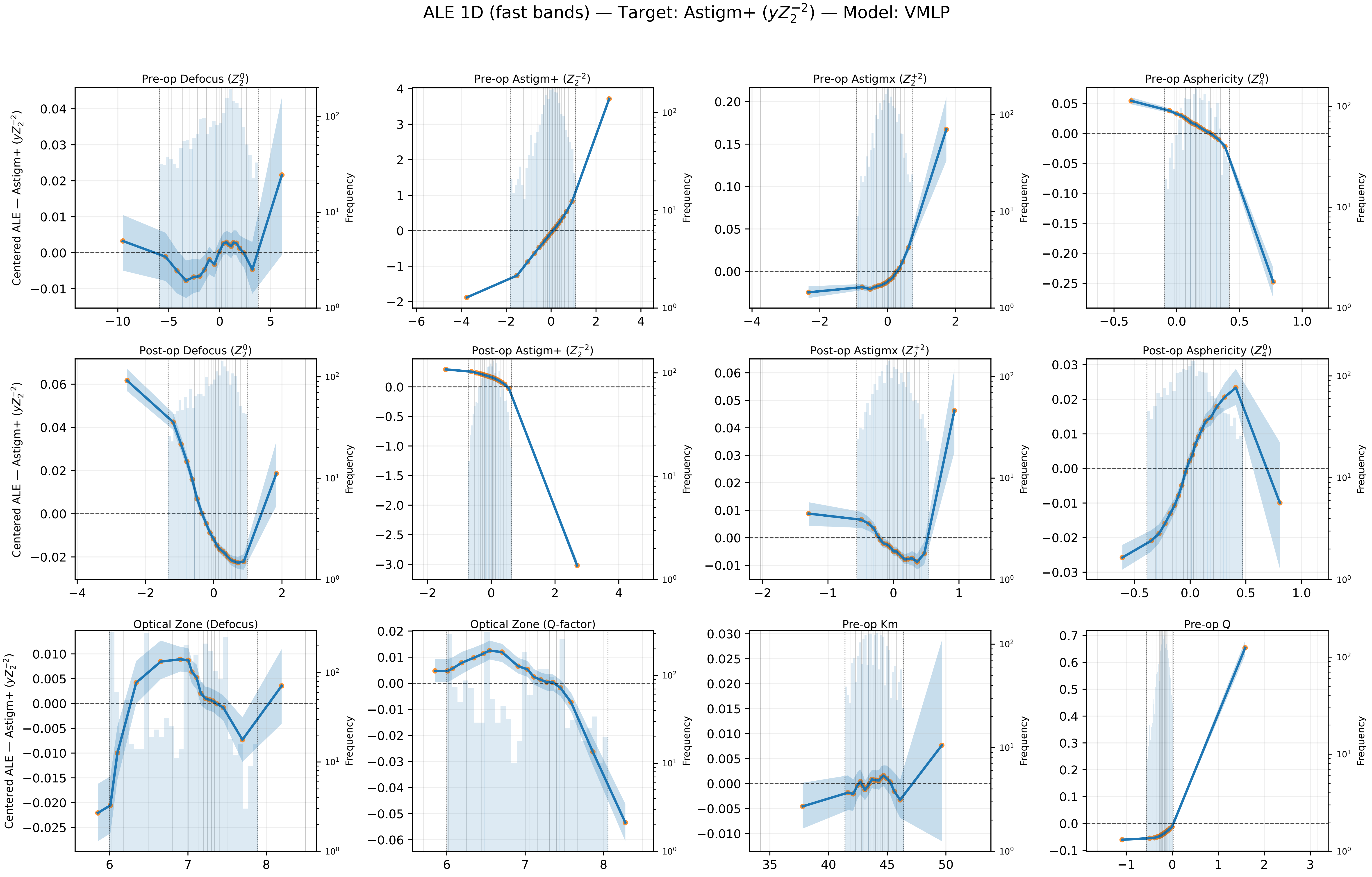

ALE 1D (fast bands) — Target: Astigm+ ($yZ_2^{-2}$) — Model: VMLP
Pre-op Defocus ($Z_2^0$)
Pre-op Astigm+ ($Z_2^{-2}$)
Pre-op Astigmx ($Z_2^{+2}$)
Pre-op Asphericity ($Z_4^0$)
Post-op Defocus ($Z_2^0$)
Post-op Astigm+ ($Z_2^{-2}$)
Post-op Astigmx ($Z_2^{+2}$)
Post-op Asphericity ($Z_4^0$)
Optical Zone (Defocus)
Optical Zone (Q-factor)
Pre-op Km
Pre-op Q
Centered ALE — Astigm+ ($yZ_2^{-2}$)
Frequency

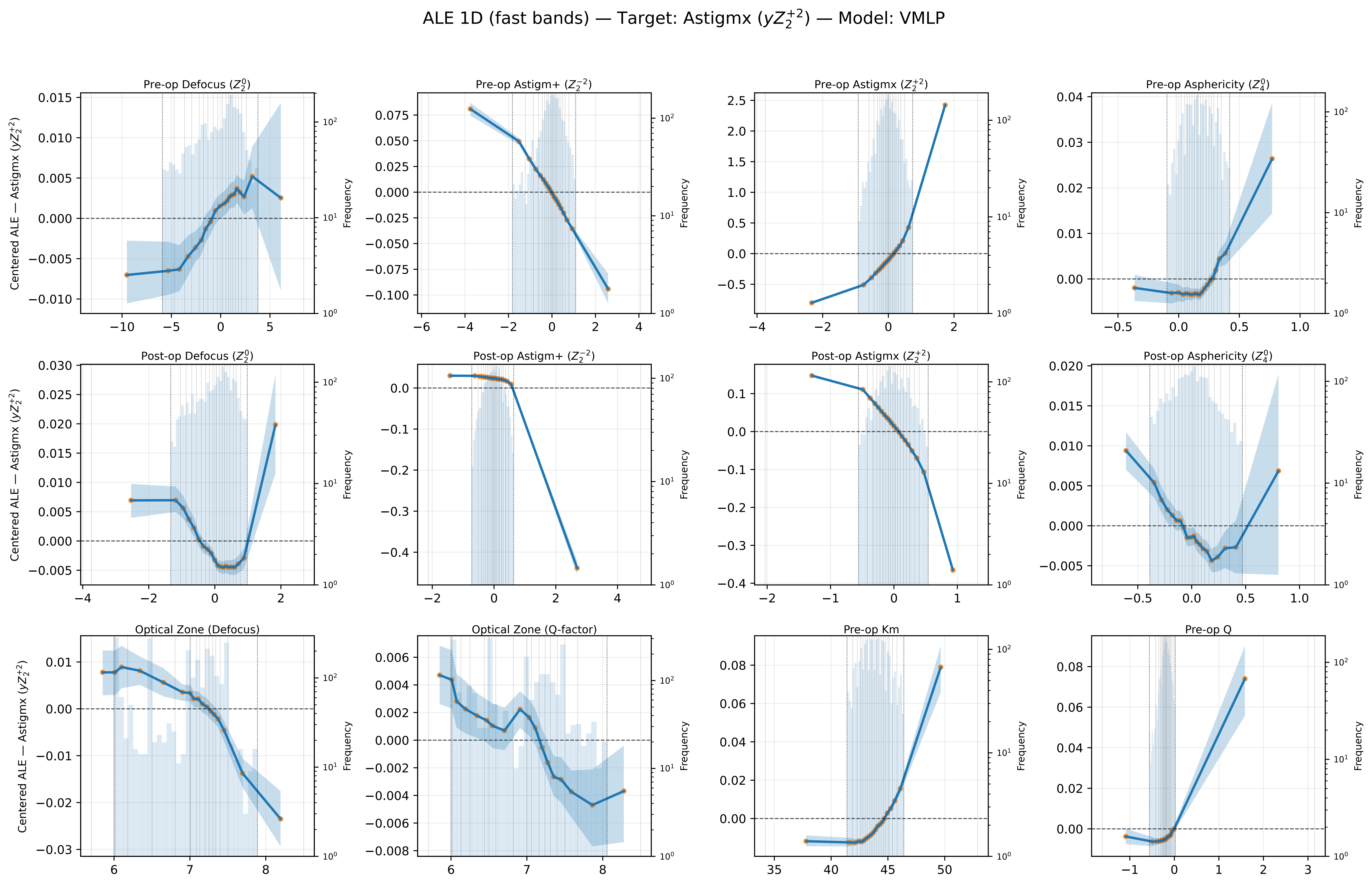

ALE 1D (fast bands) — Target: Astigmx ($yZ_2^{+2}$) — Model: VMLP
Pre-op Defocus ($Z_2^0$)
Pre-op Astigm+ ($Z_2^{-2}$)
Pre-op Astigmx ($Z_2^{+2}$)
Pre-op Asphericity ($Z_4^0$)
Post-op Defocus ($Z_2^0$)
Post-op Astigm+ ($Z_2^{-2}$)
Post-op Astigmx ($Z_2^{+2}$)
Post-op Asphericity ($Z_4^0$)
Optical Zone (Defocus)
Optical Zone (Q-factor)
Pre-op Km
Pre-op Q
Centered ALE — Astigmx ($yZ_2^{+2}$)
Frequency

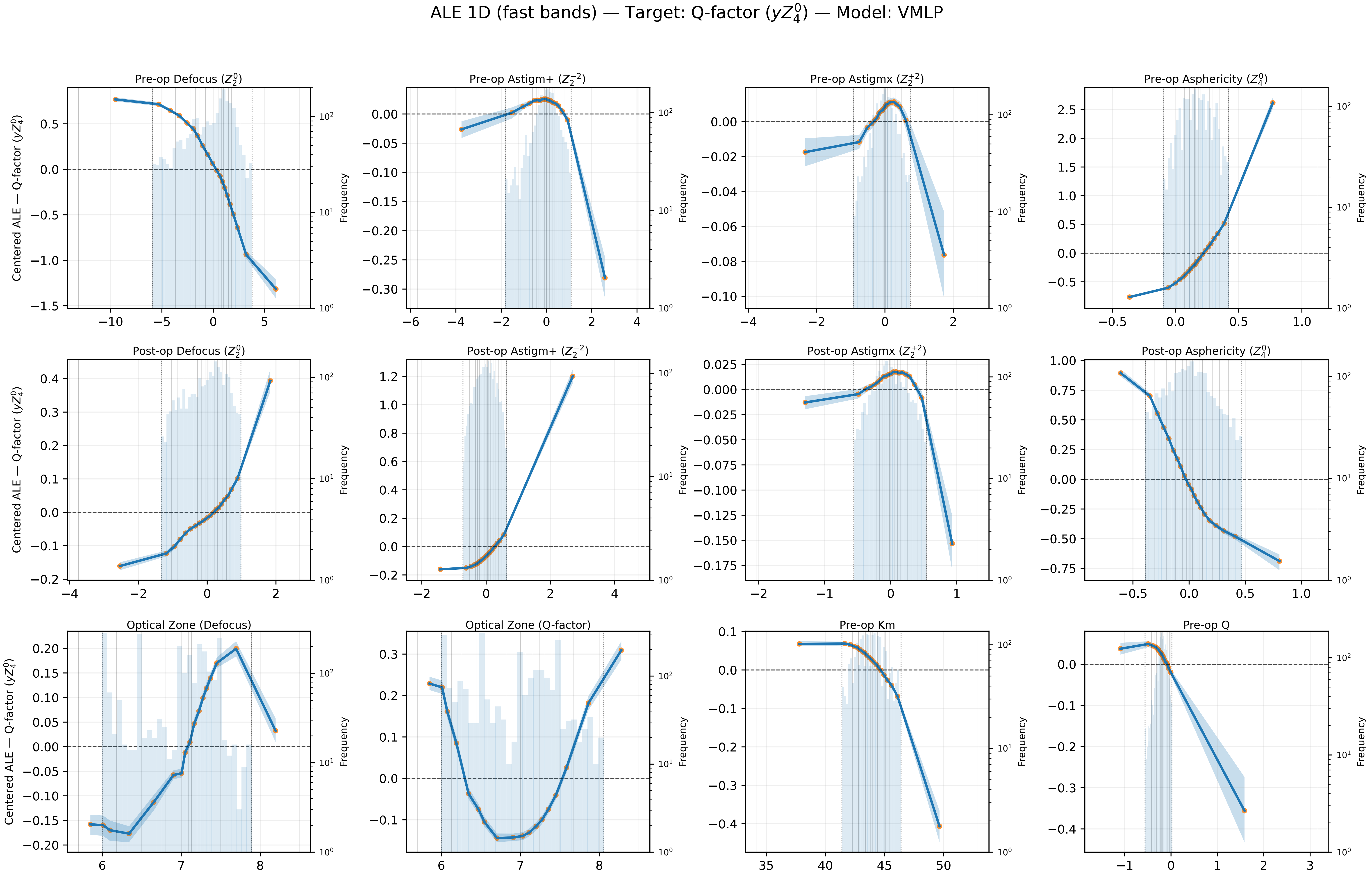
ALE 1D (fast bands) — Target: Q-factor ($yZ_4^0$) — Model: VMLP
Pre-op Defocus ($Z_2^0$)
Pre-op Astigm+ ($Z_2^{-2}$)
Pre-op Astigmx ($Z_2^{+2}$)
Pre-op Asphericity ($Z_4^0$)
Post-op Defocus ($Z_2^0$)
Post-op Astigm+ ($Z_2^{-2}$)
Post-op Astigmx ($Z_2^{+2}$)
Post-op Asphericity ($Z_4^0$)
Optical Zone (Defocus)
Optical Zone (Q-factor)
Pre-op Km
Pre-op Q
Centered ALE — Q-factor ($yZ_4^0$)
Frequency
