## Supplementary figures and images for "Machine Learning–based Prediction of LASIK Console Inputs for Aspheric Planning (Q-factor, Defocus, Astigmatism): A Translational Methods Study"

### Supplemental Notes 4

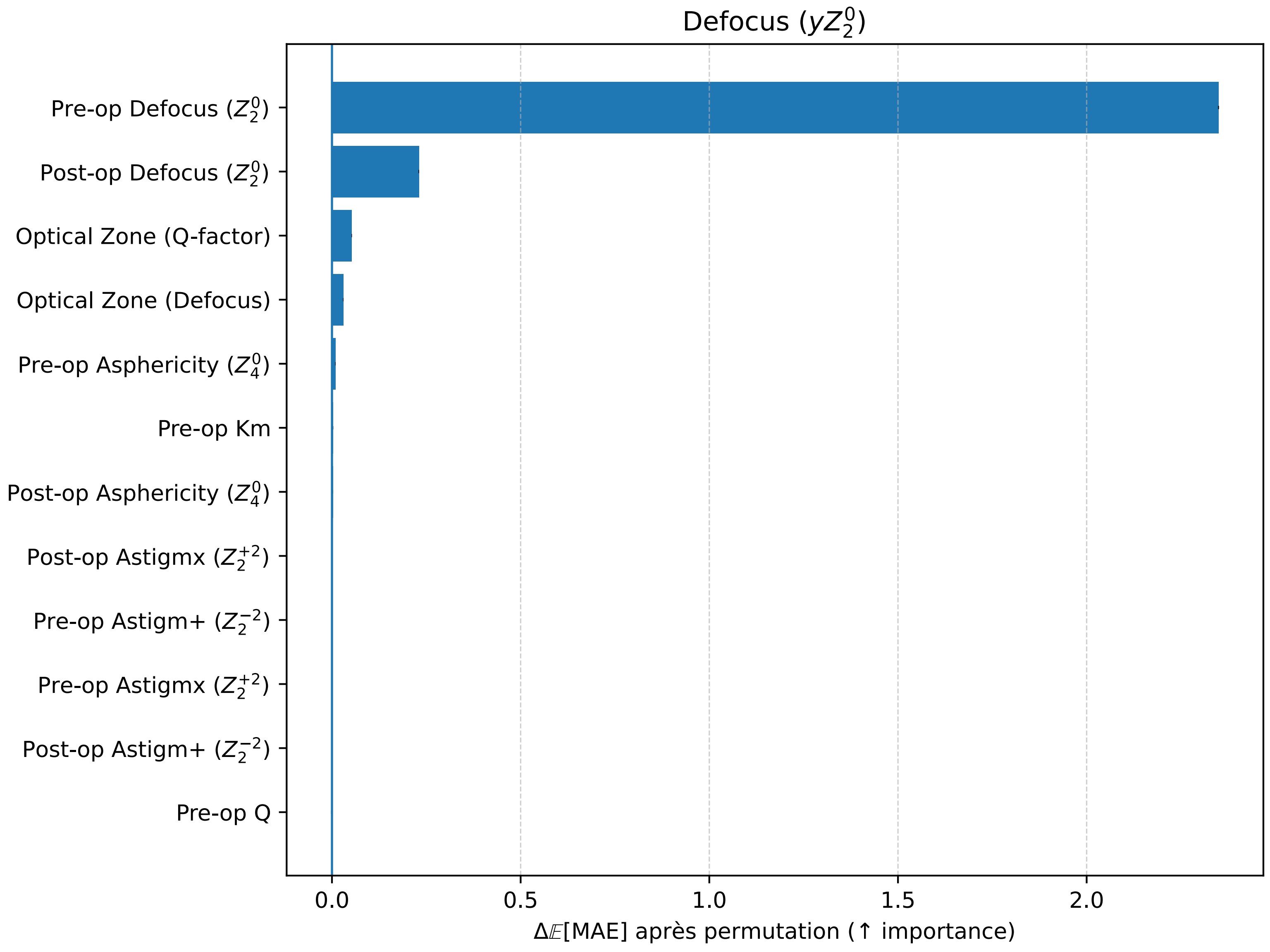

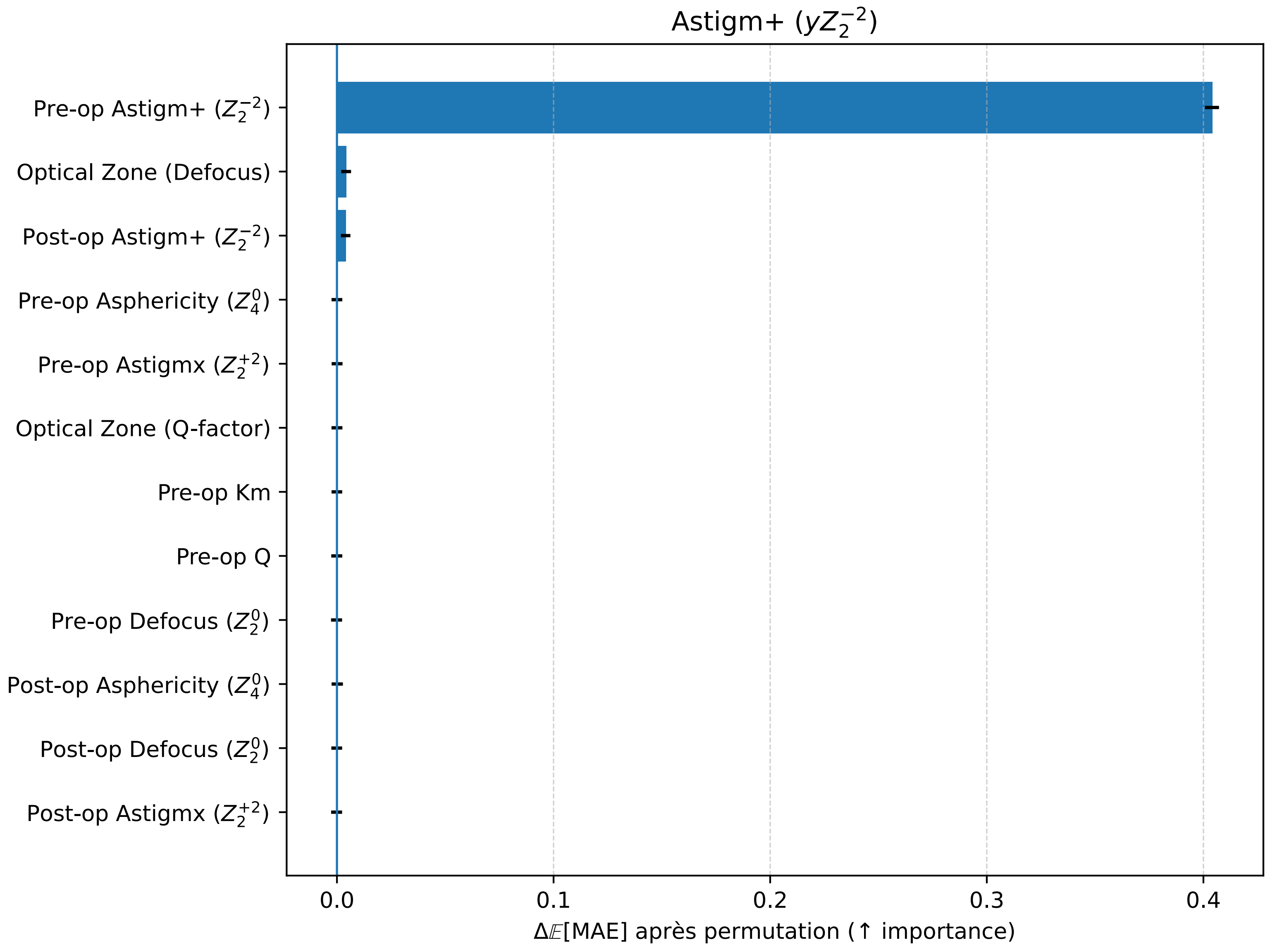

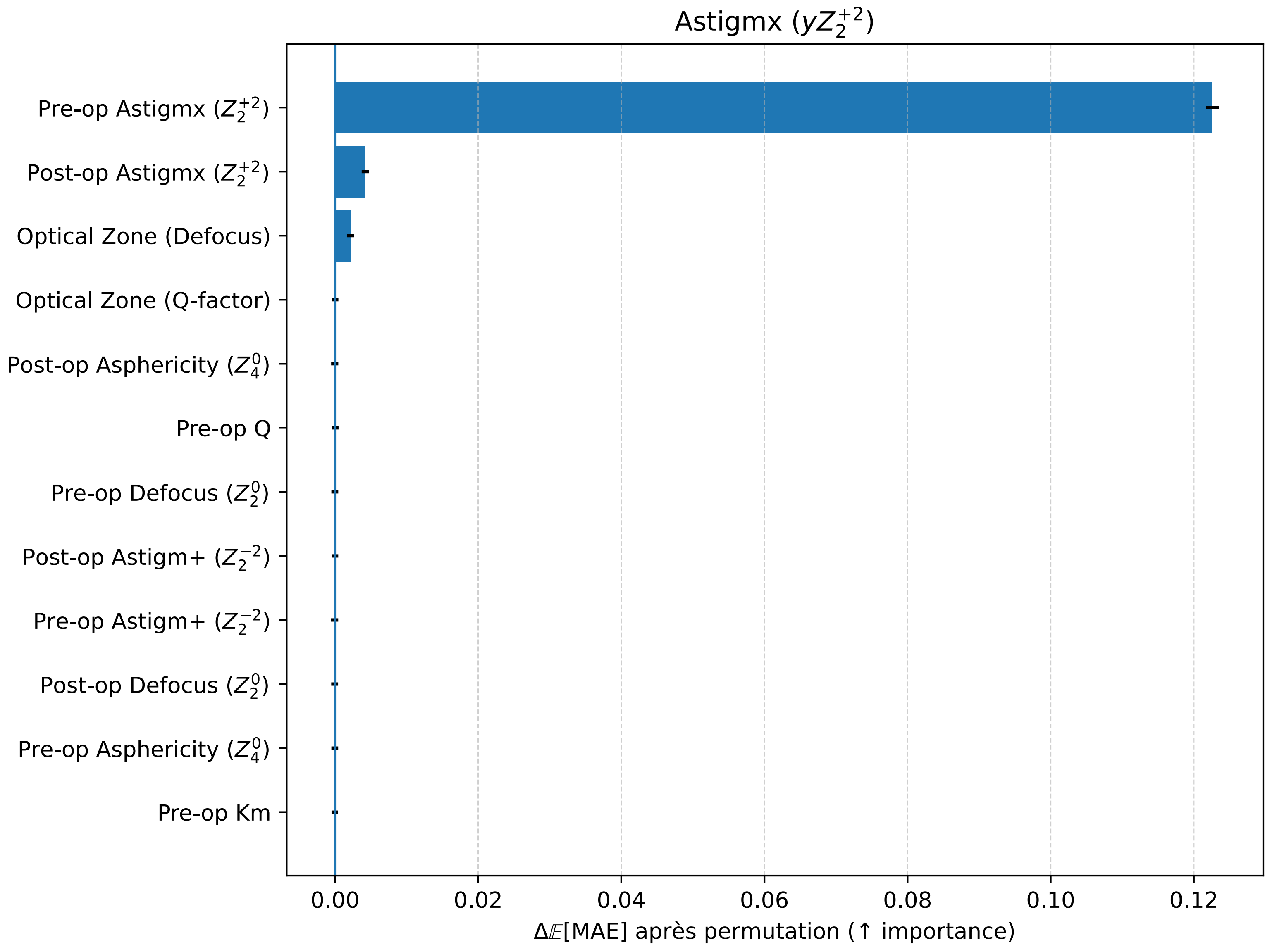

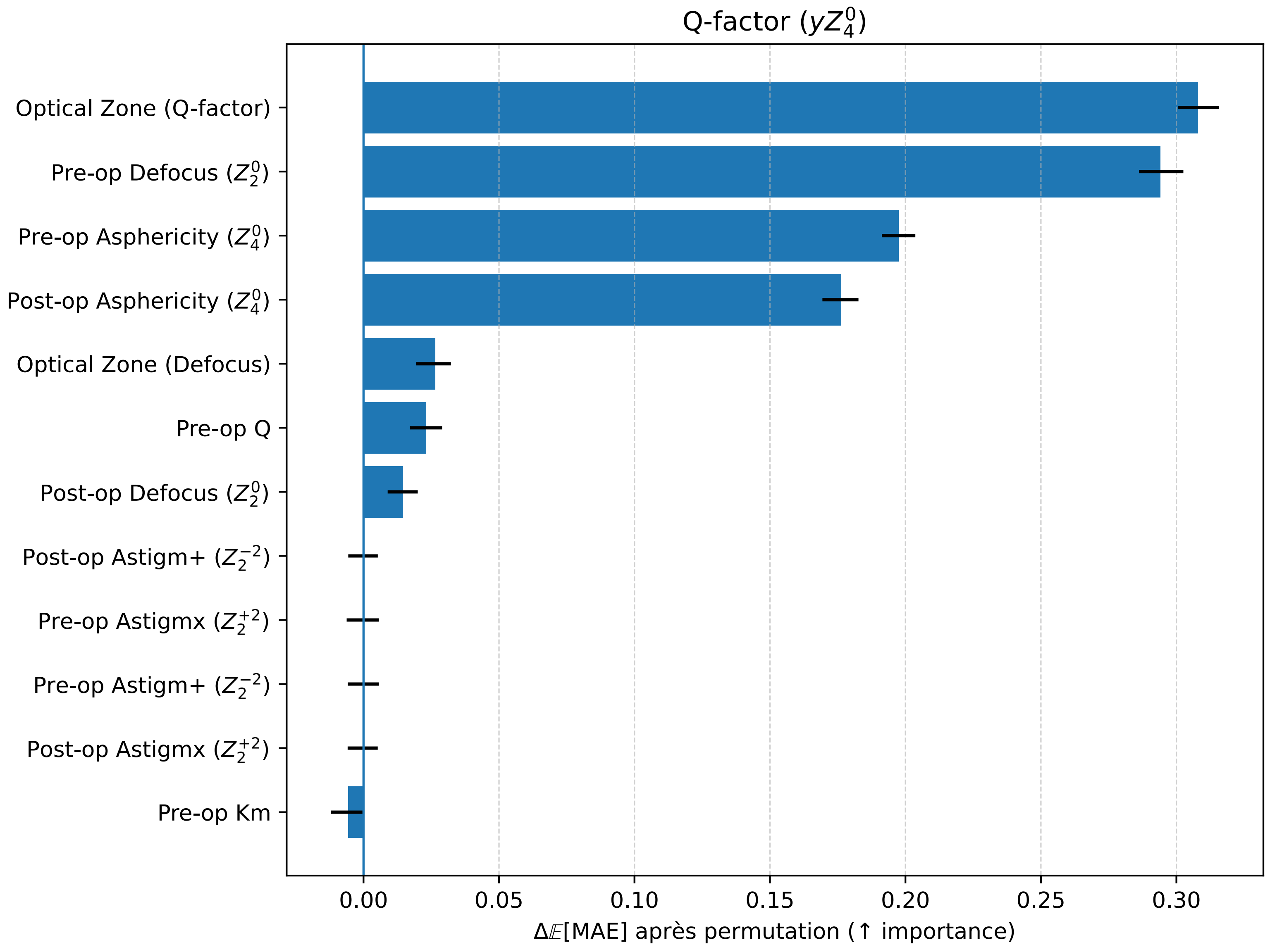
