## Supplemental Notes 5 for "Machine Learning–based Prediction of LASIK Console Inputs for Aspheric Planning (Q-factor, Defocus, Astigmatism): A Translational Methods Study"

### Sup. Note 5 — Correlation Report (8×8)

Supplementary Note 5 — Comprehensive Correlation & PCA (8×8)

Variables (order): Pre-op Defocus ($Z_2^0$), Post-op Defocus ($Z_2^0$), Pre-op Astigm+ ($Z_2^{-2}$), Post-op Astigm+ ($Z_2^{-2}$), P[illegible]

Decreasing academic importance:

(1) Bartlett’s sphericity (8×8), (2) 8×8 correlation heatmap,

(3) Pairwise correlations (r, Fisher 95% CI, Holm-adjusted p-values),

Interpretation guide:

- Bartlett $p<0.05$ ⇒ modes are not statistically orthogonal.
- Heatmap: magnitude/sign of r; blocks reveal common factors.
- Pairwise: |r| = effect size; CI excluding 0 + Holm<0.05 ⇒ robust link.

#### Global sphericity & top correlations

Bartlett (8×8): chi2=3090.03, df=28, p=0.000e+00 (n=2448)

Top-5 |r| (off-diagonal):

- Pre-op Defocus ($Z_2^0$) vs Post-op Asphericity ($Z_4^0$): r=-0.548
- Pre-op Astigmx ($Z_2^{+2}$) vs Post-op Astigmx ($Z_2^{+2}$): r=0.422
- Pre-op Asphericity ($Z_4^0$) vs Post-op Asphericity ($Z_4^0$): r=0.326
- Pre-op Astigm+ ($Z_2^{-2}$) vs Post-op Astigm+ ($Z_2^{-2}$): r=0.326
- Pre-op Defocus ($Z_2^0$) vs Post-op Astigm+ ($Z_2^{-2}$): r=0.209

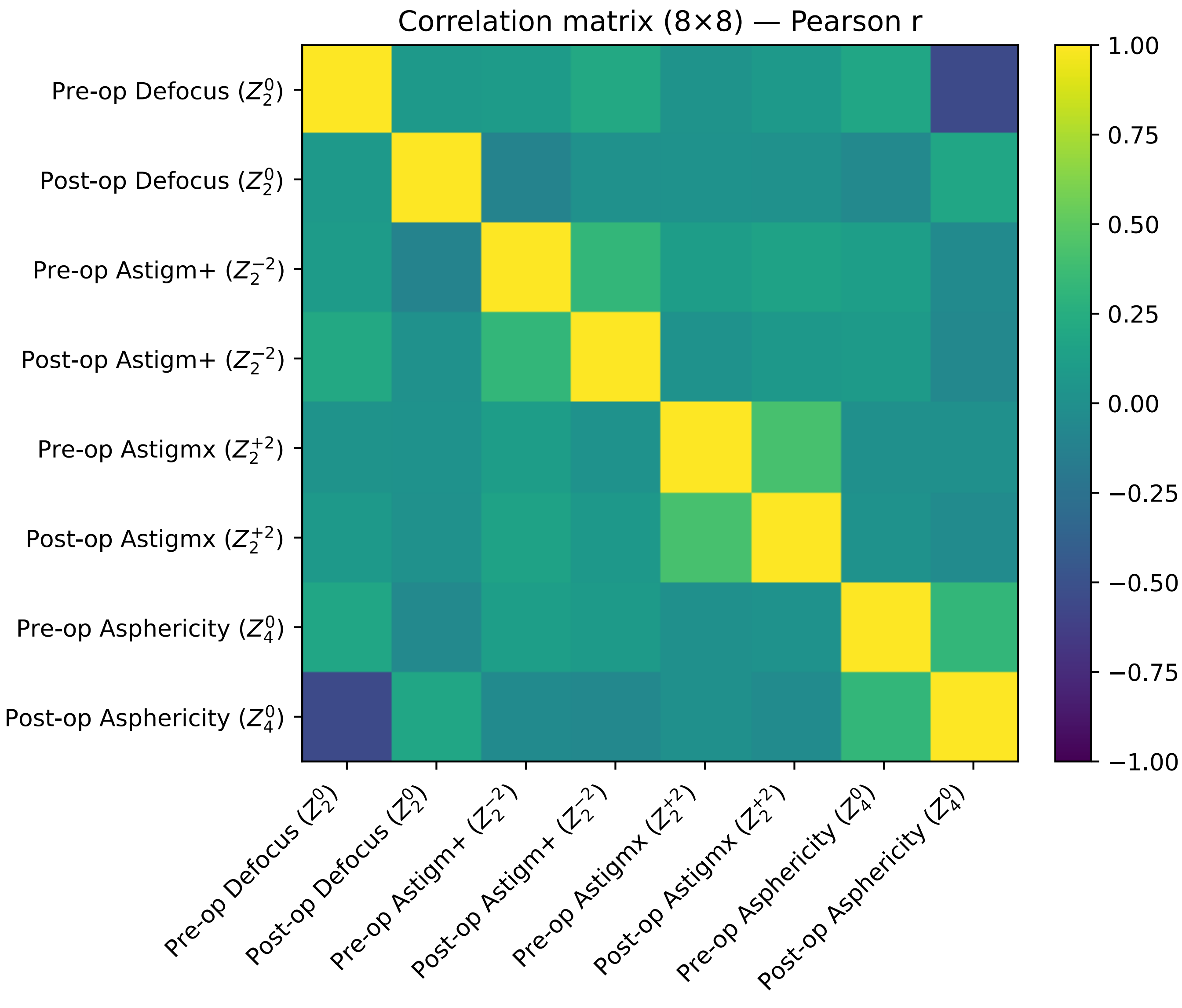

Correlation matrix (8×8) — Pearson r
Pre-op Defocus ($Z_2^0$)
Post-op Defocus ($Z_2^0$)
Pre-op Astigm+ ($Z_2^{-2}$)
Post-op Astigm+ ($Z_2^{-2}$)
Pre-op Astigmx ($Z_2^{+2}$)
Post-op Astigmx ($Z_2^{+2}$)
Pre-op Asphericity ($Z_4^0$)
Post-op Asphericity ($Z_4^0$)
1.00
0.75
0.50
0.25
0.00
−0.25
−0.50
−0.75
−1.00

### Pairwise correlations (r, CI95, Holm p) — 28 pairs (rows 1–28 of 28)

| label_i | label_j | r | CI95_low | CI95_high | p_val | p_val_holm |
|---|---|---|---|---|---|---|
| Post-op Astigm+ ($Z_2^{-2}$) | Post-op Astigmx ($Z_2^{+2}$) | 0.065 | 0.025 | 0.104 | 0.001 | 0.017 |
| Post-op Astigm+ ($Z_2^{-2}$) | Pre-op Astigmx ($Z_2^{+2}$) | 0.017 | -0.023 | 0.056 | 0.410 | 1.000 |
| Post-op Astigm+ ($Z_2^{-2}$) | Post-op Asphericity ($Z_4^{0}$) | -0.068 | -0.108 | -0.029 | 0.001 | 0.009 |
| Post-op Astigm+ ($Z_2^{-2}$) | Pre-op Asphericity ($Z_4^{0}$) | 0.087 | 0.048 | 0.126 | 0.000 | 0.000 |
| Pre-op Astigm+ ($Z_2^{-2}$) | Post-op Astigm+ ($Z_2^{-2}$) | 0.326 | 0.290 | 0.361 | 0.000 | 0.000 |
| Pre-op Astigm+ ($Z_2^{-2}$) | Post-op Astigmx ($Z_2^{+2}$) | 0.149 | 0.110 | 0.188 | 0.000 | 0.000 |
| Pre-op Astigm+ ($Z_2^{-2}$) | Pre-op Astigmx ($Z_2^{+2}$) | 0.110 | 0.071 | 0.149 | 0.000 | 0.000 |
| Pre-op Astigm+ ($Z_2^{-2}$) | Post-op Asphericity ($Z_4^{0}$) | -0.043 | -0.083 | -0.004 | 0.033 | 0.329 |
| Pre-op Astigm+ ($Z_2^{-2}$) | Pre-op Asphericity ($Z_4^{0}$) | 0.122 | 0.083 | 0.161 | 0.000 | 0.000 |
| Post-op Astigmx ($Z_2^{+2}$) | Post-op Asphericity ($Z_4^{0}$) | -0.034 | -0.073 | 0.006 | 0.094 | 0.848 |
| Post-op Astigmx ($Z_2^{+2}$) | Pre-op Asphericity ($Z_4^{0}$) | 0.020 | -0.019 | 0.060 | 0.316 | 1.000 |
| Pre-op Astigmx ($Z_2^{+2}$) | Post-op Astigmx ($Z_2^{+2}$) | 0.422 | 0.388 | 0.454 | 0.000 | 0.000 |
| Pre-op Astigmx ($Z_2^{+2}$) | Post-op Asphericity ($Z_4^{0}$) | 0.006 | -0.033 | 0.046 | 0.759 | 1.000 |
| Pre-op Astigmx ($Z_2^{+2}$) | Pre-op Asphericity ($Z_4^{0}$) | 0.002 | -0.037 | 0.042 | 0.915 | 0.915 |
| Post-op Defocus ($Z_2^{0}$) | Post-op Astigm+ ($Z_2^{-2}$) | 0.010 | -0.030 | 0.049 | 0.626 | 1.000 |
| Post-op Defocus ($Z_2^{0}$) | Pre-op Astigm+ ($Z_2^{-2}$) | -0.102 | -0.141 | -0.063 | 0.000 | 0.000 |
| Post-op Defocus ($Z_2^{0}$) | Post-op Astigmx ($Z_2^{+2}$) | 0.012 | -0.028 | 0.051 | 0.563 | 1.000 |
| Post-op Defocus ($Z_2^{0}$) | Pre-op Astigmx ($Z_2^{+2}$) | 0.020 | -0.020 | 0.059 | 0.328 | 1.000 |
| Post-op Defocus ($Z_2^{0}$) | Post-op Asphericity ($Z_4^{0}$) | 0.181 | 0.142 | 0.219 | 0.000 | 0.000 |
| Post-op Defocus ($Z_2^{0}$) | Pre-op Asphericity ($Z_4^{0}$) | -0.055 | -0.095 | -0.016 | 0.006 | 0.067 |
| Pre-op Defocus ($Z_2^{0}$) | Post-op Astigm+ ($Z_2^{-2}$) | 0.209 | 0.171 | 0.247 | 0.000 | 0.000 |
| Pre-op Defocus ($Z_2^{0}$) | Pre-op Astigm+ ($Z_2^{-2}$) | 0.100 | 0.061 | 0.139 | 0.000 | 0.000 |
| Pre-op Defocus ($Z_2^{0}$) | Post-op Astigmx ($Z_2^{+2}$) | 0.080 | 0.040 | 0.119 | 0.000 | 0.001 |
| Pre-op Defocus ($Z_2^{0}$) | Pre-op Astigmx ($Z_2^{+2}$) | 0.025 | -0.015 | 0.064 | 0.223 | 1.000 |
| Pre-op Defocus ($Z_2^{0}$) | Post-op Defocus ($Z_2^{0}$) | 0.081 | 0.041 | 0.120 | 0.000 | 0.001 |
| Pre-op Defocus ($Z_2^{0}$) | Post-op Asphericity ($Z_4^{0}$) | -0.548 | -0.575 | -0.519 | 0.000 | 0.000 |
| Pre-op Defocus ($Z_2^{0}$) | Pre-op Asphericity ($Z_4^{0}$) | 0.182 | 0.143 | 0.220 | 0.000 | 0.000 |
| Pre-op Asphericity ($Z_4^{0}$) | Post-op Asphericity ($Z_4^{0}$) | 0.326 | 0.290 | 0.361 | 0.000 | 0.000 |

### Sup. Note 5 — Correlation Report (8×8)

Supplementary Note 5 — Correlation & PCA (8×8) on testset (n=147)

Variables (order): Pre-op Defocus ($Z_2^0$), Post-op Defocus ($Z_2^0$), Pre-op Astigm+ ($Z_2^{-2}$), Post-op Astigm+ ($Z_2^{-2}$), P

Decreasing academic importance:

(1) Bartlett's sphericity (8×8), (2) 8×8 correlation heatmap,

(3) Pairwise correlations (r, Fisher 95% CI, Holm-adjusted p-values),

Interpretation guide:

- Bartlett p<0.05 ⇒ modes are not statistically orthogonal.
- Heatmap: magnitude/sign of r; blocks reveal common factors.
- Pairwise: |r| = effect size; CI excluding 0 + Holm<0.05 ⇒ robust link.

#### Global sphericity & top correlations

Bartlett (8×8): chi2=244.45, df=28, p=0.000e+00 (n=147)

Top-5 |r|:

- Pre-op Defocus ($Z_2^0$) vs Post-op Asphericity ($Z_4^0$): r=-0.644
- Pre-op Asphericity ($Z_4^0$) vs Post-op Asphericity ($Z_4^0$): r=0.401
- Pre-op Astigmx ($Z_2^{+2}$) vs Post-op Astigmx ($Z_2^{+2}$): r=0.267
- Post-op Defocus ($Z_2^0$) vs Post-op Asphericity ($Z_4^0$): r=0.249
- Post-op Defocus ($Z_2^0$) vs Pre-op Astigm+ ($Z_2^{-2}$): r=-0.224

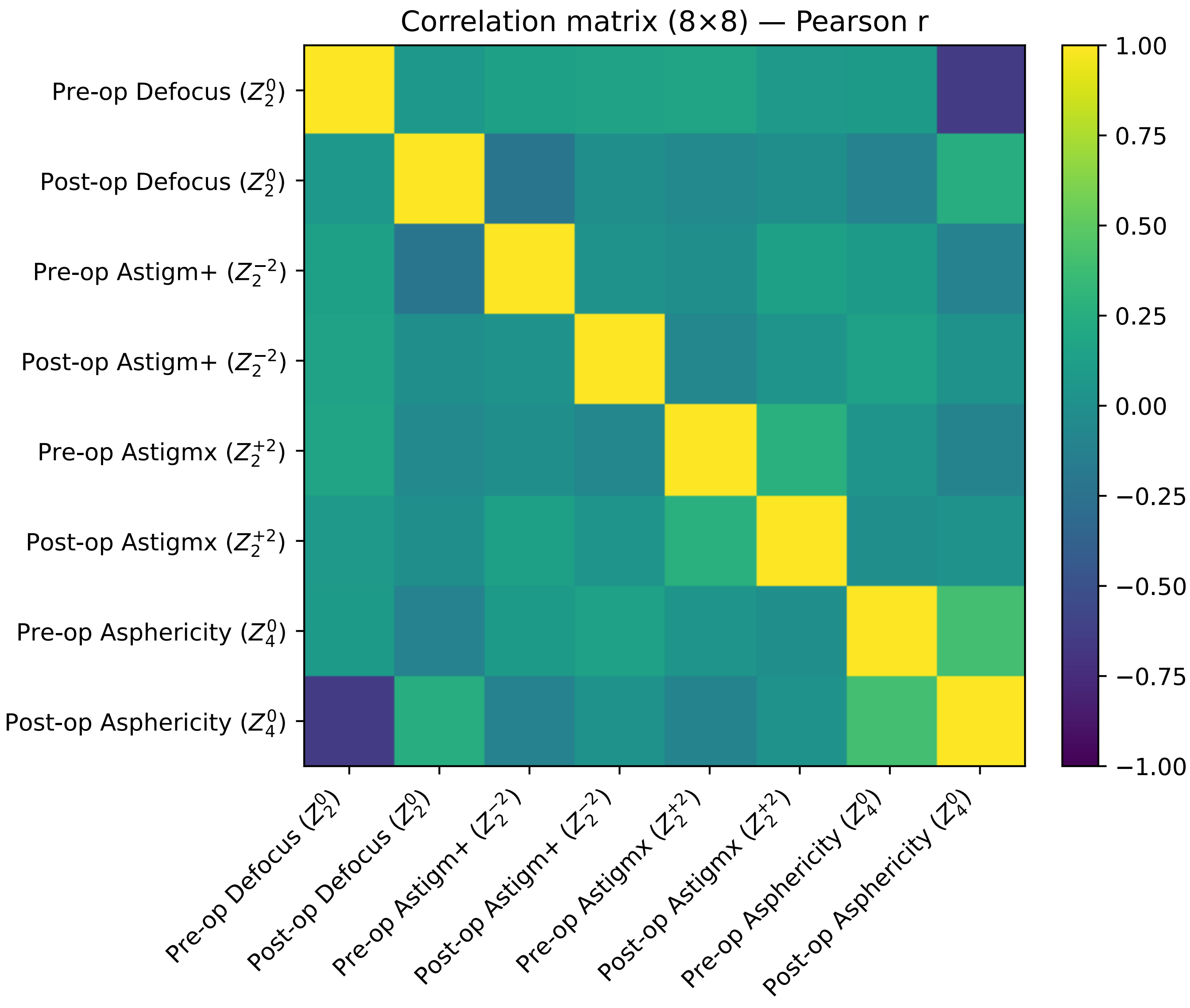

Correlation matrix (8×8) — Pearson r
Pre-op Defocus ($Z_2^0$)
Post-op Defocus ($Z_2^0$)
Pre-op Astigm+ ($Z_2^{-2}$)
Post-op Astigm+ ($Z_2^{-2}$)
Pre-op Astigmx ($Z_2^{+2}$)
Post-op Astigmx ($Z_2^{+2}$)
Pre-op Asphericity ($Z_4^0$)
Post-op Asphericity ($Z_4^0$)
1.00
0.75
0.50
0.25
0.00
−0.25
−0.50
−0.75
−1.00

#### Pairwise correlations — testset146.csv (rows 1–28 of 28)

| label_i | label_j | r | CI95_low | CI95_high | p_val | p_val_holm |
|---|---|---|---|---|---|---|
| Post-op Astigm+ ($Z_2^{-2}$) | Post-op Astigmx ($Z_2^{+2}$) | 0.031 | -0.131 | 0.192 | 0.706 | 1.000 |
| Post-op Astigm+ ($Z_2^{-2}$) | Pre-op Astigmx ($Z_2^{+2}$) | -0.074 | -0.233 | 0.089 | 0.375 | 1.000 |
| Post-op Astigm+ ($Z_2^{-2}$) | Post-op Asphericity ($Z_4^{0}$) | 0.022 | -0.141 | 0.183 | 0.793 | 1.000 |
| Post-op Astigm+ ($Z_2^{-2}$) | Pre-op Asphericity ($Z_4^{0}$) | 0.146 | -0.016 | 0.301 | 0.078 | 1.000 |
| Pre-op Astigm+ ($Z_2^{-2}$) | Post-op Astigm+ ($Z_2^{-2}$) | 0.026 | -0.136 | 0.187 | 0.751 | 1.000 |
| Pre-op Astigm+ ($Z_2^{-2}$) | Post-op Astigmx ($Z_2^{+2}$) | 0.138 | -0.025 | 0.293 | 0.097 | 1.000 |
| Pre-op Astigm+ ($Z_2^{-2}$) | Pre-op Astigmx ($Z_2^{+2}$) | -0.003 | -0.164 | 0.159 | 0.976 | 0.976 |
| Pre-op Astigm+ ($Z_2^{-2}$) | Post-op Asphericity ($Z_4^{0}$) | -0.114 | -0.271 | 0.048 | 0.167 | 1.000 |
| Pre-op Astigm+ ($Z_2^{-2}$) | Pre-op Asphericity ($Z_4^{0}$) | 0.090 | -0.073 | 0.248 | 0.278 | 1.000 |
| Post-op Astigmx ($Z_2^{+2}$) | Post-op Asphericity ($Z_4^{0}$) | 0.022 | -0.141 | 0.183 | 0.796 | 1.000 |
| Post-op Astigmx ($Z_2^{+2}$) | Pre-op Asphericity ($Z_4^{0}$) | -0.006 | -0.168 | 0.156 | 0.942 | 1.000 |
| Pre-op Astigmx ($Z_2^{+2}$) | Post-op Astigmx ($Z_2^{+2}$) | 0.267 | 0.110 | 0.411 | 0.001 | 0.028 |
| Pre-op Astigmx ($Z_2^{+2}$) | Post-op Asphericity ($Z_4^{0}$) | -0.106 | -0.263 | 0.057 | 0.201 | 1.000 |
| Pre-op Astigmx ($Z_2^{+2}$) | Pre-op Asphericity ($Z_4^{0}$) | 0.032 | -0.130 | 0.193 | 0.698 | 1.000 |
| Post-op Defocus ($Z_2^{0}$) | Post-op Astigm+ ($Z_2^{-2}$) | -0.006 | -0.168 | 0.156 | 0.942 | 1.000 |
| Post-op Defocus ($Z_2^{0}$) | Pre-op Astigm+ ($Z_2^{-2}$) | -0.224 | -0.373 | -0.065 | 0.006 | 0.151 |
| Post-op Defocus ($Z_2^{0}$) | Post-op Astigmx ($Z_2^{+2}$) | -0.005 | -0.167 | 0.157 | 0.950 | 1.000 |
| Post-op Defocus ($Z_2^{0}$) | Pre-op Astigmx ($Z_2^{+2}$) | -0.062 | -0.221 | 0.101 | 0.458 | 1.000 |
| Post-op Defocus ($Z_2^{0}$) | Post-op Asphericity ($Z_4^{0}$) | 0.249 | 0.090 | 0.395 | 0.002 | 0.060 |
| Post-op Defocus ($Z_2^{0}$) | Pre-op Asphericity ($Z_4^{0}$) | -0.111 | -0.268 | 0.052 | 0.181 | 1.000 |
| Pre-op Defocus ($Z_2^{0}$) | Post-op Astigm+ ($Z_2^{-2}$) | 0.155 | -0.007 | 0.309 | 0.060 | 1.000 |
| Pre-op Defocus ($Z_2^{0}$) | Pre-op Astigm+ ($Z_2^{-2}$) | 0.136 | -0.026 | 0.292 | 0.100 | 1.000 |
| Pre-op Defocus ($Z_2^{0}$) | Post-op Astigmx ($Z_2^{+2}$) | 0.084 | -0.079 | 0.243 | 0.311 | 1.000 |
| Pre-op Defocus ($Z_2^{0}$) | Pre-op Astigmx ($Z_2^{+2}$) | 0.171 | 0.009 | 0.324 | 0.039 | 0.889 |
| Pre-op Defocus ($Z_2^{0}$) | Post-op Defocus ($Z_2^{0}$) | 0.065 | -0.097 | 0.225 | 0.431 | 1.000 |
| Pre-op Defocus ($Z_2^{0}$) | Post-op Asphericity ($Z_4^{0}$) | -0.644 | -0.730 | -0.538 | 0.000 | 0.000 |
| Pre-op Defocus ($Z_2^{0}$) | Pre-op Asphericity ($Z_4^{0}$) | 0.088 | -0.075 | 0.246 | 0.291 | 1.000 |
| Pre-op Asphericity ($Z_4^{0}$) | Post-op Asphericity ($Z_4^{0}$) | 0.401 | 0.256 | 0.529 | 0.000 | 0.000 |
